# Identifying Priority Countries for Scaling Up Small-Quantity Lipid-Based Nutrient Supplements

**DOI:** 10.1101/2025.02.11.25322040

**Authors:** Navideh Noori, Christine P. Stewart, Christine M. McDonald, K. Ryan Wessells, Elisabeth D. Root, Kathryn G. Dewey

**Affiliations:** Institute for Disease Modeling, Gates Foundation, 500 5th Ave N, Seattle, WA, USA, 98109; Institute for Global Nutrition, Department of Nutrition, University of California, Davis, Davis, CA, U.S.A.; Departments of Pediatrics, and Epidemiology and Biostatistics, University of California, San Francisco, CA, USA

**Keywords:** malnutrition, stunting, wasting, all-cause mortality, children 6-23 months old, SQ-LNS, preventative supplement, scale-up

## Abstract

**Introduction:** Undernutrition is a cause of nearly half of all deaths among children under-5. Small-quantity lipid-based nutrient supplements (SQ-LNS) have been shown to prevent child wasting, stunting, anemia and mortality among children 6-23 months of age in low- and middle-income countries (LMICs). Scaling up effective preventive interventions is urgent given the current global food insecurity and nutrition crisis.

**Method:** To prioritize SQ-LNS scale-up activities, we identified countries with the highest burdens of wasting, stunting, and all-cause mortality among children 6-23 months of age at the national level using the most recent national survey data including the Demographic and Health Survey (DHS) and Multiple Indicator Cluster Surveys (MICS), as well as the Lives Saved Tool (LiST) in LMICs. National-level estimates informed a care cascade model to assess the potential impact of SQ-LNS on all-cause mortality, stunting, and wasting. We also conducted a sub-national level analysis among the 20 highest burden countries with the most recent available survey data to identify the highest burden regions.

**Results:** Our analysis identified the top 20 countries with the highest burden of the three outcomes as: Niger, South Sudan, Yemen, Sudan, Somalia, Democratic Republic of Congo, Eritrea, Nigeria, Central African Republic, Guinea, Equatorial Guinea, Chad, Papua New Guinea, Benin, Mali, Angola, Pakistan, Timor-Leste, Sierra Leone and Côte d’Ivoire, although for some countries the survey data were collected > 10 years ago. Some of these countries also ranked high in population estimates of acute food insecurity. The care cascade model demonstrates that a large number of cases of stunting and wasting and deaths could be potentially averted if SQ-LNS is provided.

**Conclusion:** Most of the top 20 countries are in Sub-Saharan Africa, with a few in South and Southeast Asia. This geographical concentration underscores the urgent need for targeted interventions in these regions to prevent child malnutrition.

**What is already known on this topic:** It is known that SQ-LNS reduces child wasting, stunting, anemia and mortality among children 6-23 months of age in LMICs. However, survey reports generally estimate stunting, wasting, and mortality rates among all children under 5 years of age, and not specifically for the 6-23 months old age group.

**What this study adds:** By estimating the burden of stunting, wasting and all-cause mortality for the specific age group of 6-23 months old at the national and sub-national level, we identified high burden countries and regions where SQ-LNS will be most impactful. No other study has focused on this age group even though it is a particularly vulnerable period for undernutrition.

**How this study might affect research, practice or policy:** Our analysis could help decision-makers and funders to determine where scale-up of SQ-LNS should be prioritized.

## Introduction

Undernutrition is a cause of nearly half of all deaths in children under 5 years of age [1]. In 2022, stunting affected an estimated 22.3 % or 148.1 million children under 5 years of age globally and wasting affected 6.8% or 48 million [2]. The Sustainable Development Goals (SDGs) target 2.2 calls for an end to all forms of malnutrition by 2025, with a 40% reduction in stunting (relative to 2012), and for wasting to occur in less than 5% of children [3]. There is an urgent need to scale-up effective actions to improve child nutrition. However, only one out of three wasted children receive the care they need due to insufficient resources and an uncoordinated system [4]. In addition to increasing funding for the early detection and treatment of child wasting, the prevention of wasting and other forms of child undernutrition should also be prioritized [5]. However, there are limited effective strategies for the prevention of undernutrition.

Provision of small-quantity lipid-based nutrient supplements (SQ-LNS) is a relatively new preventive intervention that was added to the list of recommended interventions to effectively address child undernutrition [6], and is one of the few interventions that can help to achieve multiple SDG targets concurrently [7]. SQ-LNS are small packets of nutrient-dense paste that supplement the diets of children 6-23 months of age with more than 20 essential vitamins and minerals and essential fatty acids to support healthy growth and development. They have been shown to reduce child wasting, stunting, anemia and mortality among children 6-23 months of age in low- and middle-income countries (LMICs) [7,8]. For certain outcomes, the greatest impact was seen in sites with higher burdens of stunting or wasting, or poorer water quality or sanitation. The benefit-cost ratio of SQ-LNS was estimated to be 13.7 times when targeted to the children 6-23 months of age in the 60% of the population with the lowest socio-economic status (SES) in the 40 LMICs with the highest prevalences of stunting [9]. In its recent guidelines on (1) prevention and management of wasting and nutritional oedema (acute malnutrition) in infants and children under 5 years, and (2) complementary feeding for infants and young children 6-23 months of age, the World Health Organization (WHO) recommended the use of SQ-LNS in food insecure populations [10,11]. UNICEF also emphasizes the importance of targeting the youngest children in the most vulnerable households and in food insecure settings with high burden of wasting and stunting and micronutrient deficiencies [12].

Provision of SQ-LNS is not a stand-alone intervention but rather should be considered as part of a package of interventions and integrated into existing programs and services including household food assistance in settings with high levels of food insecurity [12,13]. Identifying the most vulnerable populations and targeting provision of SQ-LNS to these groups could optimize its impact [7]. The goal of our work is to identify countries with a high burden of stunting, wasting and all-cause mortality among children aged 6-23 months old, for prioritizing the settings where SQ-LNS will be most impactful.

Given that blanket and geographical targeting may be the most appropriate strategy for preventive programs in high-risk populations [14], we also conducted a similar analysis at the sub-national level for selected countries, using recent survey data to identify the highest burden regions within those countries.

## Methods

### National-level analysis

To identify priority countries for the scale-up of SQ-LNS, we estimated the prevalence of moderate and severe wasting, and moderate and severe stunting and the all-cause mortality rate among children aged 6-23 months old at the national level in LMICs in Sub-Saharan Africa, South and Southeast Asia, as well as Middle east, North Africa, Central and South America regions, based on the World Bank classifications [15]. These estimates were necessary since survey reports generally report stunting, wasting, and mortality rates among all children under 5 years of age, and not specifically for the 6-23 months old age group. Moderate stunting is defined as length-for-age z score -3 ≤ (LAZ) < −2, and severe stunting is defined as LAZ < -3. Moderate wasting is defined as weight-for-length z score -3 ≤ (WLZ) < −2, and severe wasting is defined as WLZ < -3 [16].

To estimate the moderate and severe stunting prevalence, we used survey estimates from the WHO Child Malnutrition database for 2021 among children 24-59 months old [17]. We chose the 24-59 months age interval to capture the total burden of stunting up to 24 months of age, since the stunting prevalence increases sharply during the 6-23 months age interval. After 24 months, the prevalence of stunting generally stabilizes, and the estimate for 24-59 months would be a good approximation of stunting that has occurred prior to 24 months of age. To derive the most recent estimates for each country, we developed a regression model between survey-based stunting estimates for children aged 24–59 months and the annual national-level Joint Malnutrition Estimate (JME) estimates [2] for children under five for years 2000 to 2021 to predict 2021 stunting prevalence among children aged 24–59 months. JME also reports annual estimates of wasting prevalence among children under-5 years old at the regional level. However, exploratory analyses indicated that there were no consistent trends in the prevalence of wasting across years that could be used to develop a regression model, as was done for stunting. Therefore, we used the most recent national survey data including the Demographic and Health Survey (DHS), Multiple Indicator Cluster Surveys (MICS), and WHO Child Malnutrition Estimates to estimate the prevalence of moderate and severe wasting among children 6-23 months of age. This is consistent with global trends, which are downward for stunting during the past 20 years, but not for wasting prevalence.

We estimated the all-cause mortality rate per 1,000 live births among children 6-23 months of age from the Lives Saved Tool (LiST). LiST is a computer-based application for modeling the impact of maternal and child health interventions and calculates changes in cause-specific mortality based on intervention coverage change, intervention effectiveness for that cause, and the percentage of cause-specific mortality sensitive to that intervention [18]. LiST child mortality estimates are developed by the UN Inter-Agency Group for Child Mortality Estimation (UN-IGME) [19]. UN-IGME estimates mortality by fitting a smooth trend curve over values from DHS, MICS, vital registration data and population census data.

After calculating prevalence estimates of moderate and severe wasting, and moderate and severe stunting, as well as the all-cause mortality rate among children aged 6-23 months old, we ranked countries for each outcome. We then created a composite score by averaging the ranks across the following three outcomes: all-cause mortality rate, prevalence of severe wasting, and prevalence of severe stunting. We also developed a separate composite score based on the all-cause mortality rate, the prevalence of stunting, and the prevalence of wasting. We referred to the Integrated Food Security Phase Classification (IPC) scale for countries that rank high in acute food insecurity [20].

We further developed a care cascade or decision tree model to estimate the potential effects of provision of SQ-LNS on reducing each outcome at the national level using the national-level prevalence estimates and mortality rate estimates. The decision tree model was developed for the top 20 countries based on the composite scores. We estimated the national-level population of children aged 6-23 months old by obtaining the population of children under-1 and 1 to 2 years old from the World Population Prospect [21]. We divided the under-1 population by half, subtracted neonatal deaths (from UN-IGME) and deaths among one to 6 months old (∼ 5* (infant deaths – neonatal deaths)/11) and then summed the result with the population of 1 to 2 years old children. We accounted for uncertainty in the simulation by employing Monte Carlo simulation techniques and defining a range of values for the prevalence and mortality estimates, as well as for relative reduction in risk of each outcome among those who receive SQ-LNS based on the meta-analysis of randomized controlled trials of SQ-LNS provided to children 6-23 months of age [7,8,22]. For the prevalence estimates of wasting and stunting, we used confidence intervals from the source of each estimate (DHS, MICS, WHO Child Malnutrition Database, WHO-JME predicted estimates), and for the relative reduction in risk of each outcome, we used the estimated confidence intervals from the meta-analyses [7,8,22] and generated a matrix of 1000 sample points for each outcome and relative reductions within the given range using a Latin Hypercube sample (LHS) [23]. The all-cause mortality estimates from LiST do not include confidence interval [7,8,22]. The percentage of children aged 6-23 months old who would receive SQ-LNS in the models varied between 0 and 100%. Descriptions of the model parameters and the decision model structure are given in Table 1 and Figure 1.

**Table 1.** Decision tree models parameters description.

| Parameter description | Parameter | Value |
| --- | --- | --- |
| Percentage of children aged 6-23 months old who will receive SQ-LNS | $\rho$ | 0%-100% |
| Prevalence of moderate stunting and moderate wasting among children 6-23 months old | $\varepsilon_s, \varepsilon_w$ | Baseline estimates |
| Prevalence of severe stunting and severe wasting among children 6-23 months old | $\theta_s, \theta_w$ | Baseline estimates |
| Prevalence of children aged 6-23 months old who are not stunted, and not wasted | $\eta_s = 1 - \varepsilon_s - \theta_s$<br>$\eta_w = 1 - \varepsilon_w - \theta_s$ | Baseline estimates |
| All-cause mortality among children aged 6-23 months old | $\phi$ | Baseline estimates |
| Crude prevalence ratio of moderate stunting [7] | - | 0.86 (0.83, 0.89) |
| Crude prevalence ratio of severe stunting [8] | - | 0.83 (0.78, 0.90) |
| Crude prevalence ratio of moderate wasting [7] | - | 0.87 (0.80, 0.95) |
| Crude prevalence ratio of severe wasting [8] | - | 0.69 (0.55, 0.86) |
| Crude prevalence ratio of all-cause mortality [7] | - | 0.73 (0.59, 0.89) |

**Figure 1.**
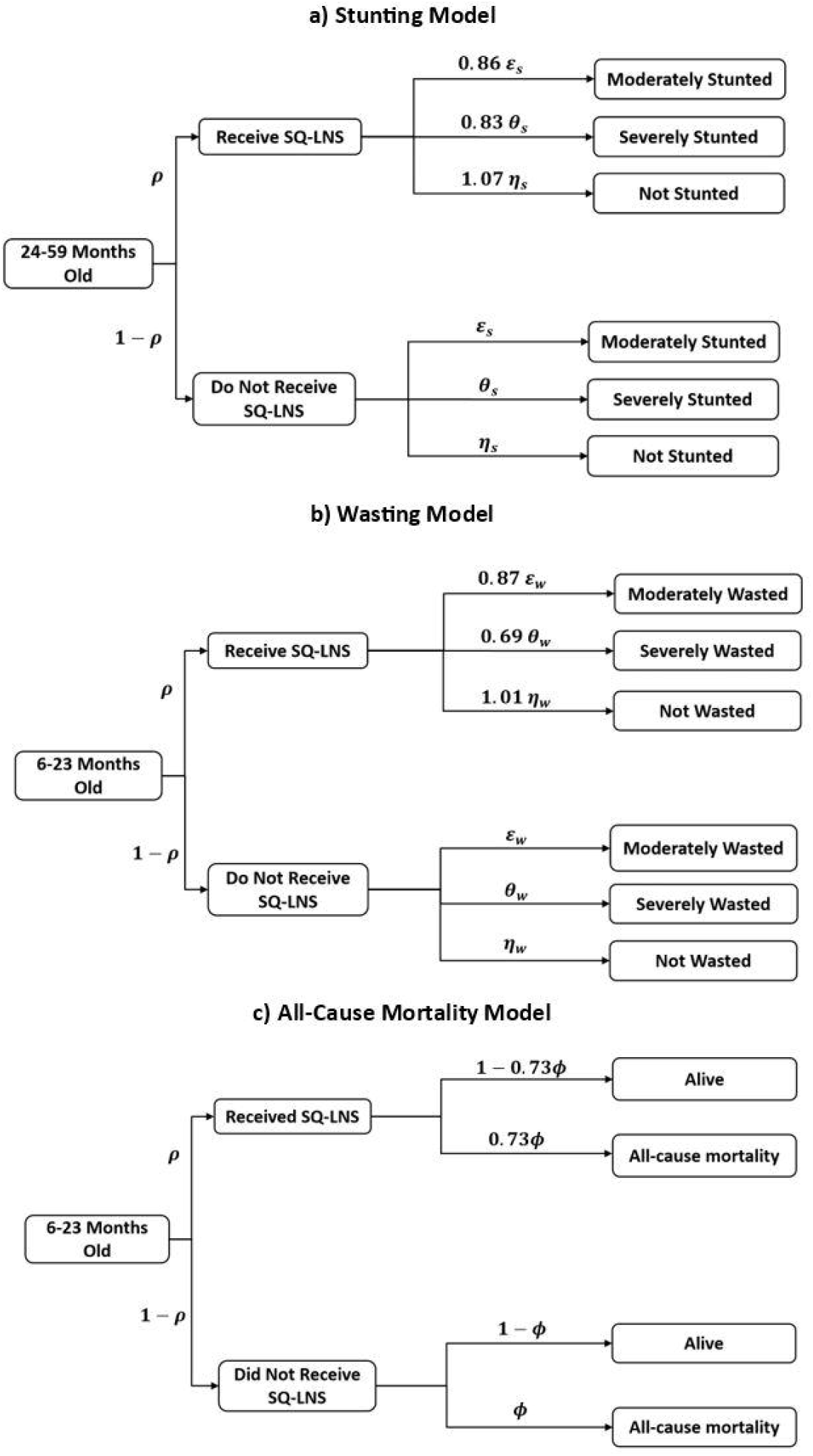
Schematic of decision tree model of a) stunting, b) wasting and c) all-cause mortality.

### Sub-national-level analysis

We conducted a similar analysis at the sub-national level among the top 20 countries with the highest composite scores based on severe stunting, severe wasting and all-cause mortality with the most recent available survey data from Demographic and Health Surveys (DHS) or Multiple Indicator Cluster Surveys (MICS) over the past 10 years to identify the highest burden regions within a country. A list of countries with recent survey data is given in Table S1.

## Results

### Heatmap Analysis

The baseline prevalence estimates of severe wasting among children aged 6 to 23 months old based on the most recent survey data, prevalence estimates of severe stunting among children aged 24-59 months old for the year 2021, and the all-cause mortality rate among children aged 6 to 23 months per 1,000 live births for 2021 from LiST are given in Figure 2. For each outcome, the top 50 countries with the highest burden are shown. Estimates for other LMICs are shown in Figure S1. The y-axis for severe wasting shows the survey year used to estimate prevalence for each country. We ranked countries for each of these outcomes and averaged the ranks across the three outcomes. For each outcome, the ranking value ranges between 1 to 119 representing the number of countries included in this analysis. Figure 3 shows the heatmap of the composite score of countries and Figure S2 shows rankings in a bar graph. The composite score ranges between 1 to 115, and the higher the score is, the higher the mortality rate and the prevalence estimates of severe wasting and severe stunting. Niger, South Sudan, Yemen, Sudan, and Somalia are the top five countries with the highest composite scores. The composite score we developed based on the mortality rate, prevalence of stunting, and prevalence of wasting, showed similar results (Figures S3 & S4).

**Figure 2.**
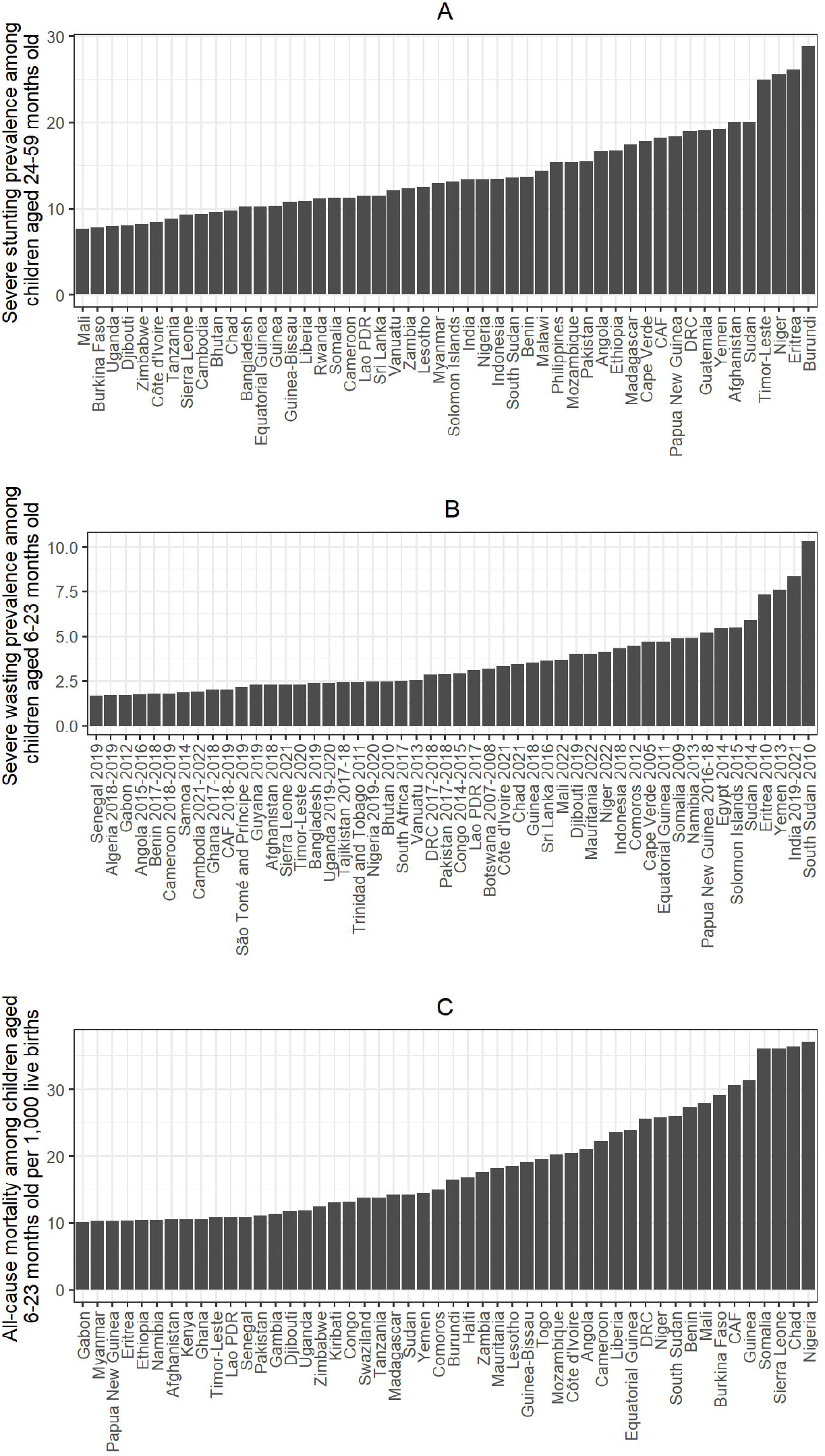
A) Prevalence of severe stunting among children aged 24-59 months old for the year 2021, B) Prevalence of severe wasting among children aged 6 to 23 months old based on recent survey data, and C) All-cause mortality among children aged 6 to 23 months per 1,000 live births for the year 2021.

**Figure 3.**
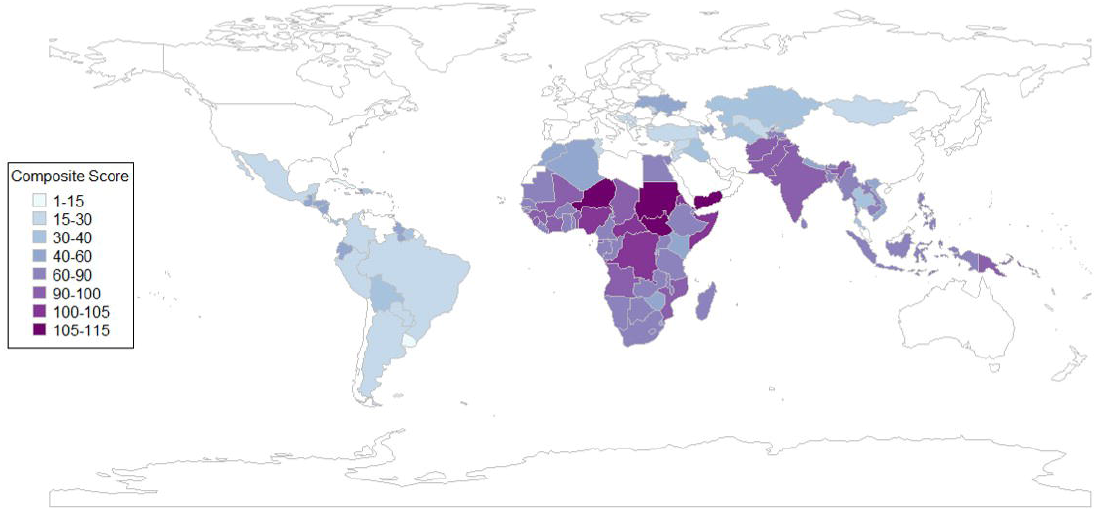
Heatmap of composite score of countries based on severe stunting, severe wasting and all-cause mortality.

Our analysis suggests that 14 countries rank in the top 20 for at least two of the three outcomes: Niger, South Sudan, Yemen, Sudan, Somalia, Democratic Republic of Congo, Eritrea, Central African Republic, Guinea, Equatorial Guinea, Papua New Guinea, Benin, Mali, and Angola, in rank order, although for some countries the survey data were collected over 10 years ago. Figure S6 shows the countries with IPC phases 3 and higher for acute food insecurity in at least 20% of the country population. The x-axis in Figure S6 shows the most recent period for which IPC severity was estimated for each country. Among these countries, Yemen and South Sudan, with over 50% IPC phase 3 and above, experienced the highest acute food insecurity. The IPC rankings, however, focus on food insecurity often in countries focusing humanitarian emergencies, whereas our rankings are based on mortality rates and the prevalence of child undernutrition among children 6-23 months old. Therefore, not all highly ranked countries in our analysis were ranked highly in IPC estimates.

### Decision Tree Model of Potential Effects of SQ-LNS on Stunting, Wasting and All-Cause Mortality

We estimated the potential downstream benefits of provision of SQ-LNS on the burden of stunting, wasting and all-cause mortality at the national level using the previously described decision tree model. We present the effect as the absolute numbers as well as percentage point reduction for each adverse outcome at the national level. Our simulation suggests that over 1.2 million cases of moderate stunting could be potentially averted, if half of children aged 6-23 months old received SQ-LNS in the top 20 countries. Similarly, with 50% coverage, over 930,000 cases of severe stunting, over 200,000 cases of moderate wasting, over 165,000 cases of severe wasting, and over 90,000 deaths could be potentially averted (Table 2).

**Table 2.** Total number of cases for each outcome and potential number of cases of wasting and stunting and number of deaths averted, if half or all of children aged 6-23 months old in the 20 countries with the highest composite scores (based on prevalence of severe stunting, severe wasting and all-cause mortality) receive SQ-LNS.

|  | Total number of cases for each outcome without provision of SQ-LNS | SQ-LNS Coverage |  |
| --- | --- | --- | --- |
|  |  | 50% | 100% |
| Moderate Wasting | 3,440,339 ± 11,668* | 215,005 ± 1,834 | 430,011 ± 3,668 |
| Severe Wasting | 1,120,618 ± 2,754 | 165,255 ± 1,198 | 330,510 ± 2,396 |
| Moderate Stunting | 17,176,930 ± 66,712 | 1,202,495 ± 5,772 | 2,404,990 ± 11,545 |
| Severe Stunting | 11,638,660 ± 7,521 | 931,033 ± 4,542 | 1,862,068 ± 9,085 |
| All-Cause Mortality | 718, 416 ± 0 | 93,393 ± 695 | 186,787 ± 1,391 |

\* mean ± standard error

### Sub-national-level analysis

The full sub-national rankings and prevalence estimates for severe wasting, severe stunting, and mortality are provided in the Supplementary Material. These highlight the regions with the highest composite scores for each outcome within each country. Sudan, DRC and Nigeria are the highest burden countries with recent survey data. In Nigeria, using DHS 2018, the states of Kebbi, Sokoto and Katsina have the highest composite score with highest mortality rate and highest prevalence of severe stunting and severe wasting among children aged 6-23 months old. In DRC, using MICS 2017, Sankuru and Maniema are the highest burden provinces. In Sudan, using MICS 2014, the states of Gadarif, central and north Darfor have highest burden of the three outcomes.

## Discussion

We identified countries with the highest burdens of stunting, wasting and all-cause mortality among children 6-23 months of age, with the objective of identifying candidate countries for the scale-up of SQ-LNS. Developing an approach to estimate these burdens in this age group is an important contribution of these analyses. This age group, the period of complementary feeding [10], is a key target age group for multiple nutrition programs, although data specific to this period are not reported in global burden statistics. A focus on this age group is critical given that rates of linear growth faltering and wasting are highest during this period [24].

Some of the top-ranked countries in our analysis also rank highly in the IPC population estimates of acute food insecurity. Prioritizing the top-ranked countries is thus in alignment with the WHO guideline recommendations on the use of SQ-LNS in food insecure populations [10]. The World Bank has also provided recommendations for effective design and implementation of SQ-LNS interventions in the Sahel region to improve child nutrition and health in this region, where high levels of child undernutrition are persistent [13]. Some of the Sahelian countries such as Niger, Nigeria, Mali, Guinea and Chad were ranked in our analysis as the countries with the highest burden of wasting, stunting and mortality among children aged 6-23 months old. The recent World Bank Investment Framework for Nutrition listed SQ-LNS as one of the high-priority interventions for scalable strategies and emphasized that scaling up SQ-LNS requires identifying vulnerable populations and integrating SQ-LNS into existing programs [25].

The results of the decision tree model demonstrate that a large number of children could potentially benefit from provision of SQ-LNS in terms of reduced all-cause mortality, moderate and severe stunting, and moderate and severe wasting at the national level. For example, in Niger, which had highest mortality rate and highest prevalence of severe wasting and severe stunting, provision of SQ-LNS to all children aged 6-23 months could potentially avert over 15,000 cases of severe wasting, over 116,000 cases of severe stunting, and over 7,000 deaths per year. We recognize that 100% coverage of SQ-LNS is not realistic, but by providing a range of values for the coverage, we show the potential number of children’s lives that could be improved and saved in each country given a specific level of SQ-LNS coverage. It should be noted that the estimated relative risk reductions in severe wasting and severe stunting in those models may be underestimates, given that more substantial relative risk reductions were observed in the higher-burden sites in the individual participant data (IPD) analysis, e.g., a 48% reduction in severe wasting in sites with poor water quality [8]. Therefore, the number of cases of severe wasting and severe stunting potentially averted could be higher than shown herein.

One key limitation of our analysis is that the survey data we used to estimate wasting prevalence in some countries were collected over 10 years ago. Also, we used cross-sectional surveys in our analysis, which underestimates the incidence and total burden of wasting in comparison with longitudinal cohort data [26,27]. In addition, we assumed that the effect of SQ-LNS on each adverse outcome in our simulation was linear, and we did not account for the potentially nonlinear dynamic interplay between this preventive intervention and wasting and stunting prevalence. Future work is needed to develop a dynamic model of wasting and stunting and to account for the potential nonlinear tradeoff between treatment and preventive interventions as well as the cost-effectiveness of provision of SQ-LNS in reducing the treatment demand and overall burden of undernutrition.

## Conclusion

Our analysis identified countries with a high burden of undernutrition and mortality among children 6-23 months old, with the majority located in Sub-Saharan Africa and a few in South and Southeast Asia. These findings highlight the urgent need for targeted interventions in these regions. Scaling up of SQ-LNS, integrated into existing programs and services, could play a critical role in reducing undernutrition and improving child health outcomes. While our analysis is not designed for direct programmatic decision-making within countries, it provides information to help decision-makers and funders identify priority countries and sub-regions for SQ-LNS scale-up.

## Data Availability

This study was based on secondary analysis of Demographic and Health Surveys (DHS) and Multiple Indicator Cluster Surveys (MICS), and WHO Child Malnutrition Estimates which are available online.

## Contributors

KGD, CMM, CPS and KRW helped develop the research concept and approach. NN did the statistical analysis and wrote the initial draft of the manuscript. EDR provided a technical review of the analysis. All authors contributed to the interpretation of results and writing of the manuscript and have read and approved the final manuscript.

## Ethics Approval

This study was based on secondary analysis of Demographic and Health Surveys (DHS) and Multiple Indicator Cluster Surveys (MICS), and WHO Child Malnutrition Estimates. The ethical clearance was provided by the Institutional Review Board of ICF International. Therefore, this secondary analysis was exempt from ethical review approval, since it used publicly available, de-identified data.

## Supplementary Material Supplementary Material.pdf

