## Supplementary Material for "Identifying Priority Countries for Scaling Up Small-Quantity Lipid-Based Nutrient Supplements"

Navideh Noori^*^

^1^ Institute for Disease Modeling, Gates Foundation, 500 5th Ave N, Seattle, WA, USA, 98109.

^2^ Institute for Global Nutrition, Department of Nutrition, University of California, Davis, Davis, CA, U.S.A.,

^3^ Departments of Pediatrics, and Epidemiology and Biostatistics, University of California, San Francisco, CA, USA

**Table S1**. Top 20 countries with the highest composite scores developed based on severe stunting, severe wasting and all-cause mortality with recent survey data available from Demographic and Health Surveys (DHS) or Multiple Indicator Cluster Surveys (MICS) over the past 10 years for the sub-national level analysis.

**Section A.** National level Analysis

**Figure S1**. Prevalence of severe stunting among children aged 24-59 months old for the year 2021, prevalence of severe wasting among children aged 6 to 23 months old based on recent survey data, and all-cause mortality among children aged 6 to 23 months per 1,000 live births for the year 2021 among LMICs.

**Figure S2**. Ranking of countries by highest mortality rate and highest prevalence of severe stunting and severe wasting.

**Figure S3**. Heatmap of composite score of countries based on stunting, wasting and all-cause mortality.

**Figure S4**. Ranking of countries by highest mortality rate and highest prevalence of stunting and wasting.

**Figure S5**. Ranking of countries by highest mortality rate and highest prevalence of moderate and severe stunting and moderate and severe wasting.

**Figure S6**. Countries with IPC severity phases 3 and higher for acute food insecurity. The classification was conducted at the sub-national level and represents the highest severity affecting at least 20% of the population and has five phases of 1. Minimal, 2. Stressed, 3. Crisis, 4. Emergency, and 5. Famine.

**Section B**. Decision Tree Model of Potential Effects of SQ-LNS on Stunting, Wasting and All-Cause Mortality

**Figure S7**. Number of cases of a & b) moderate and severe stunting, c & d) moderate and severe wasting and e) number of deaths that could be potentially averted among children aged 6-23 months old in the 20 countries with the highest composite scores based on prevalence of severe stunting, severe wasting and all-cause mortality.

**Figure S8**. Percentage point reduction in a & b) moderate and severe stunting prevalence, c & d) moderate and severe wasting prevalence and e) all-cause mortality rate per 1,000 live births among children aged 6-23 months old in top 20 countries.

**Section C.** Subnational level analysis

**Figure S9.** Sub-national prevalence of severe stunting among children aged 24-59 months old, prevalence of severe wasting among children aged 6 to 23 months old, and all-cause mortality among children aged 6 to 23 months per 1,000 live births based on most recent MICS survey in Sudan.

**Figure S10**. Sub-national prevalence of severe stunting among children aged 24-59 months old, prevalence of severe wasting among children aged 6 to 23 months old, and all-cause mortality among children aged 6 to 23 months per 1,000 live births based on most recent MICS survey in Democratic Republic of Congo.

**Figure S11**. Sub-national prevalence of severe stunting among children aged 24-59 months old, prevalence of severe wasting among children aged 6 to 23 months old, and all-cause mortality among children aged 6 to 23 months per 1,000 live births based on the most recent DHS survey in Nigeria.

**Figure S12**. Sub-national prevalence of severe stunting among children aged 24-59 months old, prevalence of severe wasting among children aged 6 to 23 months old, and all-cause mortality among children aged 6 to 23 months per 1,000 live births based on most recent MICS survey in Central African Republic.

**Figure S13**. Sub-national prevalence of severe stunting among children aged 24-59 months old, prevalence of severe wasting among children aged 6 to 23 months old, and all-cause mortality among children aged 6 to 23 months per 1,000 live births based on the most recent DHS survey in Guinea. Figure S14. Sub-national prevalence of severe stunting among children aged 24-59 months old, prevalence of severe wasting among children aged 6 to 23 months old, and all-cause mortality among children aged 6 to 23 months per 1,000 live births based on most recent MICS survey in Chad.

**Figure S15**. Sub-national prevalence of severe stunting among children aged 24-59 months old, prevalence of severe wasting among children aged 6 to 23 months old, and all-cause mortality among children aged 6 to 23 months per 1,000 live births based on the most recent DHS survey in Papua New Guinea.

**Figure S16**. Sub-national prevalence of severe stunting among children aged 24-59 months old, prevalence of severe wasting among children aged 6 to 23 months old, and all-cause mortality among children aged 6 to 23 months per 1,000 live births based on the most recent DHS survey in Benin.

**Figure S17**. Sub-national prevalence of severe stunting among children aged 24-59 months old, prevalence of severe wasting among children aged 6 to 23 months old, and all-cause mortality among children aged 6 to 23 months per 1,000 live births based on the most recent DHS survey in Mali.

**Figure S18**. Sub-national prevalence of severe stunting among children aged 24-59 months old, prevalence of severe wasting among children aged 6 to 23 months old, and all-cause mortality among children aged 6 to 23 months per 1,000 live births based on the most recent DHS survey in Pakistan.

**Figure S19**. Sub-national prevalence of severe stunting among children aged 24-59 months old, prevalence of severe wasting among children aged 6 to 23 months old, and all-cause mortality among children aged 6 to 23 months per 1,000 live births based on the most recent DHS survey in Timor-Leste.

**Figure S20**. Sub-national prevalence of severe stunting among children aged 24-59 months old, prevalence of severe wasting among children aged 6 to 23 months old, and all-cause mortality among children aged 6 to 23 months per 1,000 live births based on the most recent DHS survey in Sierra Leone.

**Figure S21**. Sub-national prevalence of severe stunting among children aged 24-59 months old, prevalence of severe wasting among children aged 6 to 23 months old, and all-cause mortality among children aged 6 to 23 months per 1,000 live births based on the most recent DHS survey in Angola.

**Figure S22.** Sub-national prevalence of severe stunting among children aged 24-59 months old, prevalence of severe wasting among children aged 6 to 23 months old, and all-cause mortality among children aged 6 to 23 months per 1,000 live births based on the most recent DHS survey in Côte d’Ivoire.

**Figure S23**. Sub-national ranking of Sudan by highest mortality rate and highest prevalence of severe stunting and severe wasting as well as heatmap of composite score at the sub-national level.

**Figure S24**. Sub-national ranking of Democratic Republic of Congo by highest mortality rate and highest prevalence of severe stunting and severe wasting as well as heatmap of composite score at the sub-national level.

**Figure S25**. Sub-national ranking of Nigeria by highest mortality rate and highest prevalence of severe stunting and severe wasting as well as heatmap of composite score at the sub-national level.

**Figure S26**. Sub-national ranking of Central African Republic by highest mortality rate and highest prevalence of severe stunting and severe wasting as well as heatmap of composite score at the sub-national level.

**Figure S27**. Sub-national ranking of Guinea by highest mortality rate and highest prevalence of severe stunting and severe wasting as well as heatmap of composite score at the sub-national level.

**Figure S28**. Sub-national ranking of Chad by highest mortality rate and highest prevalence of severe stunting and severe wasting as well as heatmap of composite score at the sub-national level.

**Figure S29**. Sub-national ranking of Papua New Guinea by highest mortality rate and highest prevalence of severe stunting and severe wasting as well as heatmap of composite score at the sub-national level.

**Figure S30**. Sub-national ranking of Benin by highest mortality rate and highest prevalence of severe stunting and severe wasting as well as heatmap of composite score at the sub-national level.

**Figure S31**. Sub-national ranking of Mali by highest mortality rate and highest prevalence of severe stunting and severe wasting as well as heatmap of composite score at the sub-national level.

**Figure S32**. Sub-national ranking of Pakistan by highest mortality rate and highest prevalence of severe stunting and severe wasting as well as heatmap of composite score at the sub-national level.

**Figure S33**. Sub-national ranking of Timor-Leste by highest mortality rate and highest prevalence of severe stunting and severe wasting as well as heatmap of composite score at the sub-national level.

**Figure S34**. Sub-national ranking of Sierra Leone by highest mortality rate and highest prevalence of severe stunting and severe wasting as well as heatmap of composite score at the sub-national level.

**Figure S35**. Sub-national ranking of Angola by highest mortality rate and highest prevalence of severe stunting and severe wasting as well as heatmap of composite score at the sub-national level.

**Figure S36**. Sub-national ranking of Côte d’Ivoire by highest mortality rate and highest prevalence of severe stunting and severe wasting as well as heatmap of composite score at the sub-national level.

Table S1. Top 20 countries with the highest composite scores developed based on severe stunting, severe wasting and all-cause mortality with recent survey data available from Demographic and Health Surveys (DHS) or Multiple Indicator Cluster Surveys (MICS) over the past 10 years for the sub-national level analysis

| Country | Data Source and Year |
| --- | --- |
| Sudan | MICS 2014 |
| Democratic Republic of Congo | MICS 2017 |
| Nigeria | DHS 2018 |
| Central African Republic | MICS 2018 |
| Guinea | DHS 2018 |
| Chad | MICS 2019 |
| Papa New Guinea | DHS 2016-18 |
| Benin | DHS 2017-18 |
| Mali | DHS 2018 |
| Pakistan | DHS 2017-18 |
| Timor-Leste | DHS 2016 |
| Sierra Leone | DHS 2019 |
| Angola | DHS 2015 |
| Côte d’Ivoire | DHS 2021 |

**Section A. National Level Analysis**

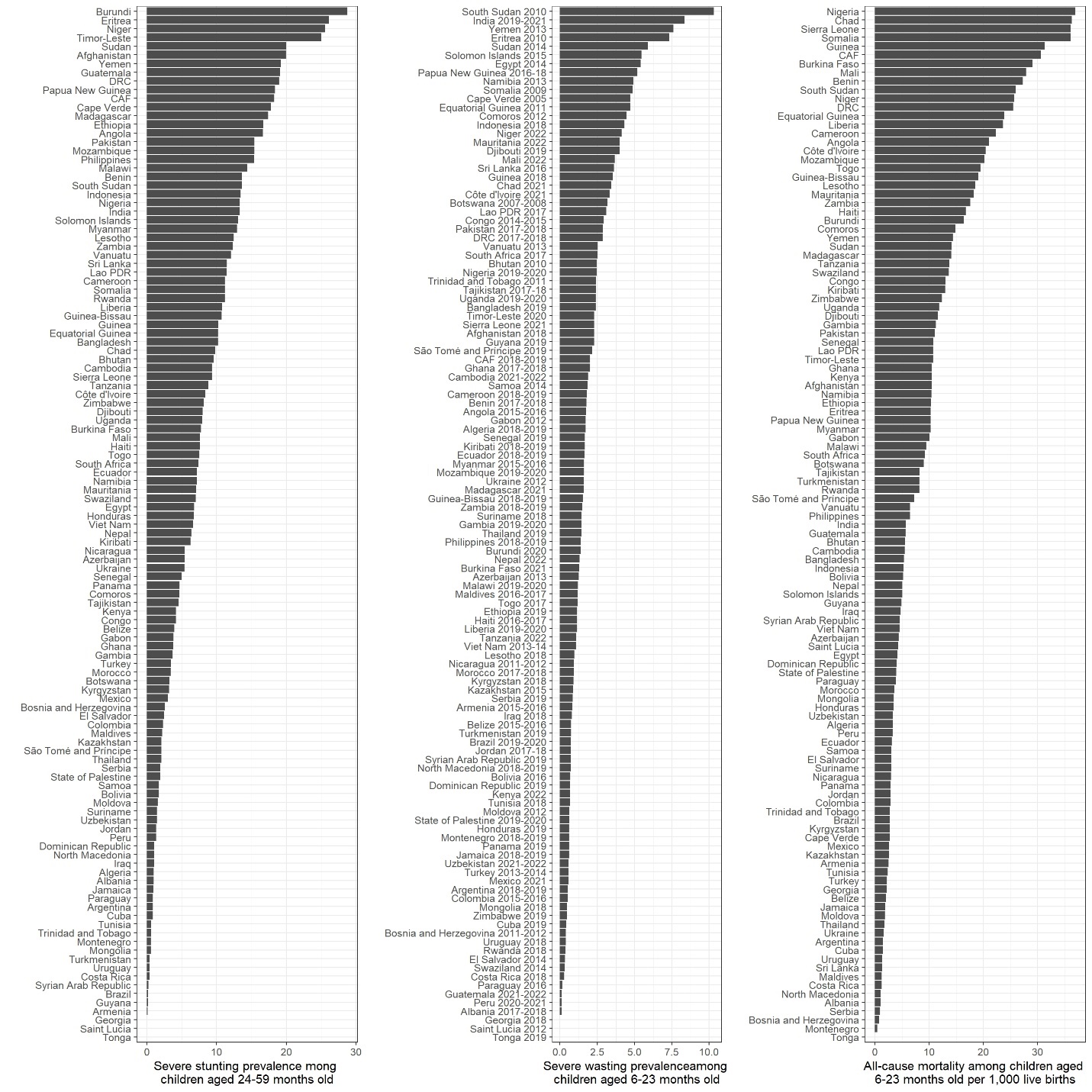

Figure S1. Prevalence of severe stunting among children aged 24-59 months old for the year 2021, prevalence of severe wasting among children aged 6 to 23 months old based on recent survey data, and all-cause mortality among children aged 6 to 23 months per 1,000 live births for the year 2021 among LMICs.

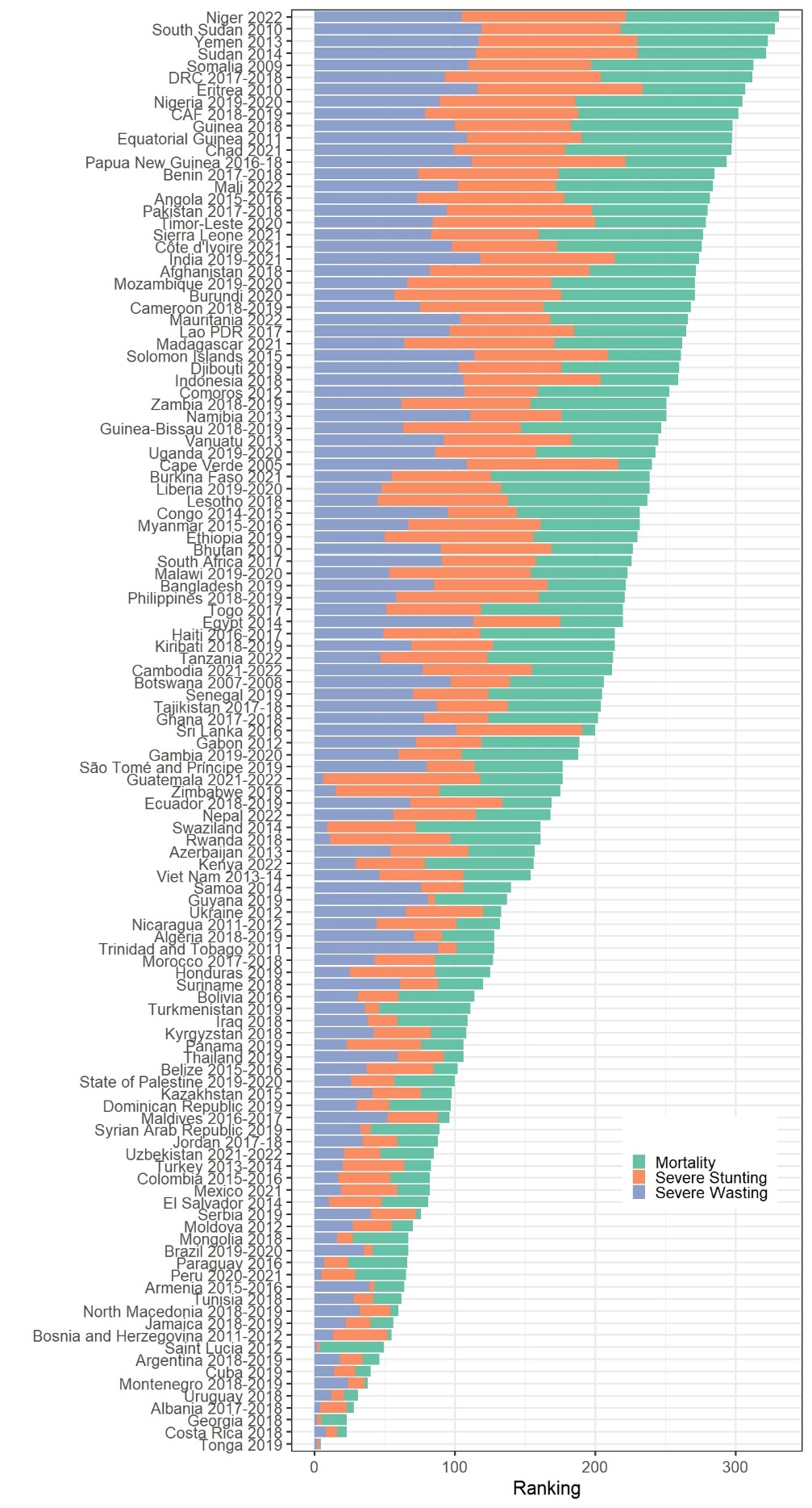

Figure S2. Ranking of countries by highest mortality rate and highest prevalence of severe stunting and severe wasting

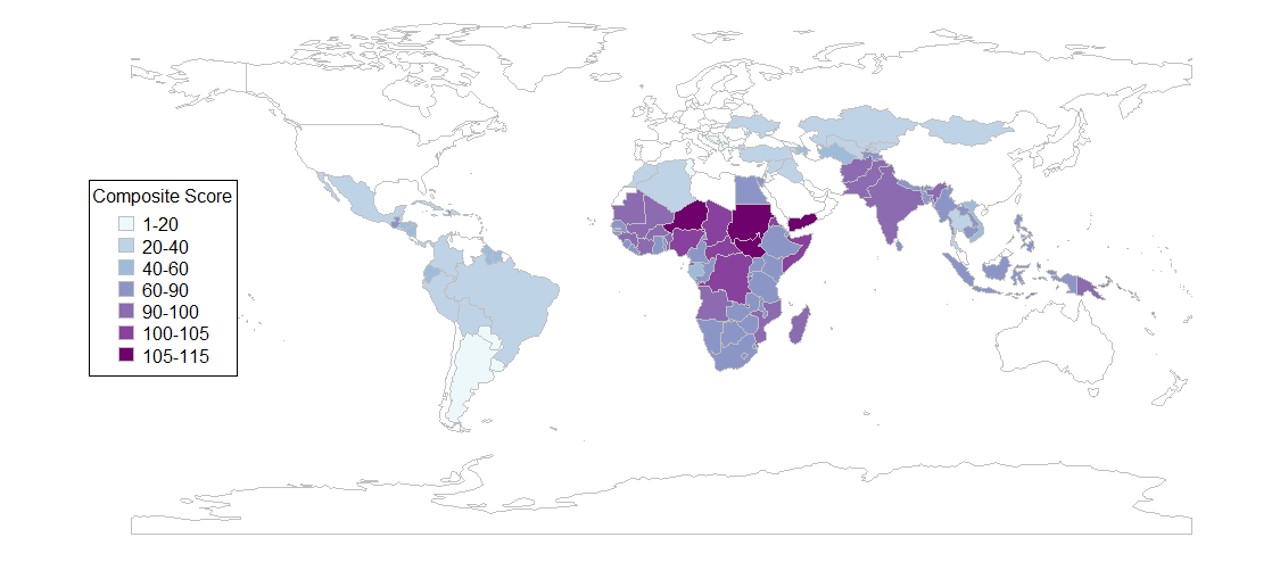

Figure S3. Heatmap of composite score of countries based on stunting, wasting and all-cause mortality.

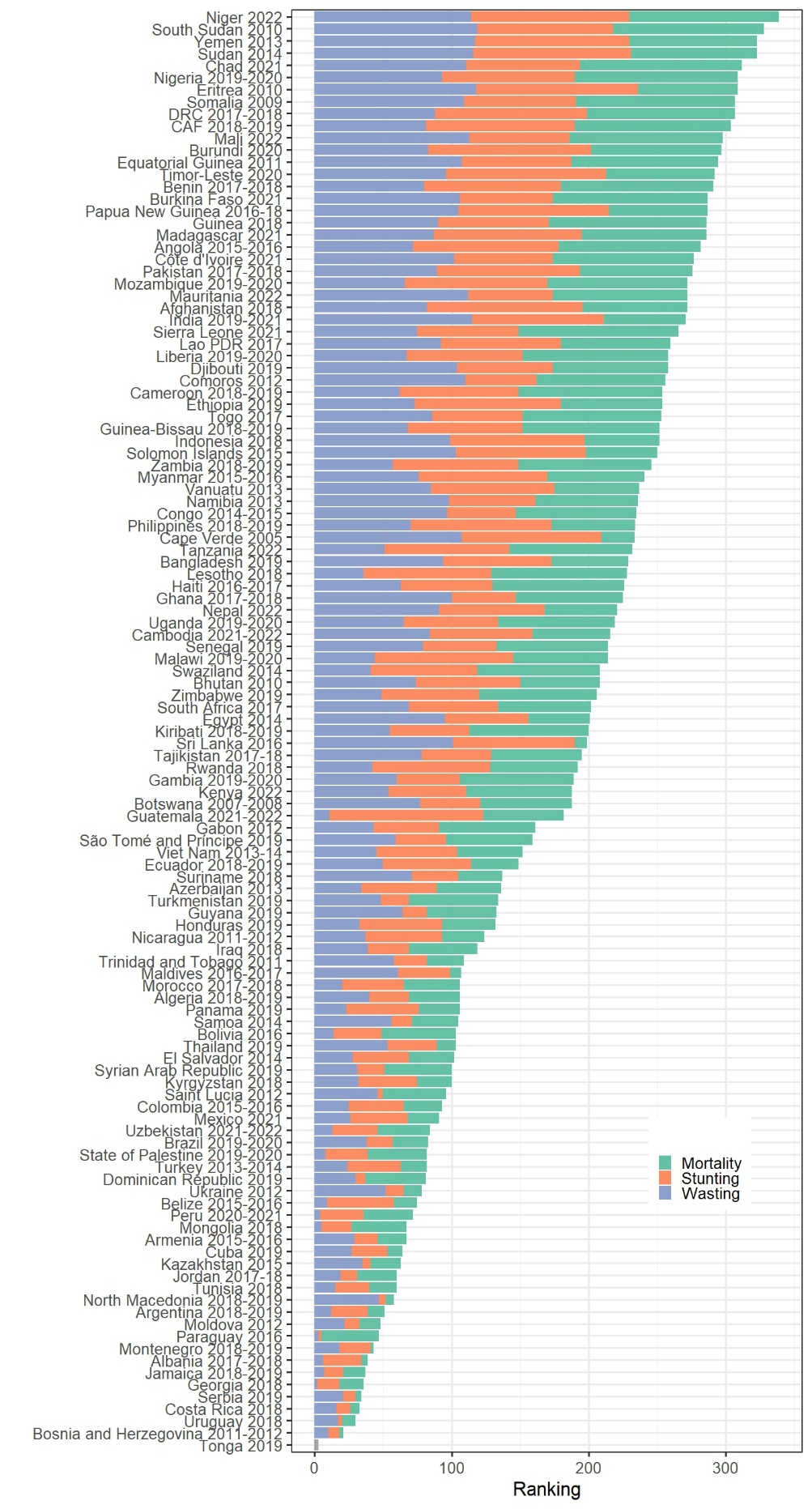

Figure S4. Ranking of countries by highest mortality rate and highest prevalence of stunting and wasting

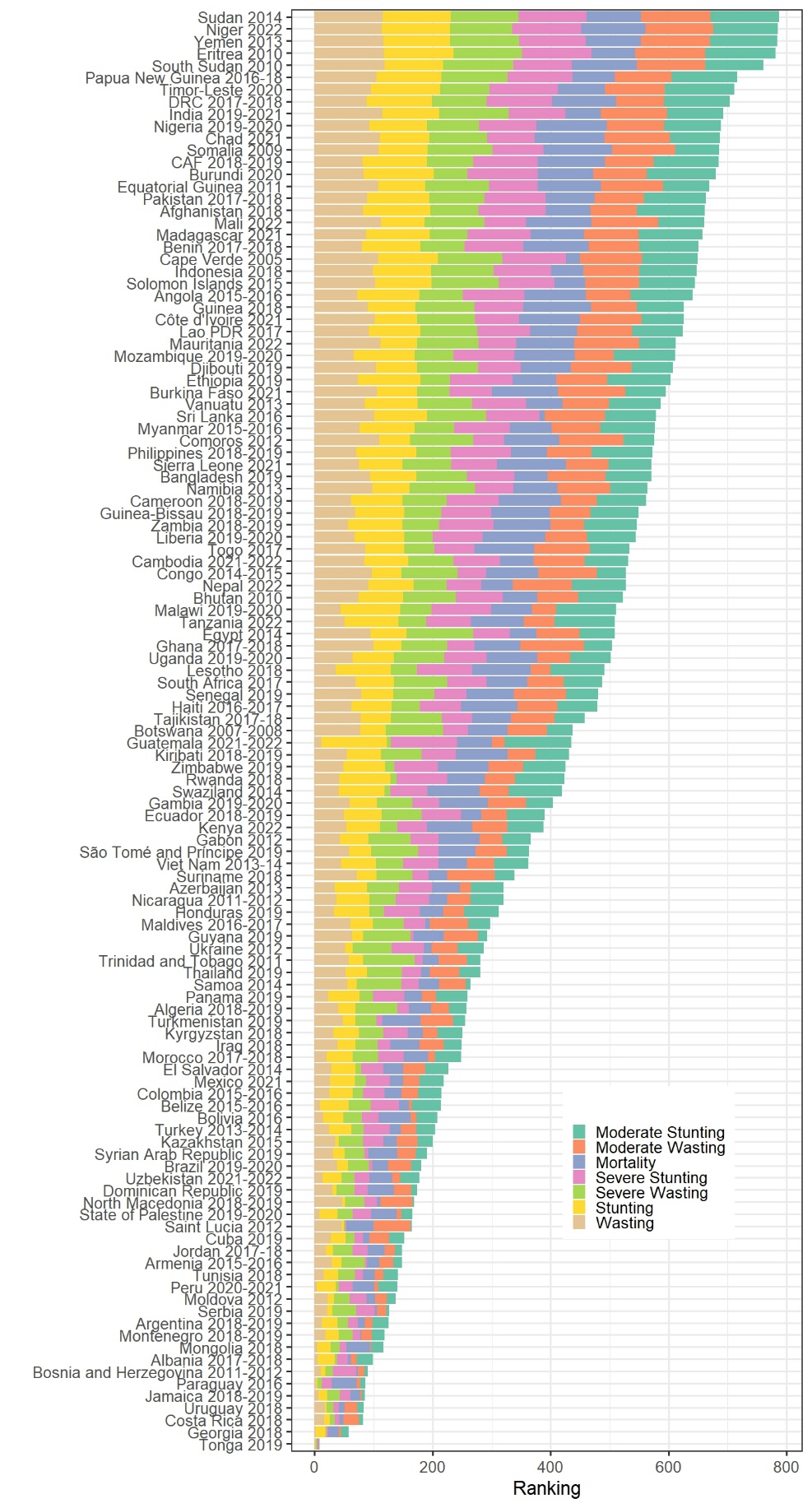

Figure S5. Ranking of countries by highest mortality rate and highest prevalence of moderate and severe stunting and moderate and severe wasting

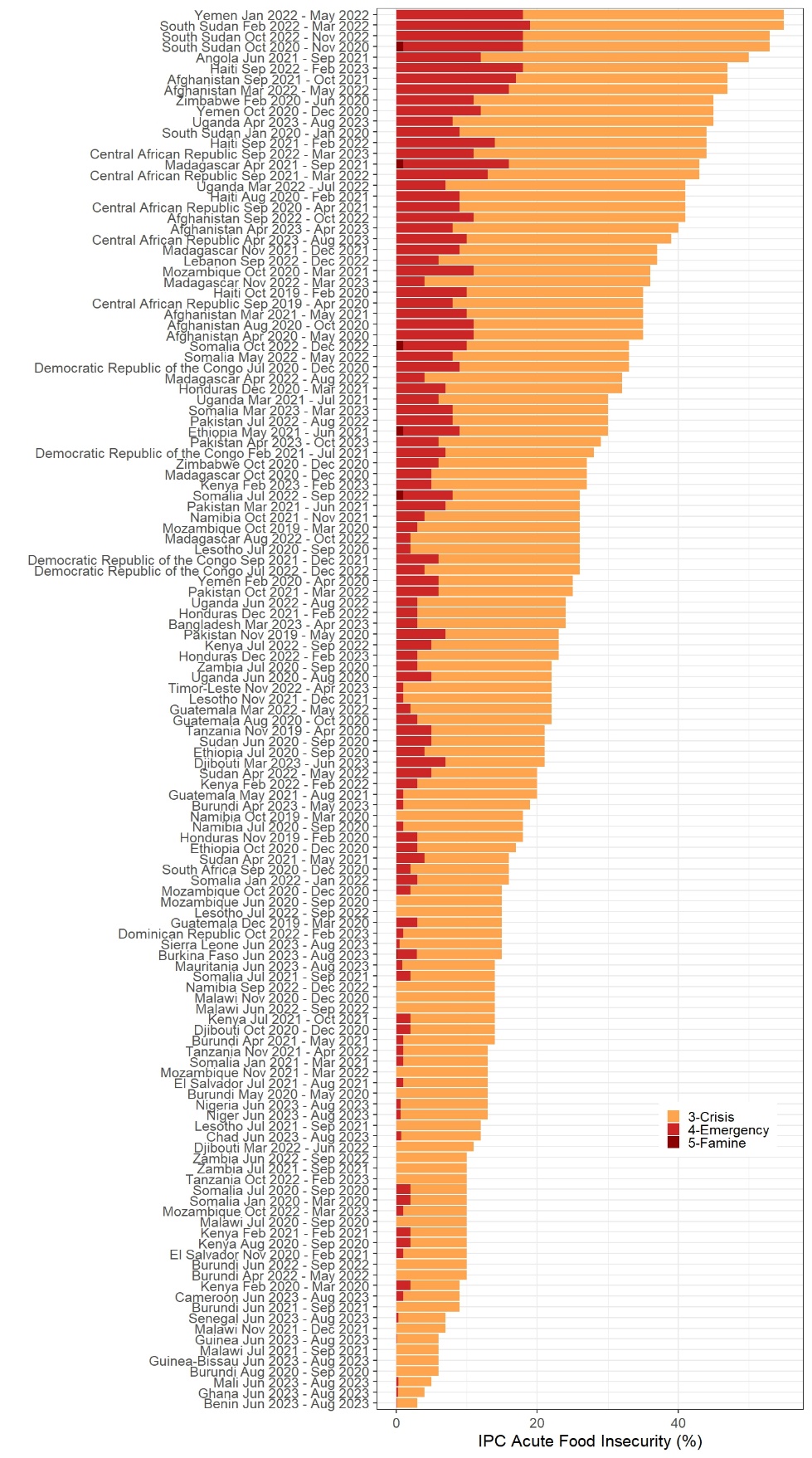

Figure S6. Countries with IPC severity phases 3 and higher for acute food insecurity. The classification was conducted at the sub-national level and represents the highest severity affecting at least 20% of the population and has five phases of 1. Minimal, 2. Stressed, 3. Crisis, 4. Emergency, and 5. Famine.

### **Section B. Decision Tree Model of Potential Effects of SQ-LNS on Stunting, Wasting and All-Cause Mortality**

The number of cases of moderate and severe wasting and moderate and severe stunting and number of deaths that could be potentially averted among children aged 6-23 months in the 20 countries with the highest composite scores are given in Figure S7 a-e. The x-axis shows SQ-LNS coverage varying between 0 to 100%. The percentage point reduction for each adverse outcome is also given in Figure S8 a-e.

a)

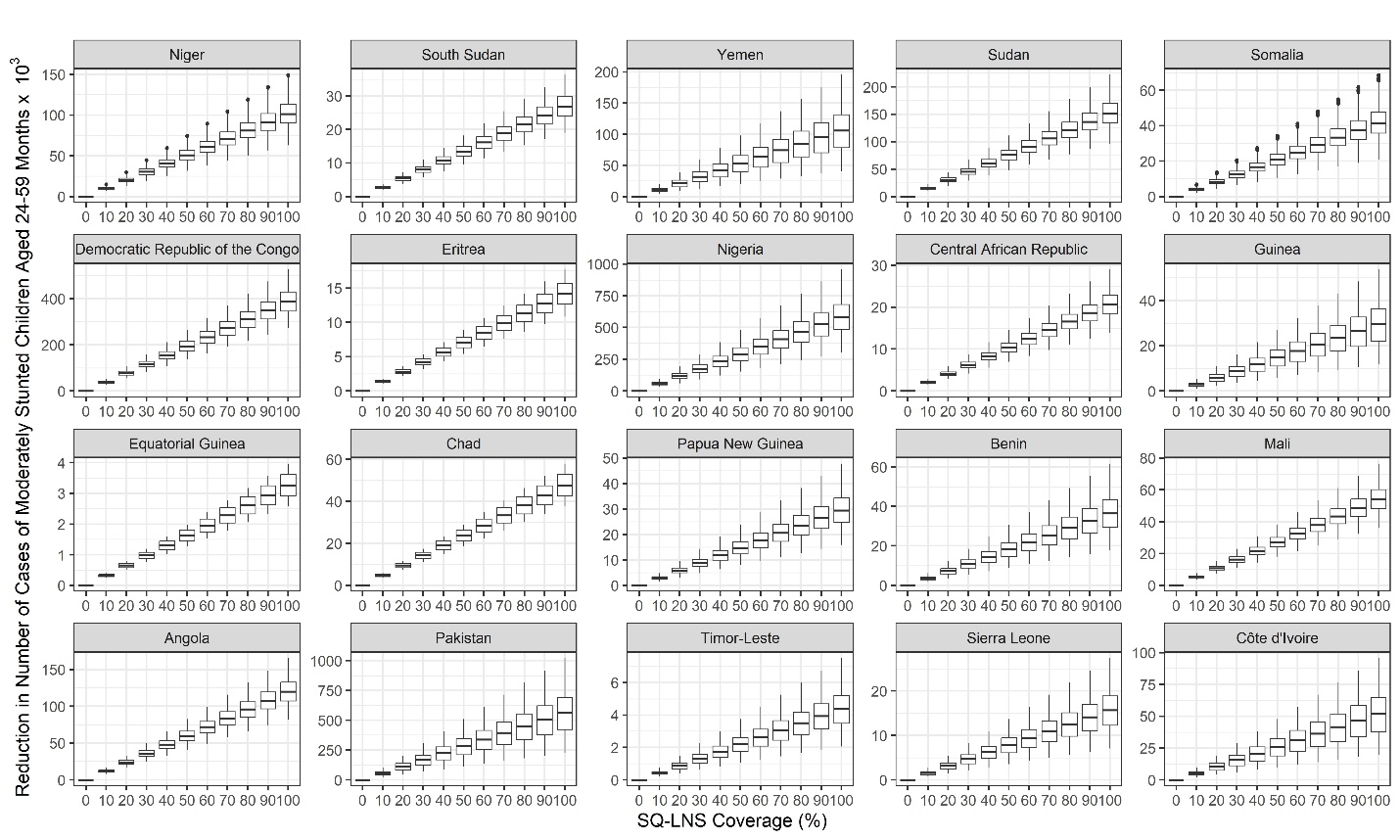

b)

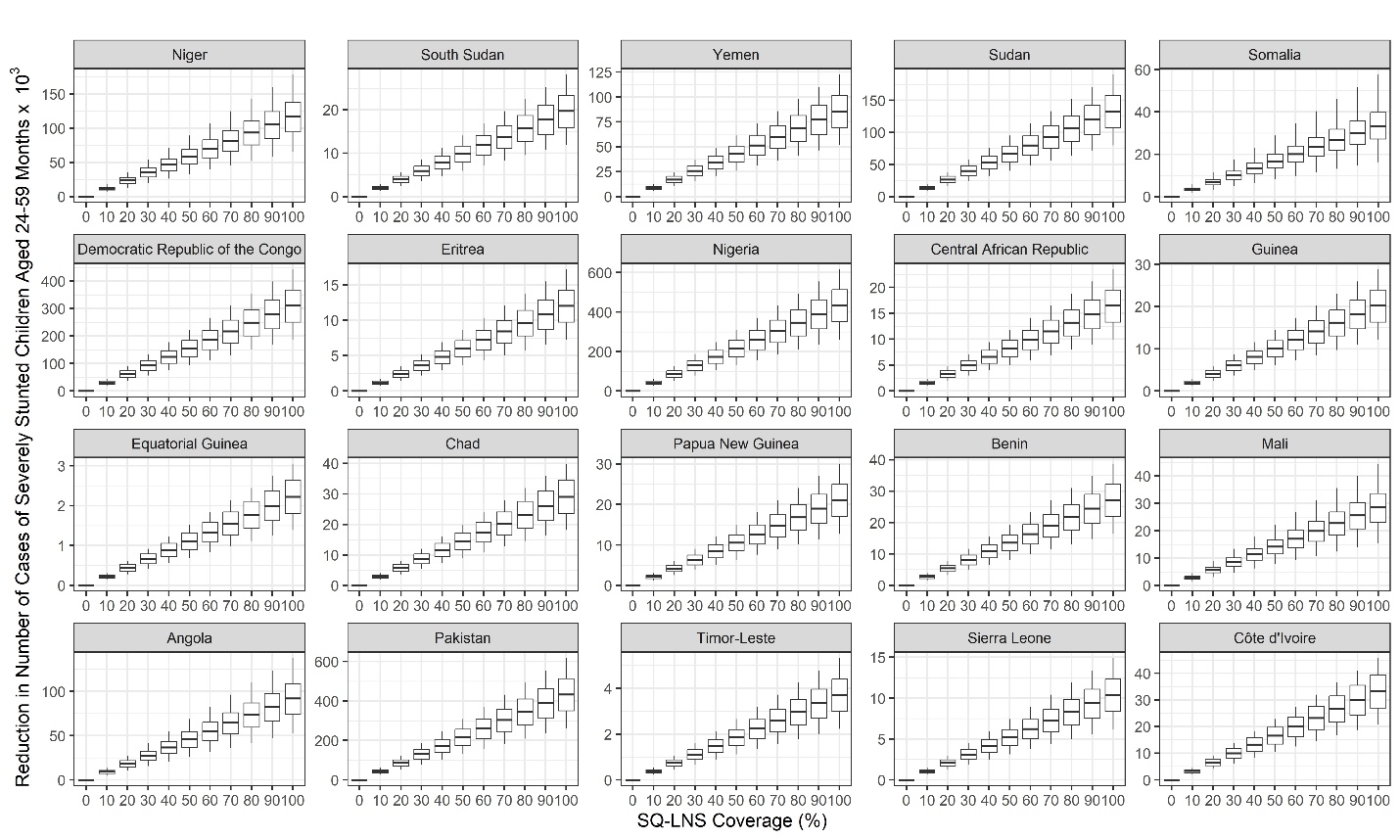

c)

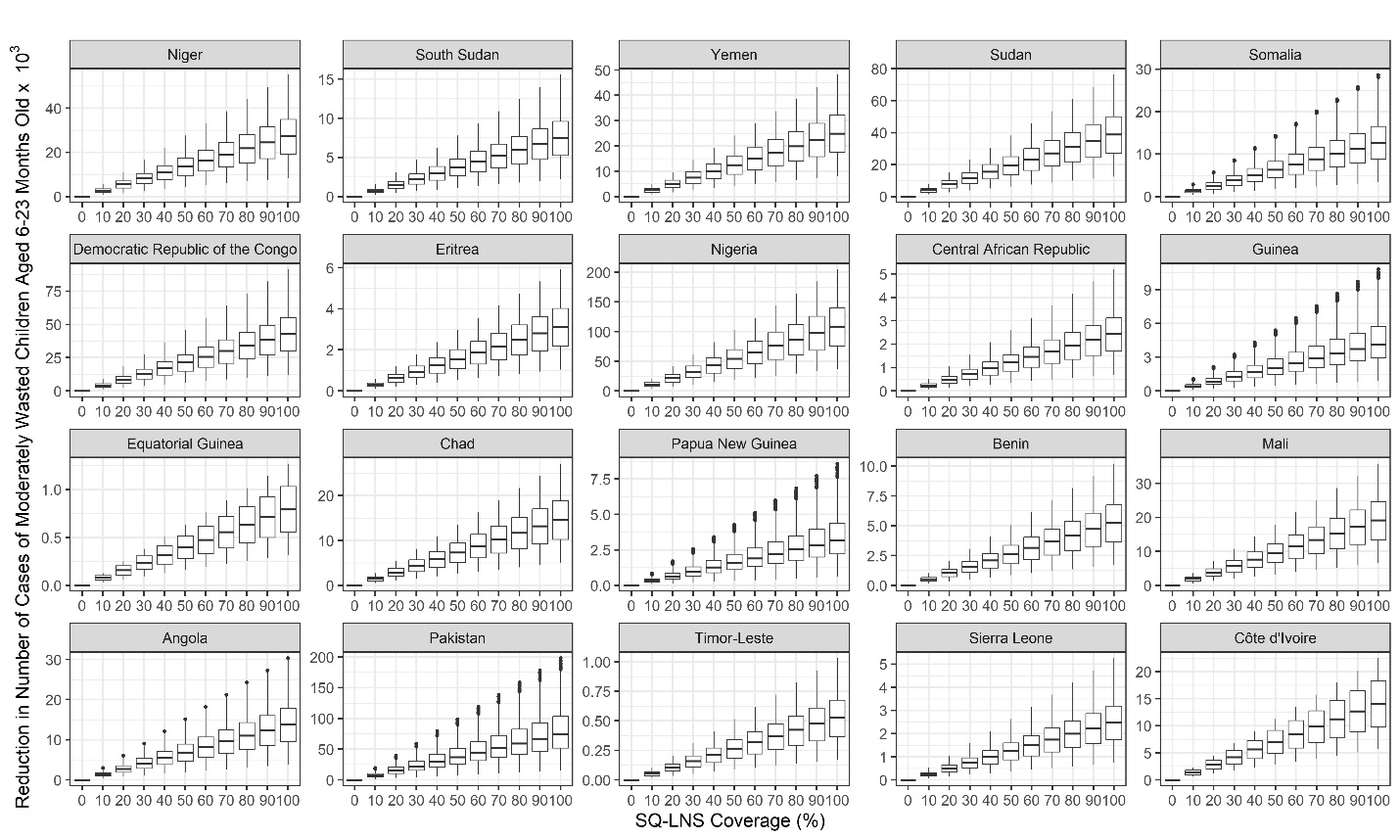

d)

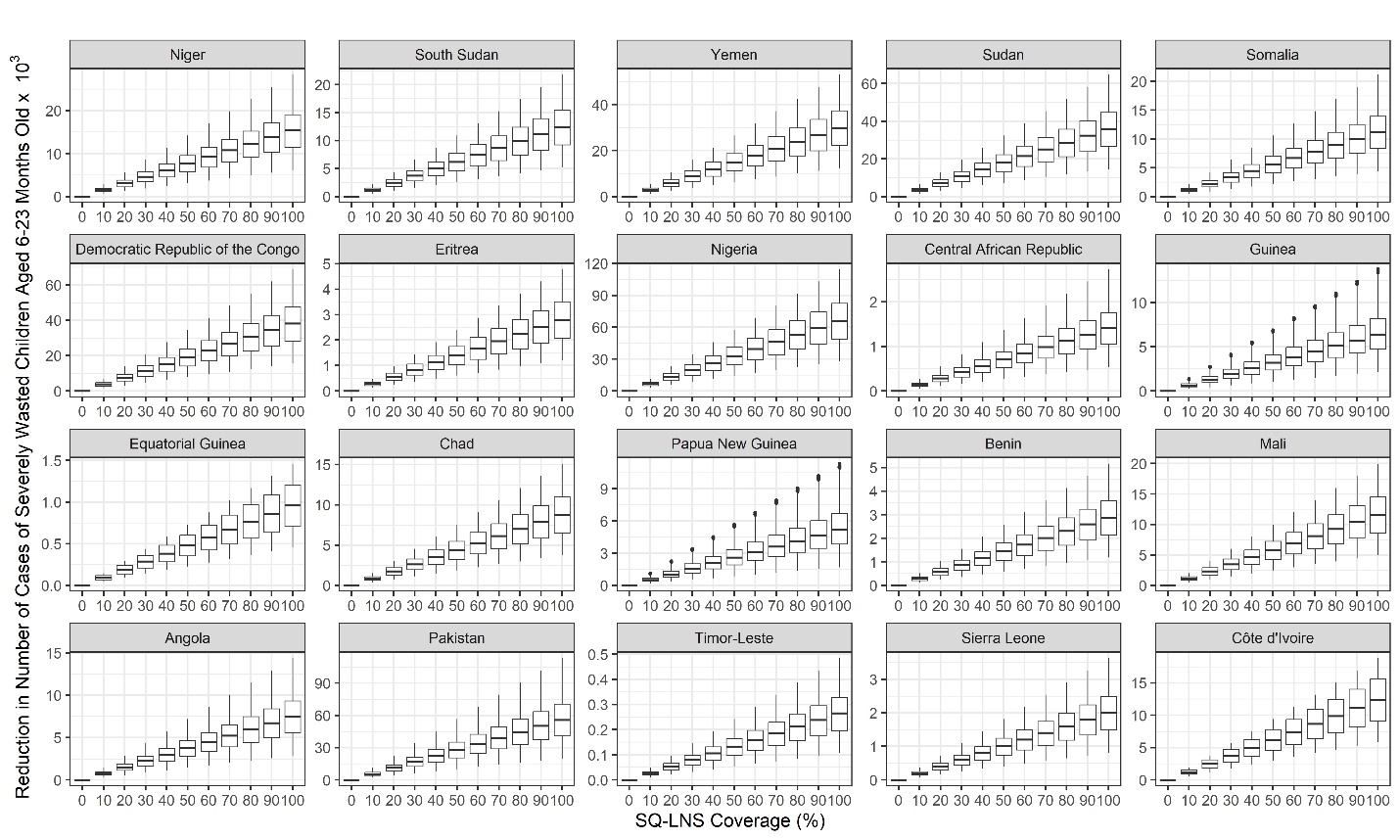

e)

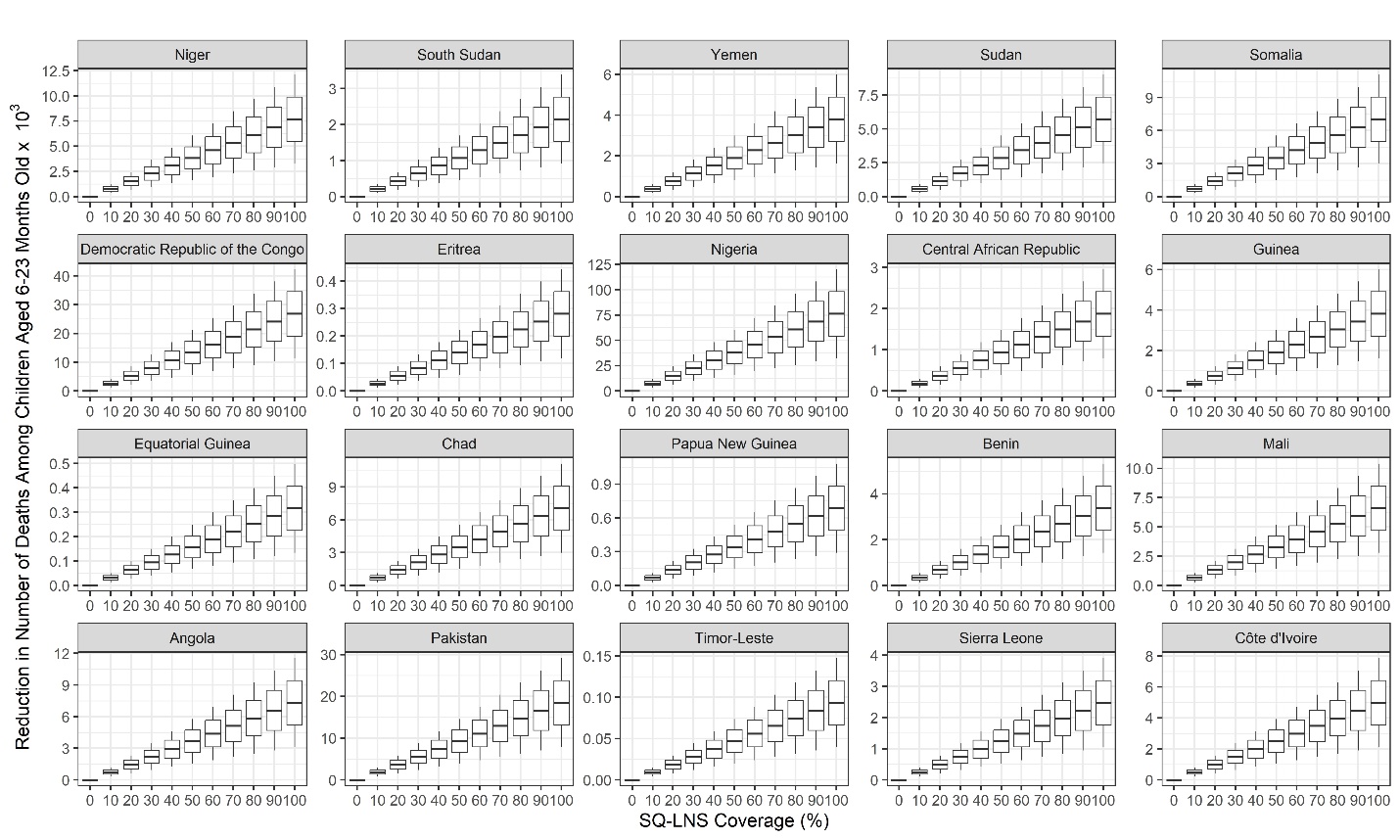

Figure S7. Number of cases of a & b) moderate and severe stunting, c & d) moderate and severe wasting and e) number of deaths that could be potentially averted among children aged 6-23 months old in the 20 countries with the highest composite scores based on prevalence of severe stunting, severe wasting and all-cause mortality.

a)

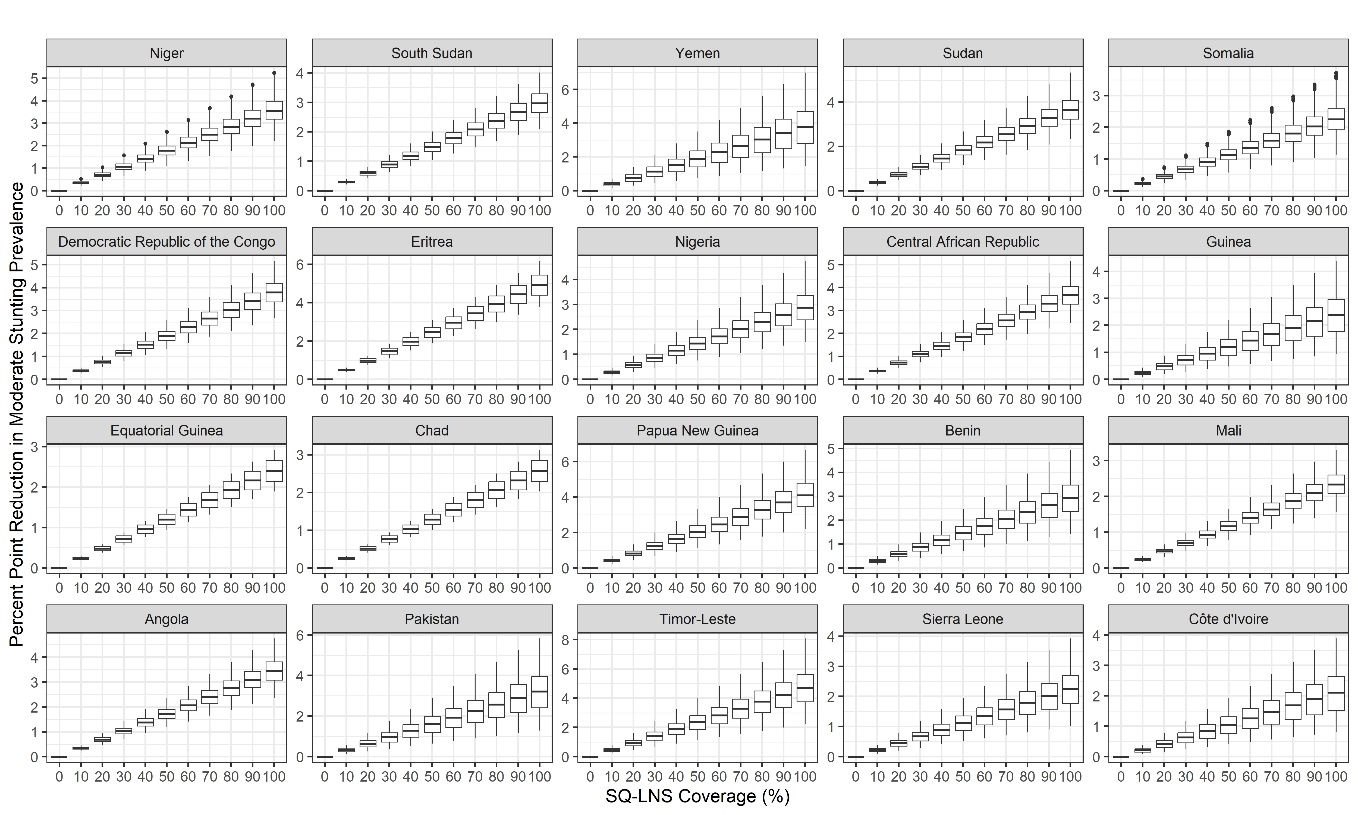

b)

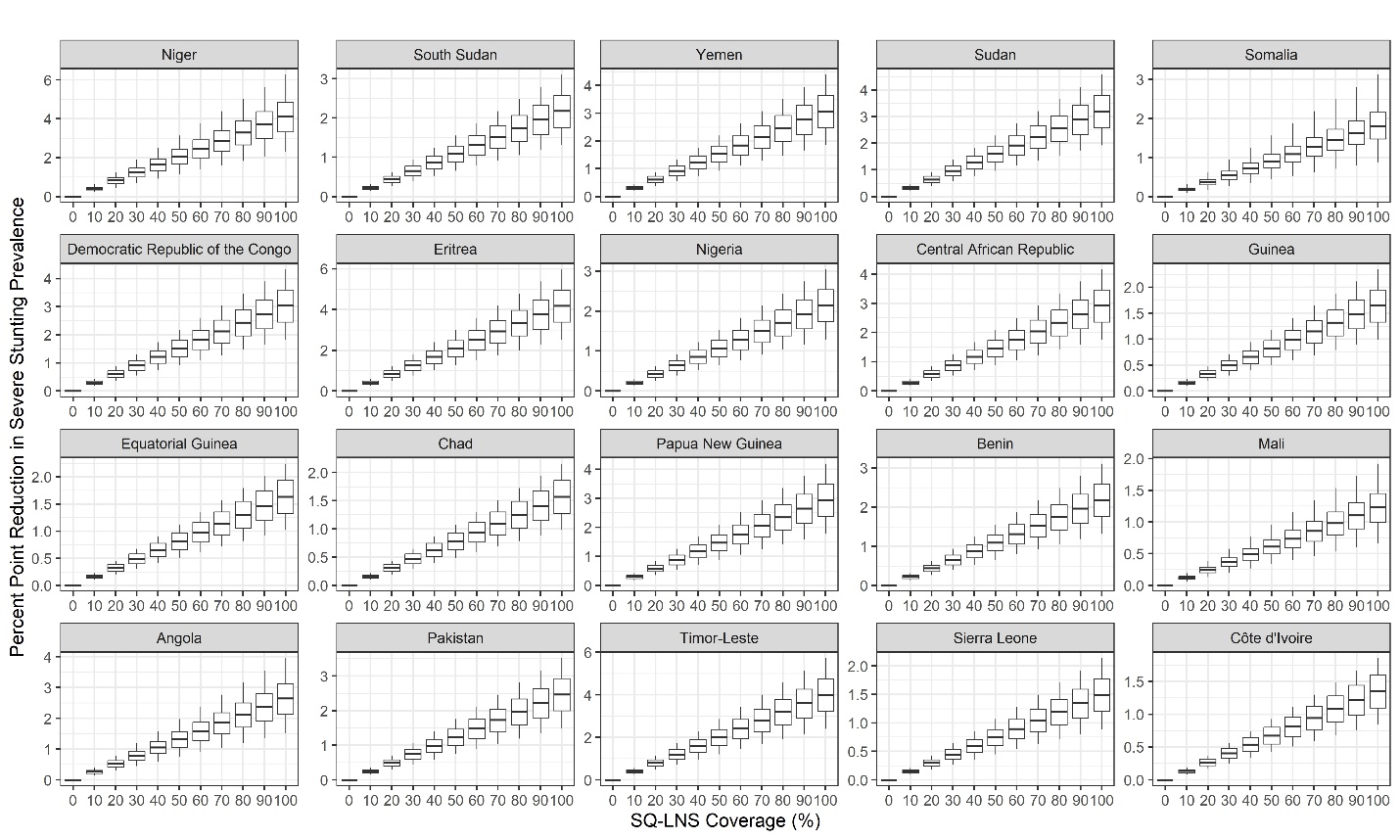

c)

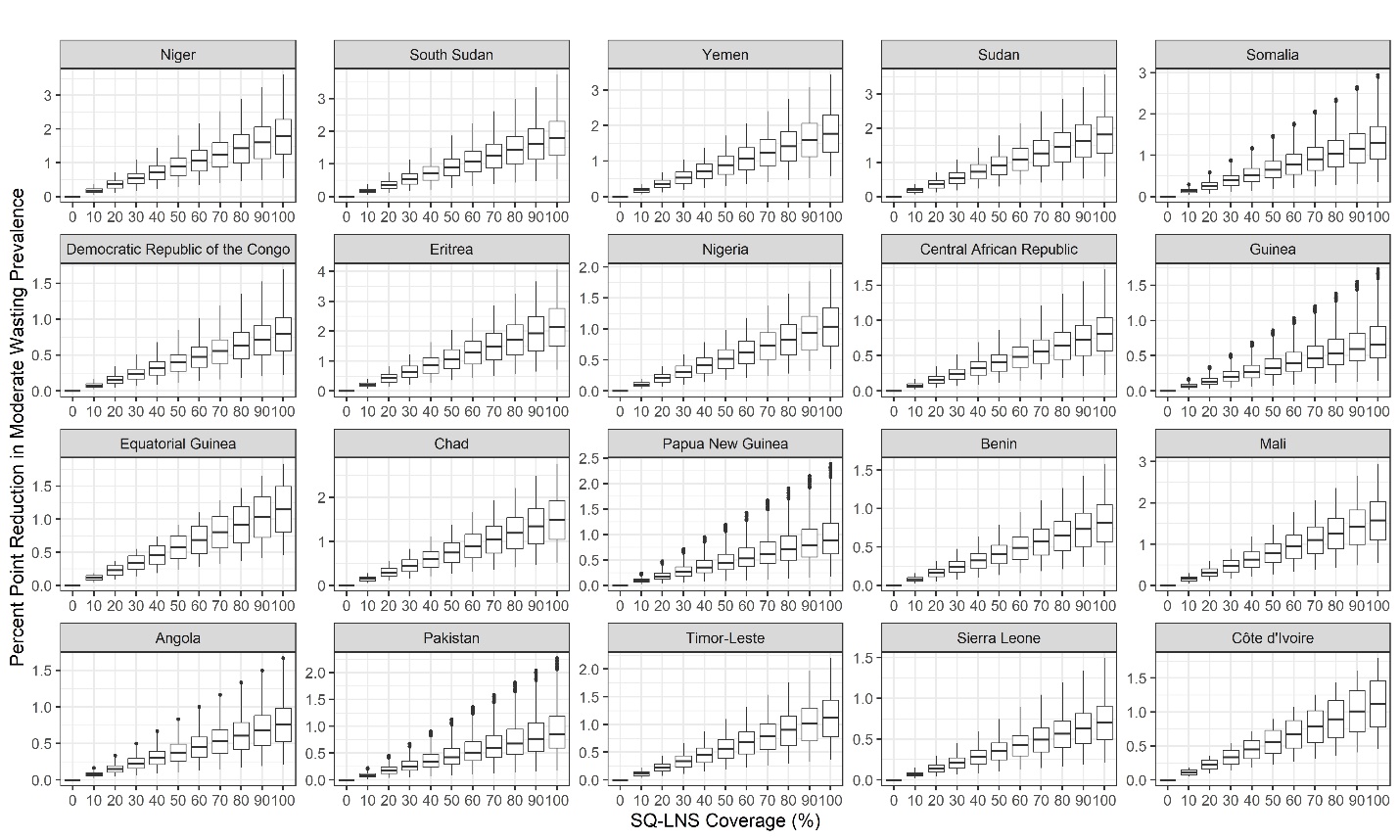

d)

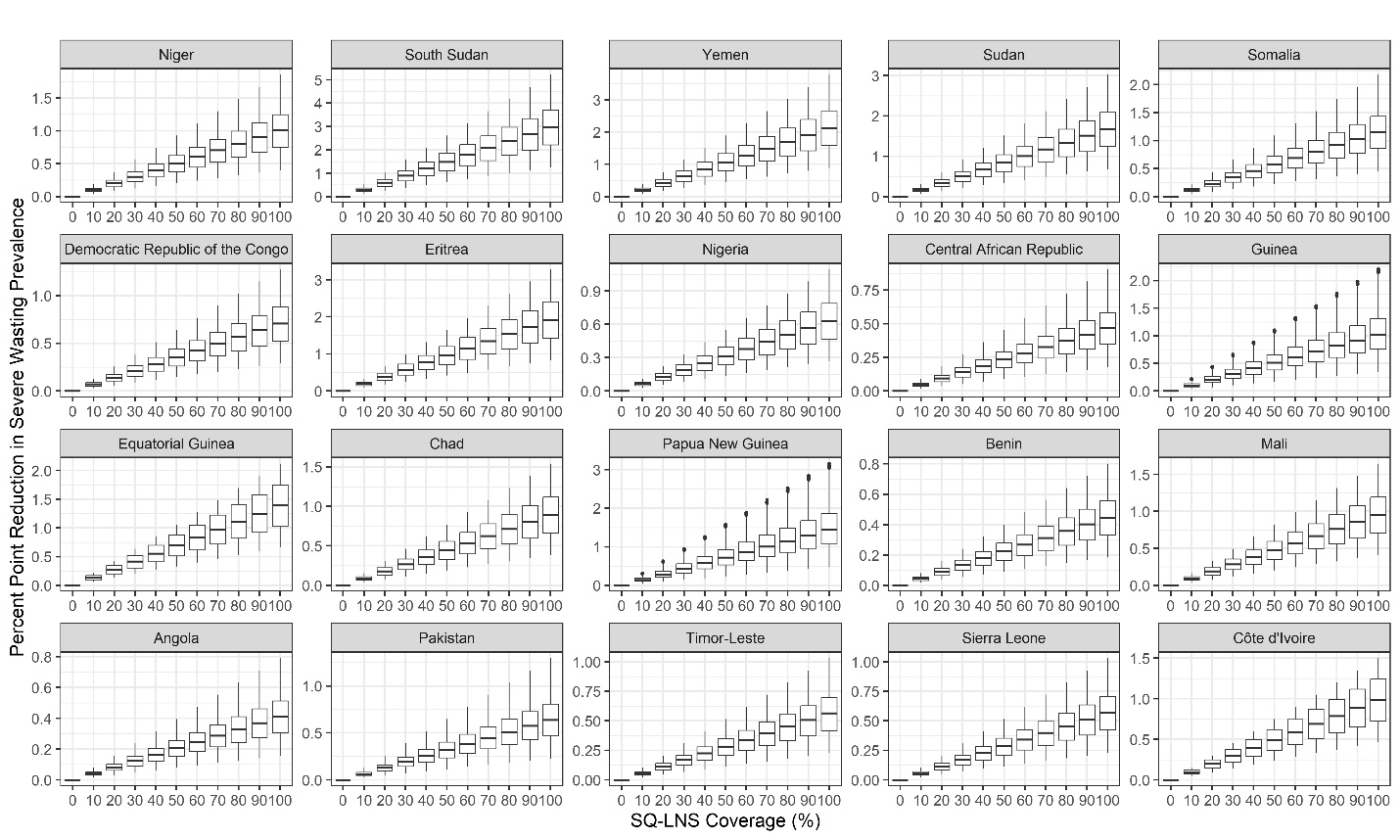

e)

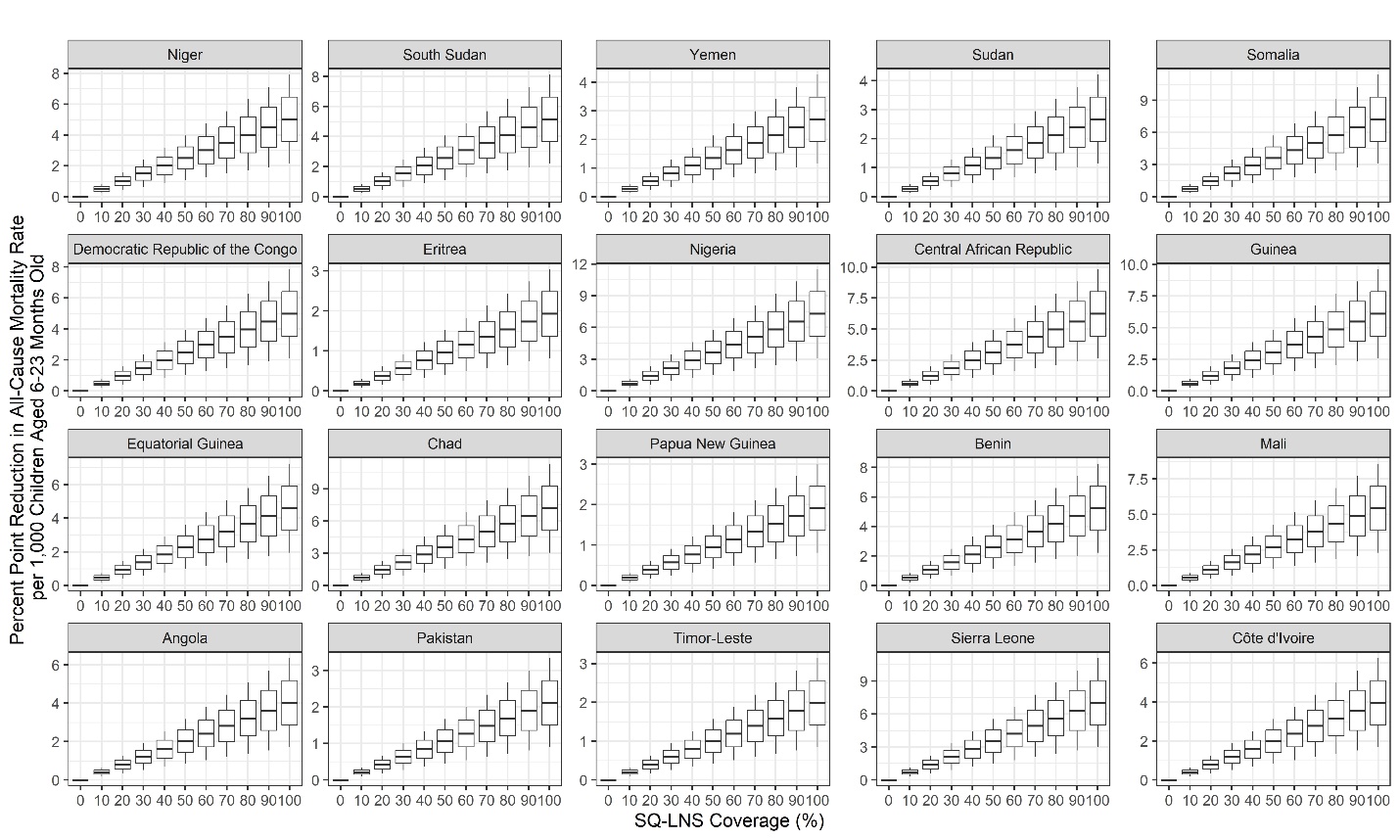

Figure S8. Percentage point reduction in a & b) moderate and severe stunting prevalence, c & d) moderate and severe wasting prevalence and e) all-cause mortality rate per 1,000 live births among children aged 6-23 months old in top 20 countries.

**Section C. Subnational Level Analysis**

The sub-national prevalences estimates of severe wasting among children aged 6 to 23 months old, severe stunting among children aged 24-59 months old, and the all-cause mortality rate among children aged 6 to 23 months per 1,000 live births based on recent survey data in countries listed in Table S1 are given in Figure S9 to S22. We ranked sub-regions within each country for each of these outcomes and averaged the ranks across the three outcomes. Figure S23 to S36 shows the heatmap of the composite score as well as bar graphs of rankings.

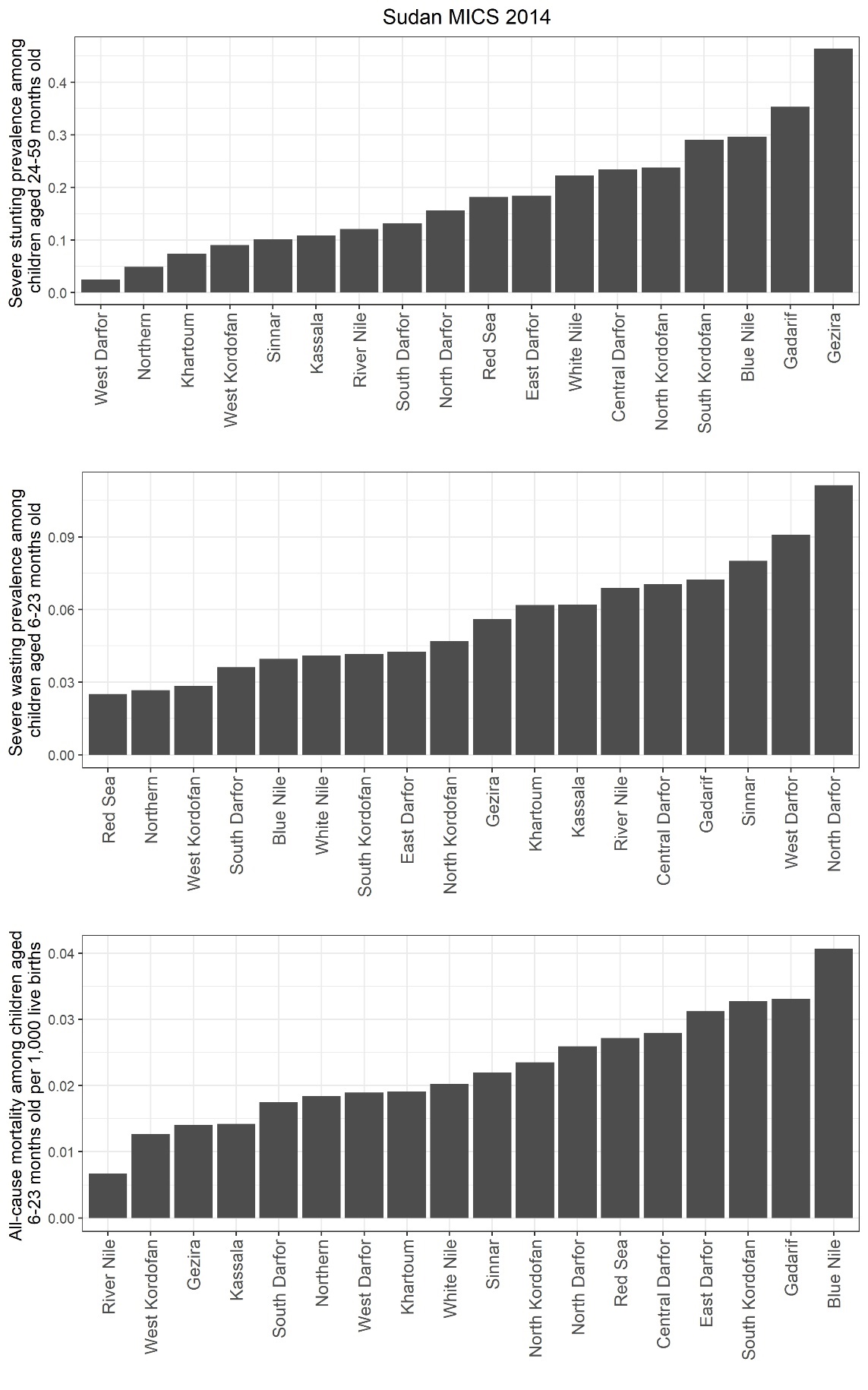

Figure S9. Sub-national prevalence of severe stunting among children aged 24-59 months old, prevalence of severe wasting among children aged 6 to 23 months old, and all-cause mortality among children aged 6 to 23 months per 1,000 live births based on most recent MICS survey in Sudan.

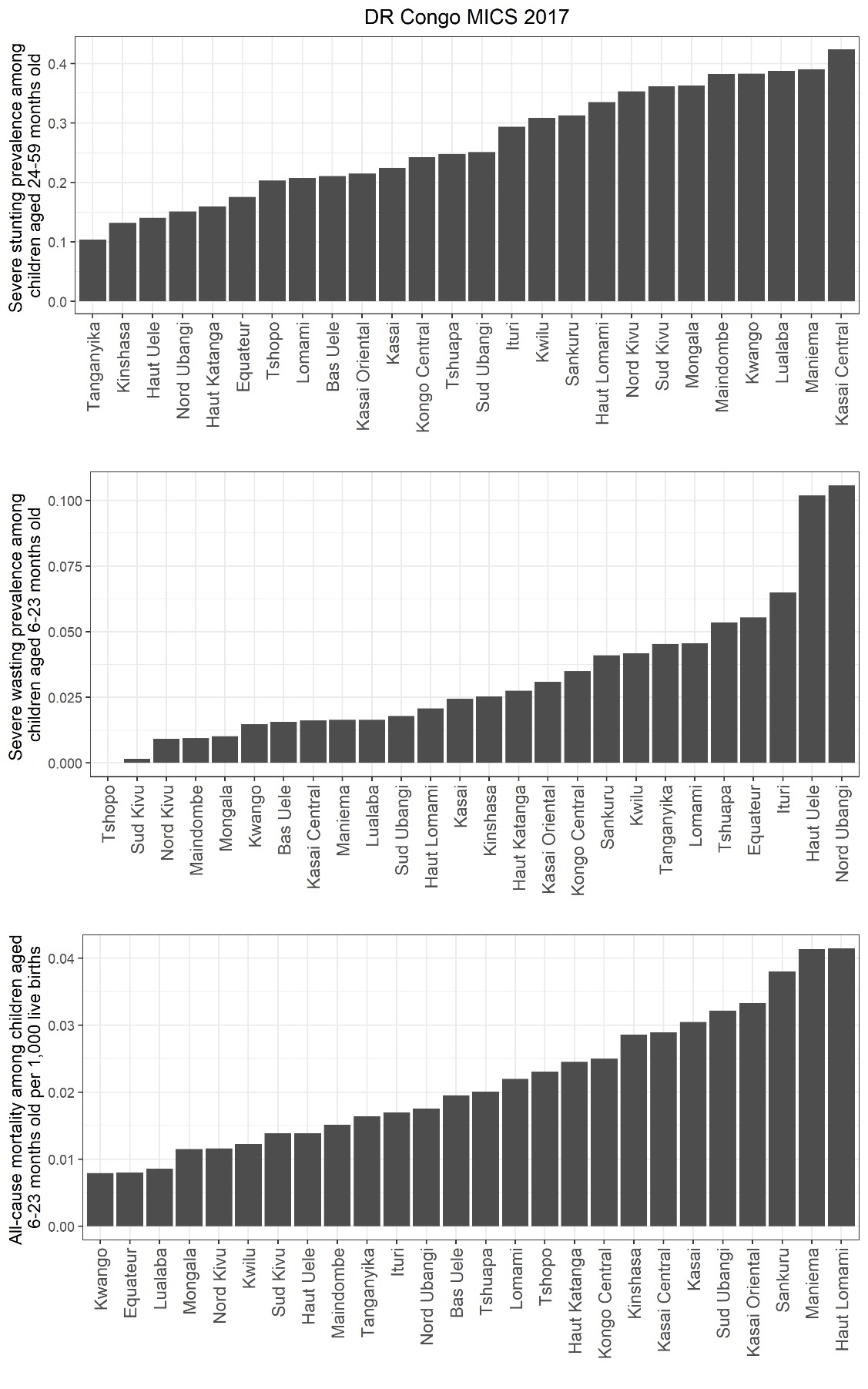

Figure S10. Sub-national prevalence of severe stunting among children aged 24-59 months old, prevalence of severe wasting among children aged 6 to 23 months old, and all-cause mortality among children aged 6 to 23 months per 1,000 live births based on most recent MICS survey in Democratic Republic of Congo.

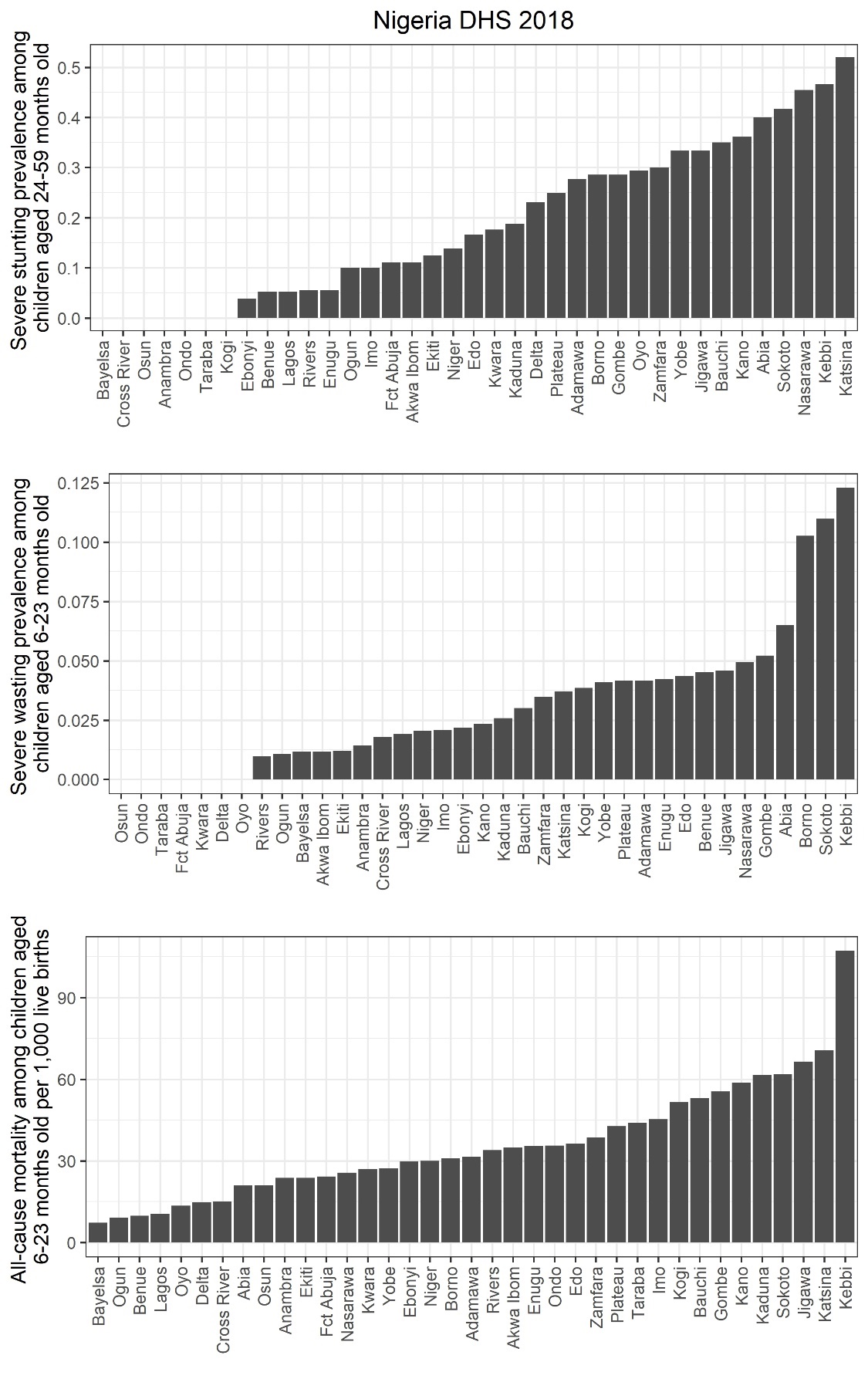

Figure S11. Sub-national prevalence of severe stunting among children aged 24-59 months old, prevalence of severe wasting among children aged 6 to 23 months old, and all-cause mortality among children aged 6 to 23 months per 1,000 live births based on the most recent DHS survey in Nigeria.

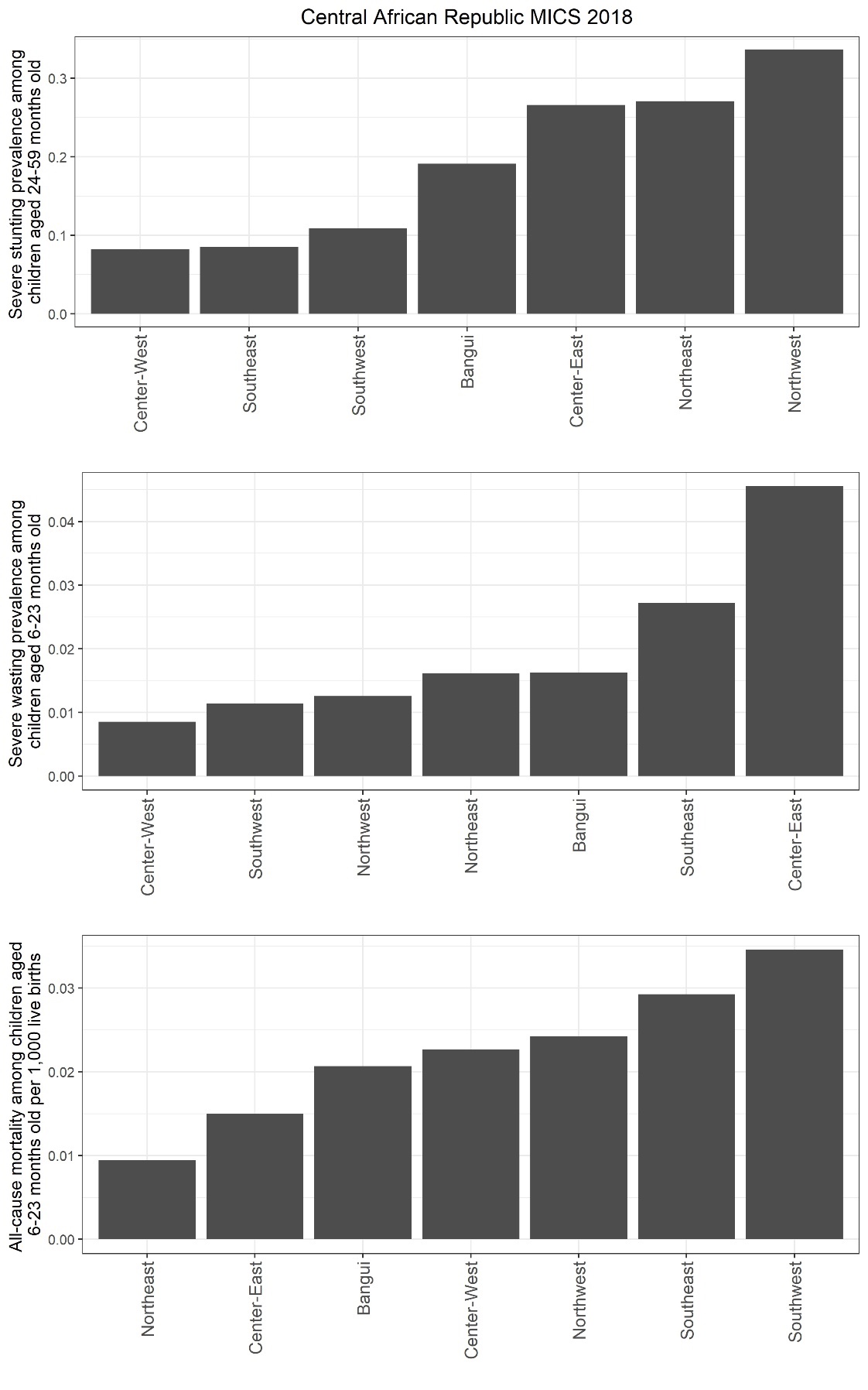

Figure S12. Sub-national prevalence of severe stunting among children aged 24-59 months old, prevalence of severe wasting among children aged 6 to 23 months old, and all-cause mortality among children aged 6 to 23 months per 1,000 live births based on most recent MICS survey in Central African Republic.

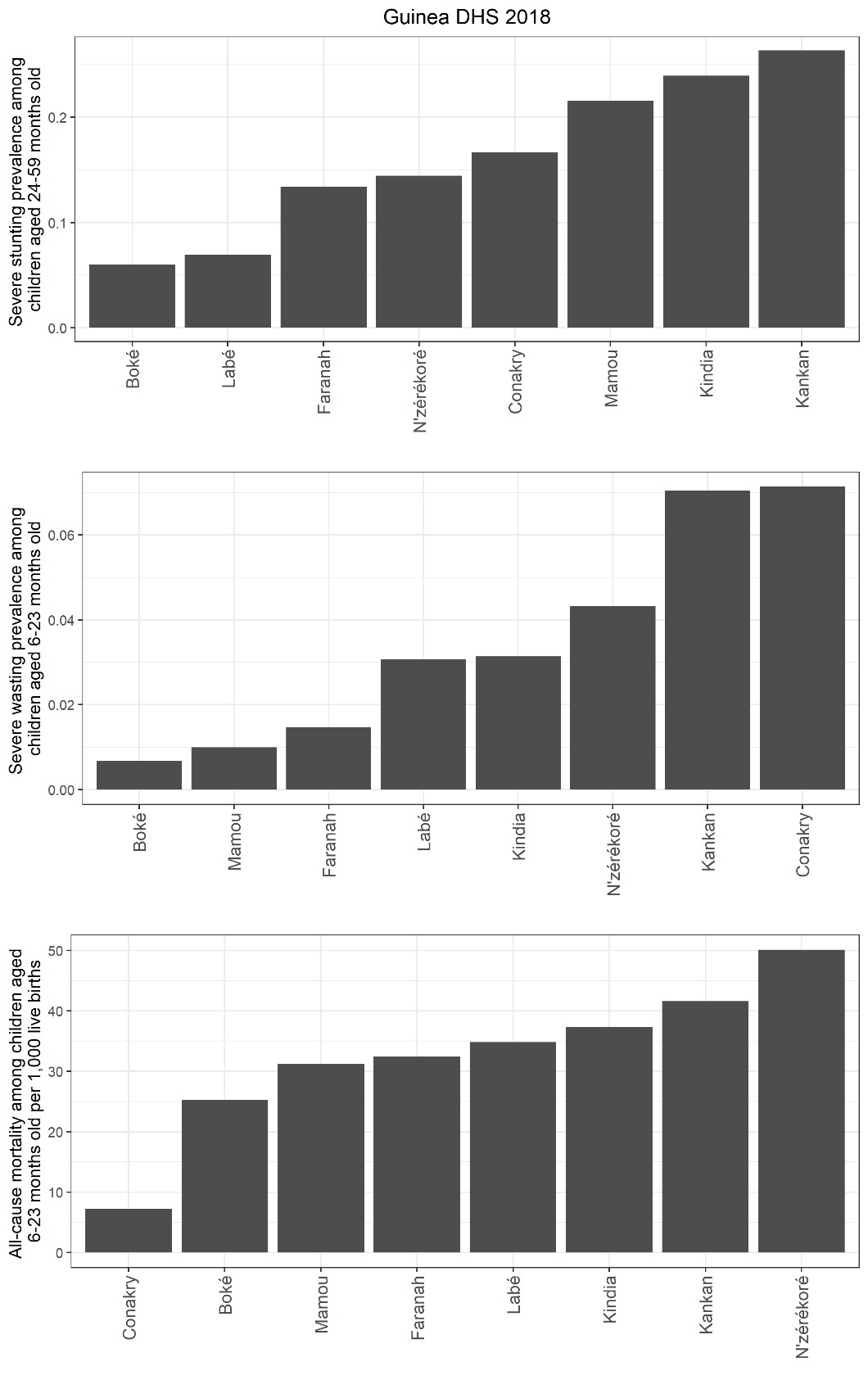

Figure S13. Sub-national prevalence of severe stunting among children aged 24-59 months old, prevalence of severe wasting among children aged 6 to 23 months old, and all-cause mortality among children aged 6 to 23 months per 1,000 live births based on the most recent DHS survey in Guinea.

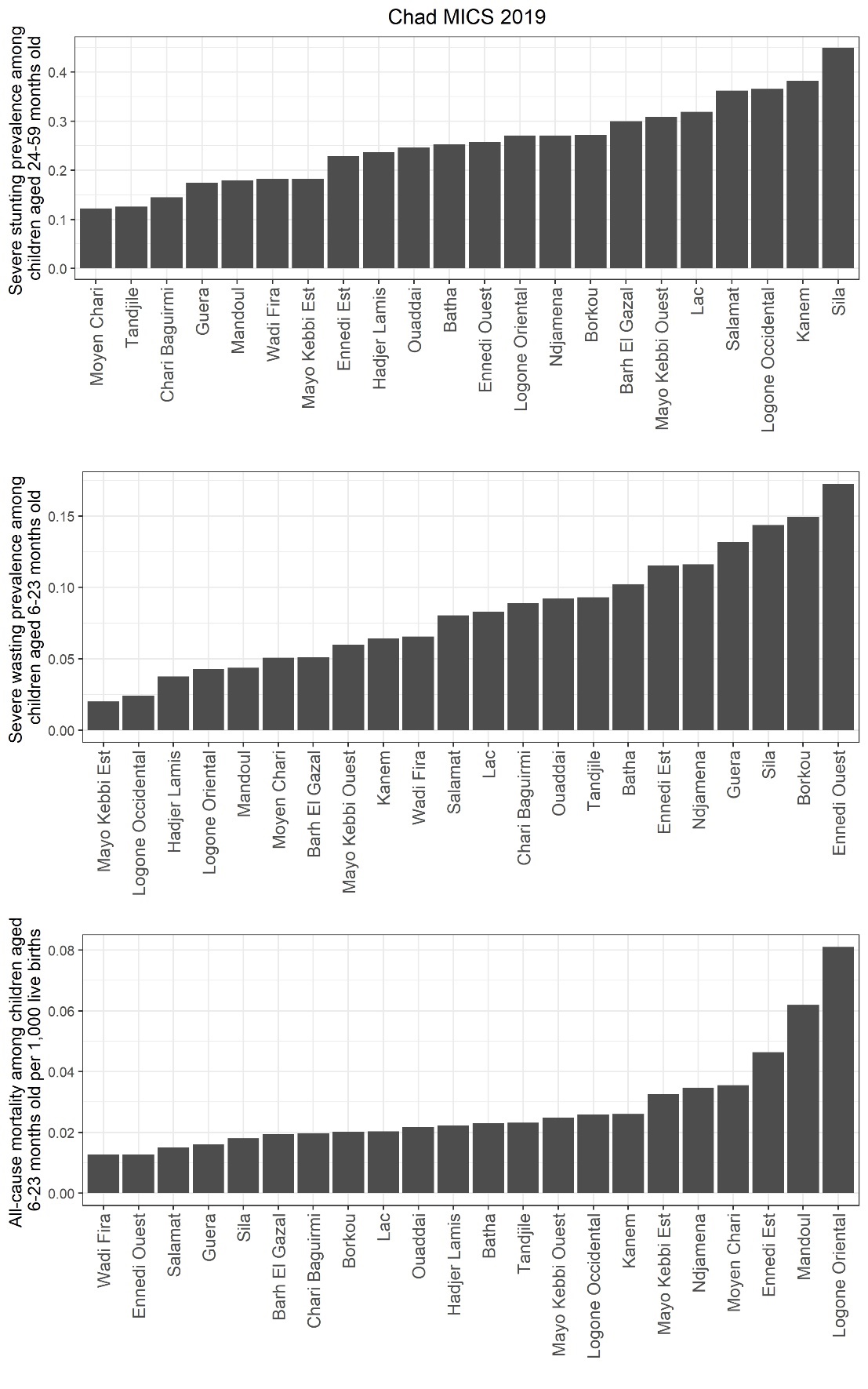

Figure S14. Sub-national prevalence of severe stunting among children aged 24-59 months old, prevalence of severe wasting among children aged 6 to 23 months old, and all-cause mortality among children aged 6 to 23 months per 1,000 live births based on most recent MICS survey in Chad.

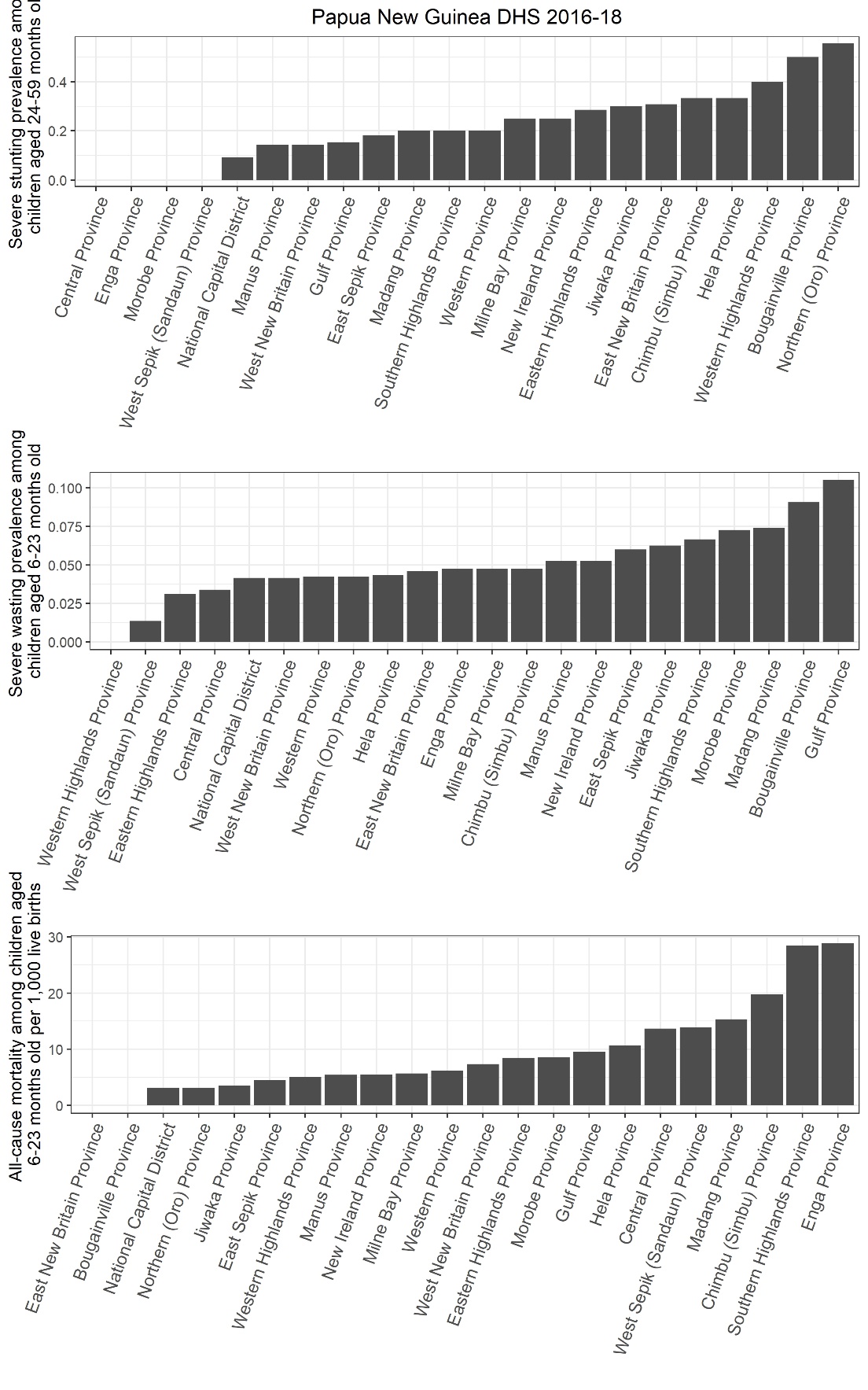

Figure S15. Sub-national prevalence of severe stunting among children aged 24-59 months old, prevalence of severe wasting among children aged 6 to 23 months old, and all-cause mortality among children aged 6 to 23 months per 1,000 live births based on the most recent DHS survey in Papua New Guinea.

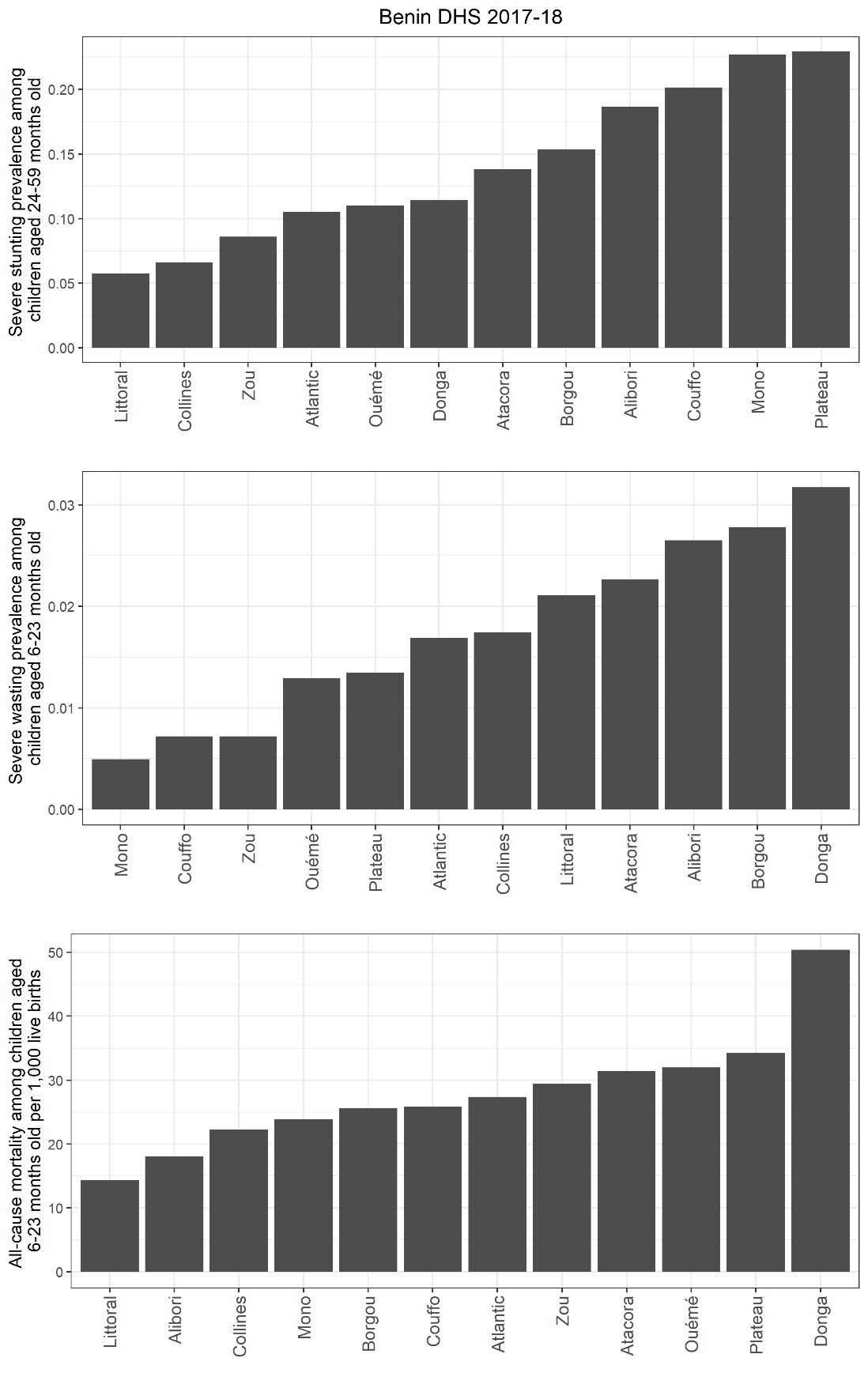

Figure S16. Sub-national prevalence of severe stunting among children aged 24-59 months old, prevalence of severe wasting among children aged 6 to 23 months old, and all-cause mortality among children aged 6 to 23 months per 1,000 live births based on the most recent DHS survey in Benin.

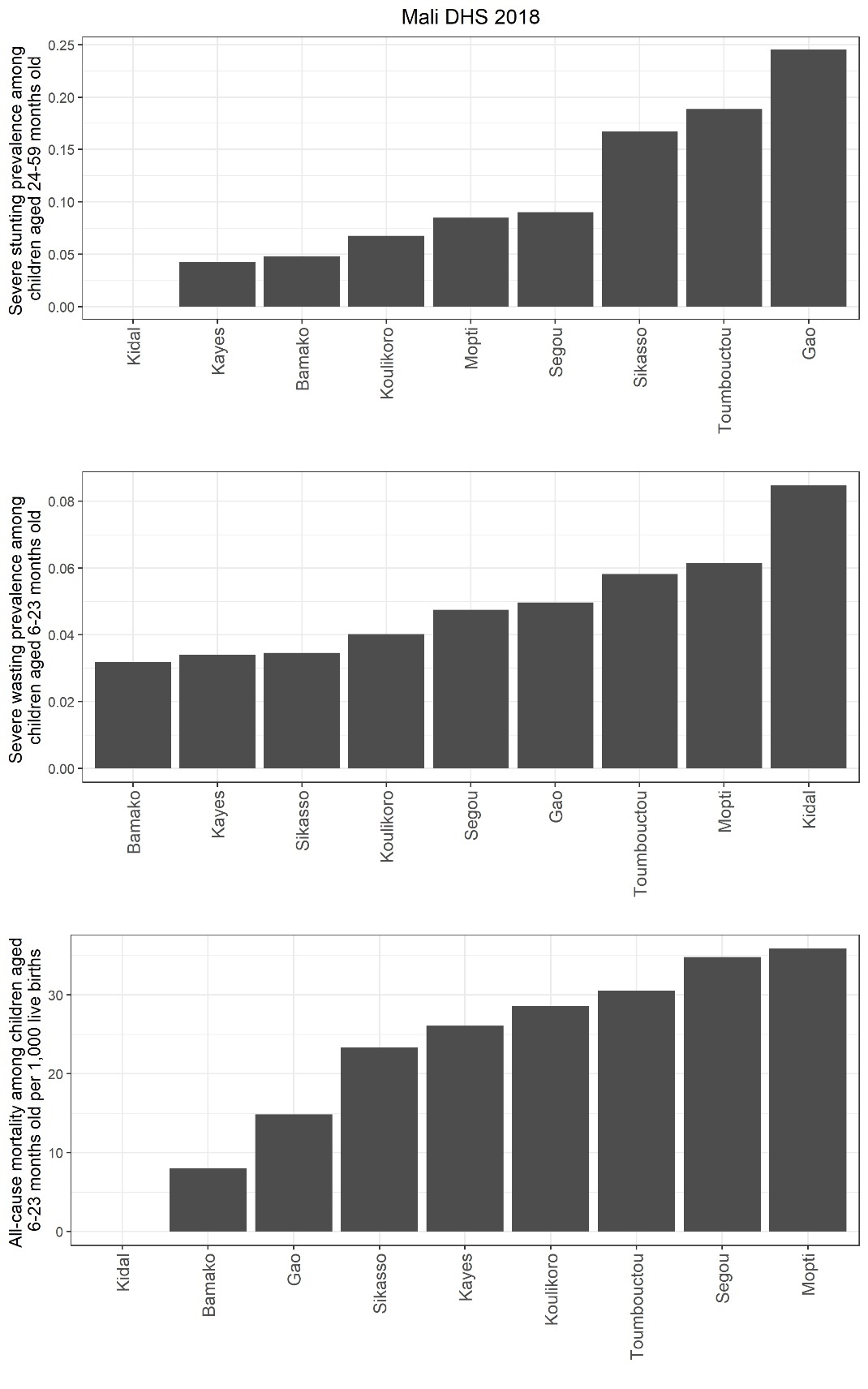

Figure S17. Sub-national prevalence of severe stunting among children aged 24-59 months old, prevalence of severe wasting among children aged 6 to 23 months old, and all-cause mortality among children aged 6 to 23 months per 1,000 live births based on the most recent DHS survey in Mali.

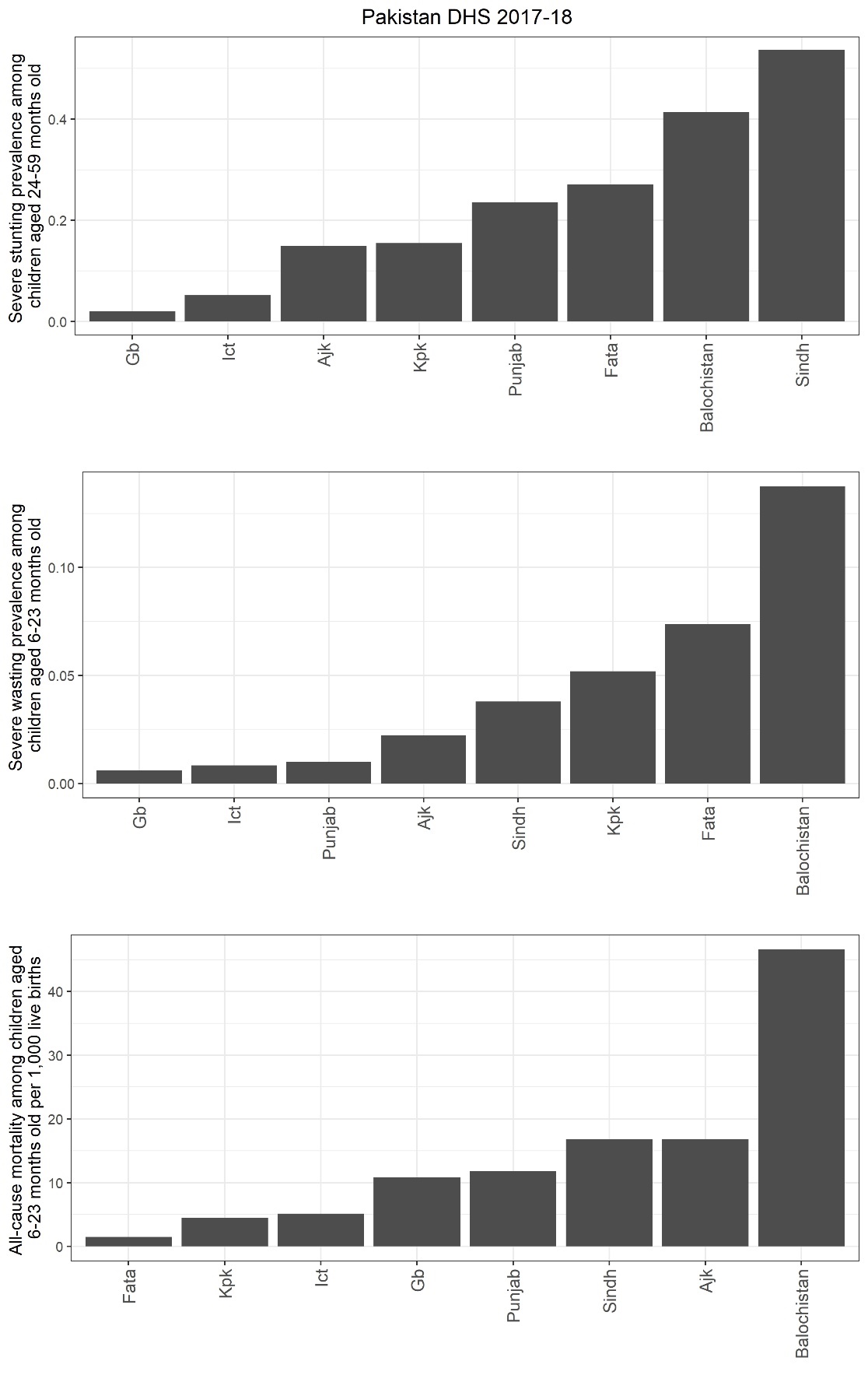

Figure S18. Sub-national prevalence of severe stunting among children aged 24-59 months old, prevalence of severe wasting among children aged 6 to 23 months old, and all-cause mortality among children aged 6 to 23 months per 1,000 live births based on the most recent DHS survey in Pakistan.

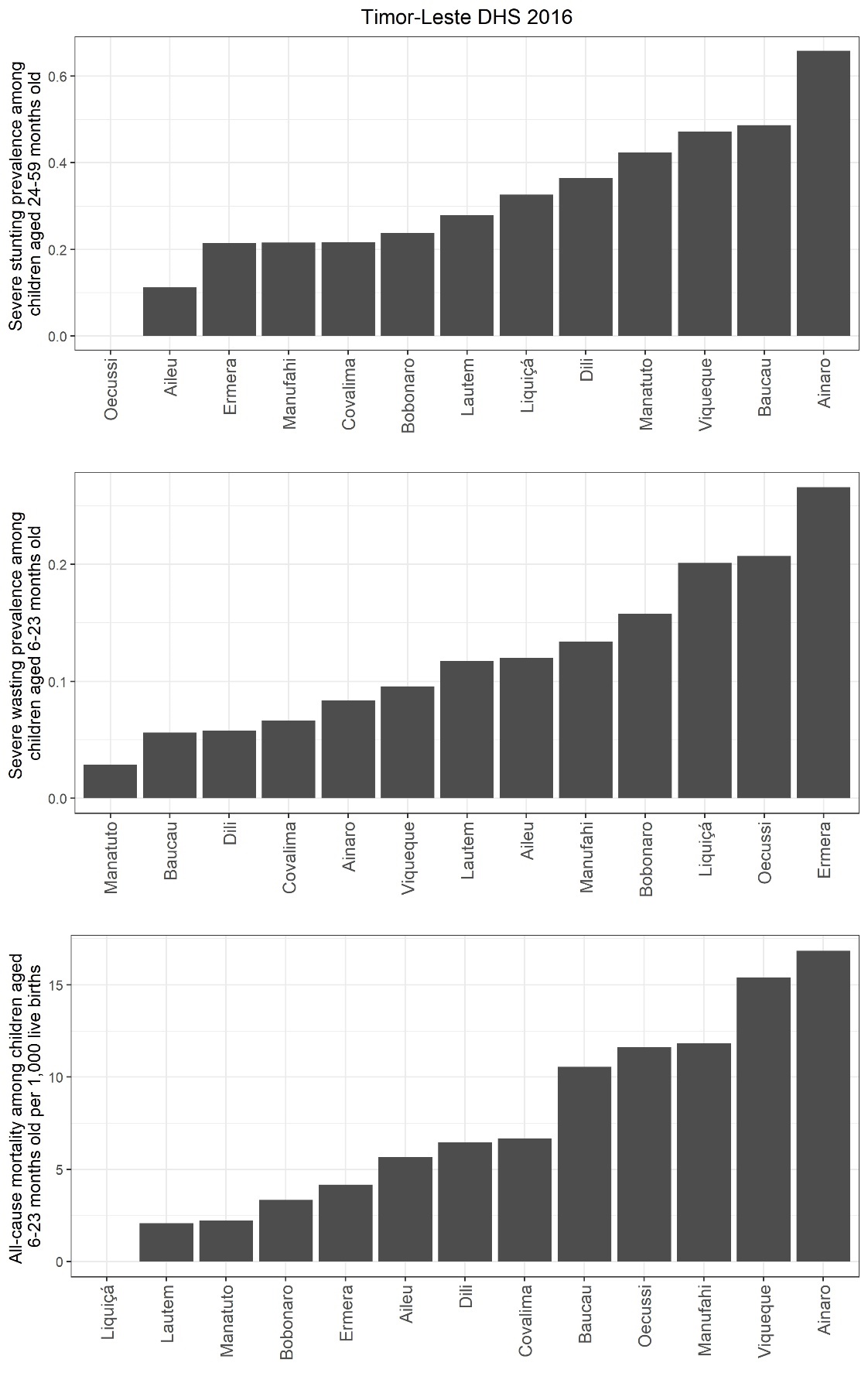

Figure S19. Sub-national prevalence of severe stunting among children aged 24-59 months old, prevalence of severe wasting among children aged 6 to 23 months old, and all-cause mortality among children aged 6 to 23 months per 1,000 live births based on the most recent DHS survey in Timor-Leste.

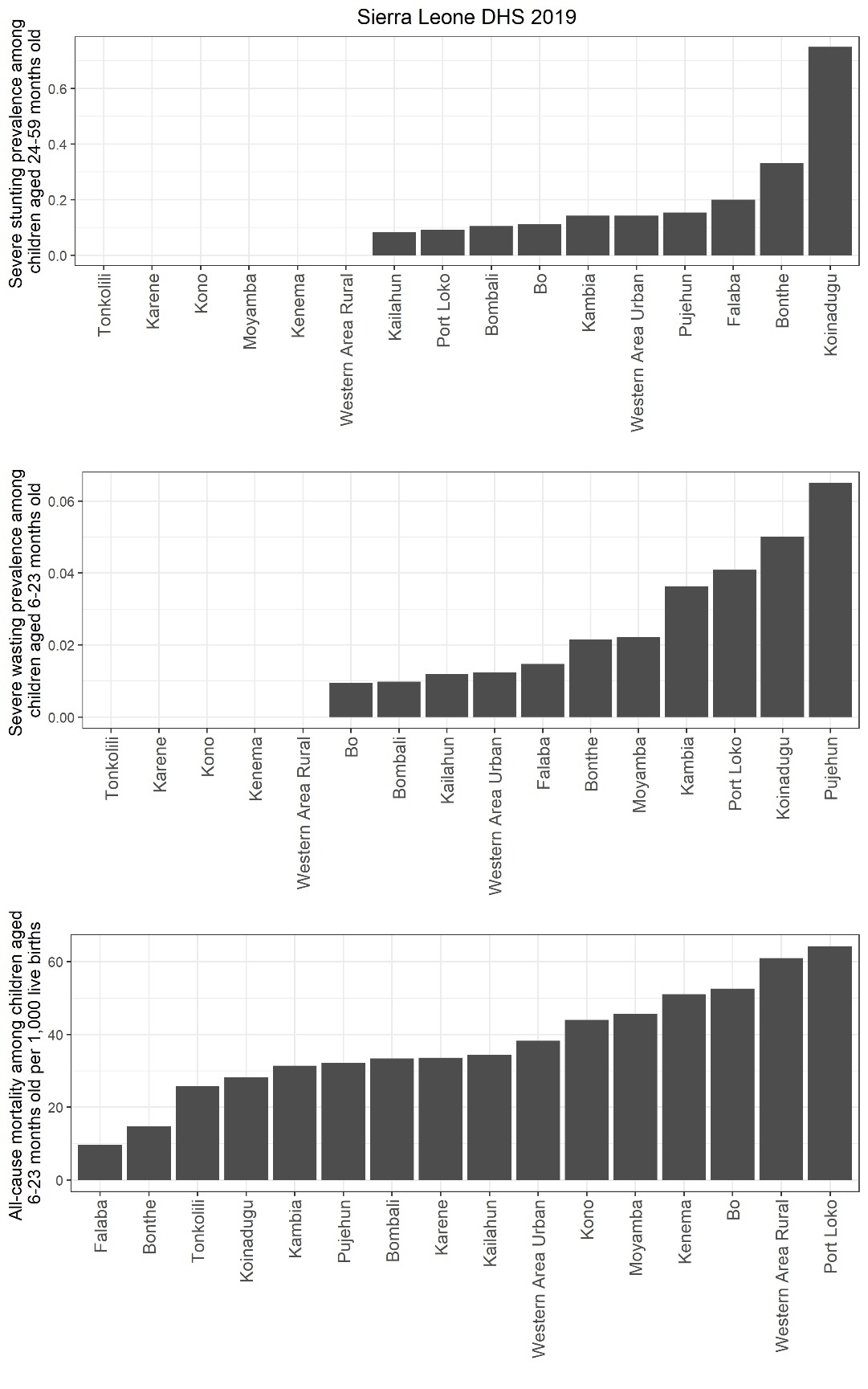

Figure S20. Sub-national prevalence of severe stunting among children aged 24-59 months old, prevalence of severe wasting among children aged 6 to 23 months old, and all-cause mortality among children aged 6 to 23 months per 1,000 live births based on the most recent DHS survey in Sierra Leone.

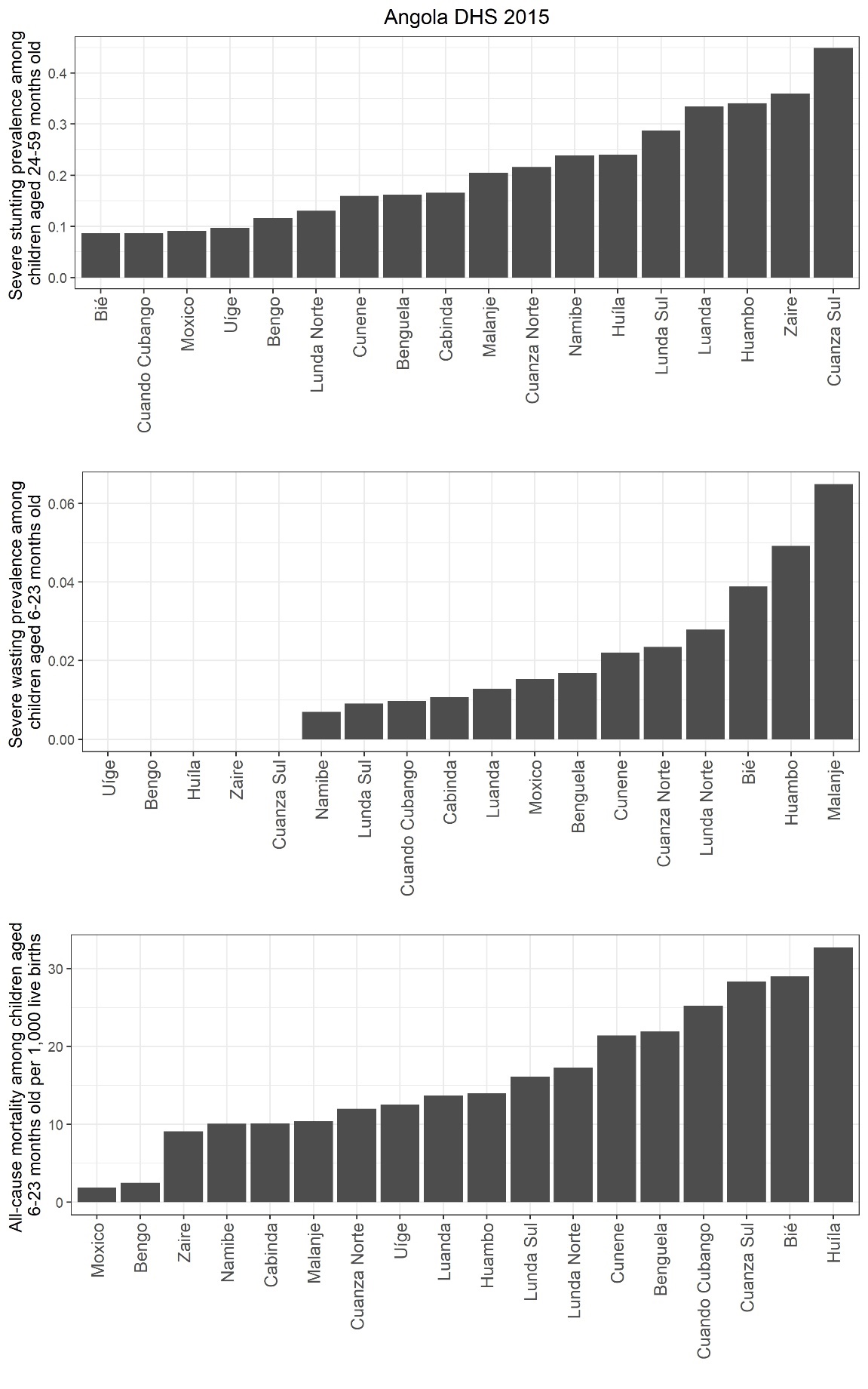

Figure S21. Sub-national prevalence of severe stunting among children aged 24-59 months old, prevalence of severe wasting among children aged 6 to 23 months old, and all-cause mortality among children aged 6 to 23 months per 1,000 live births based on the most recent DHS survey in Angola.

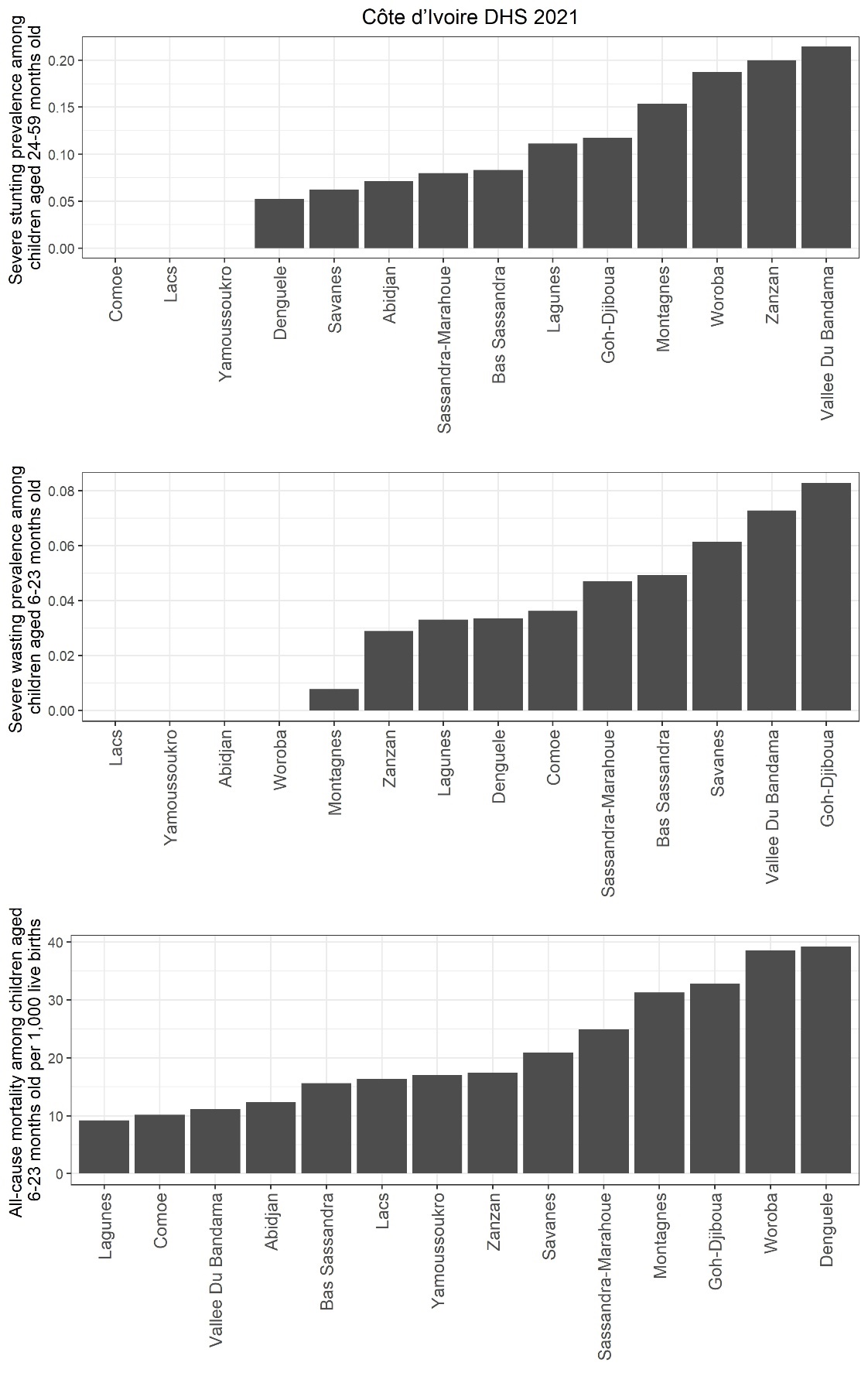

Figure S22. Sub-national prevalence of severe stunting among children aged 24-59 months old, prevalence of severe wasting among children aged 6 to 23 months old, and all-cause mortality among children aged 6 to 23 months per 1,000 live births based on the most recent DHS survey in Côte d’Ivoire.

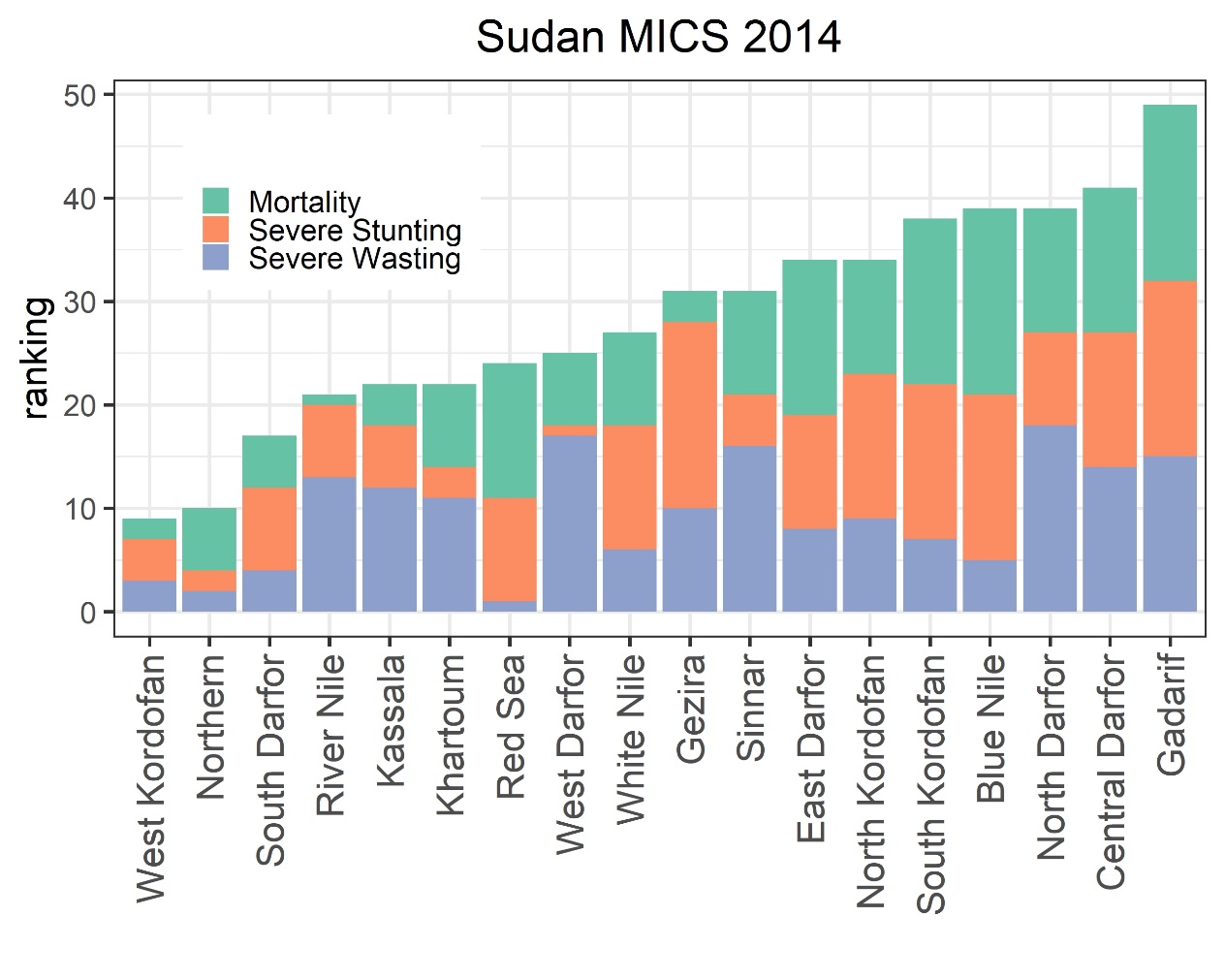

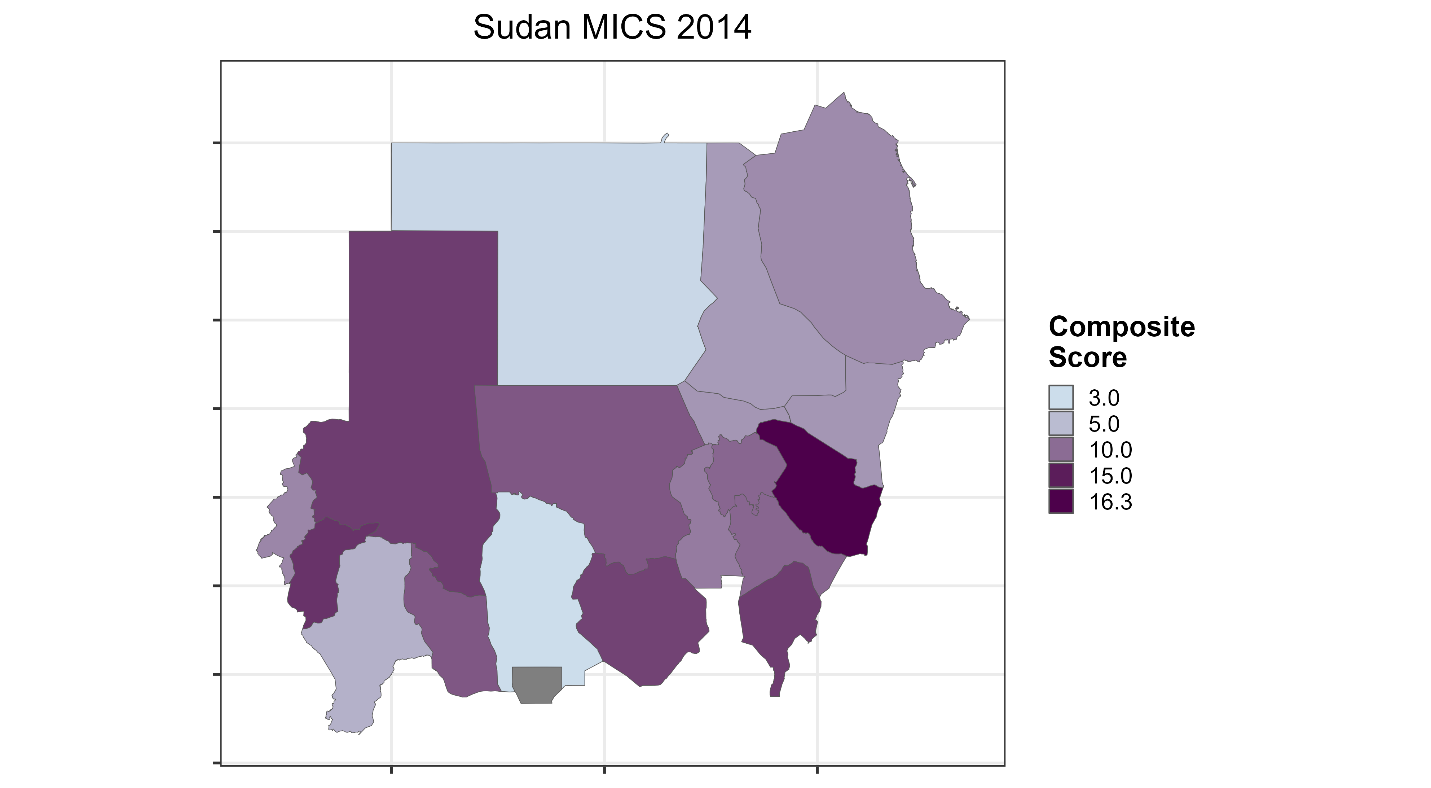

Figure S23. Sub-national ranking of Sudan by highest mortality rate and highest prevalence of severe stunting and severe wasting as well as heatmap of composite score at the sub-national level

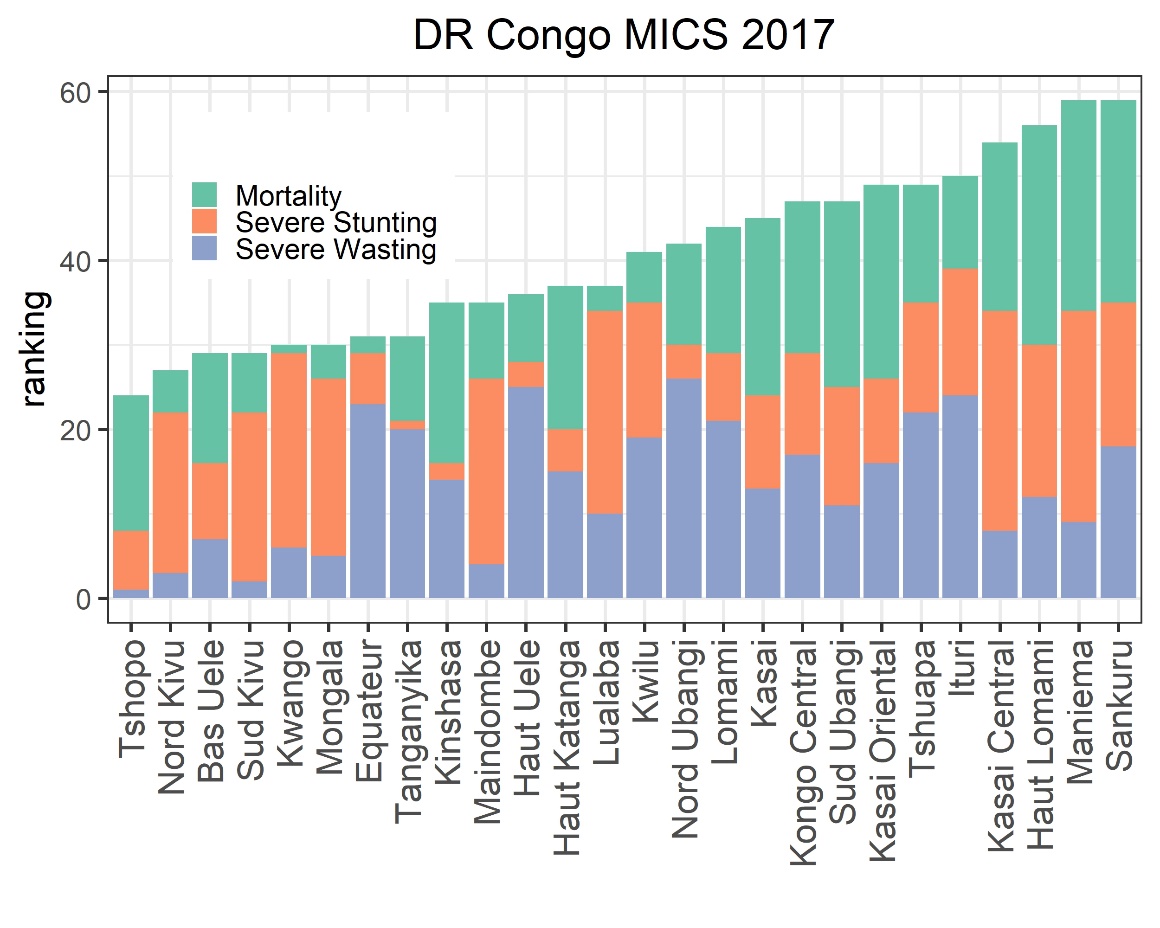

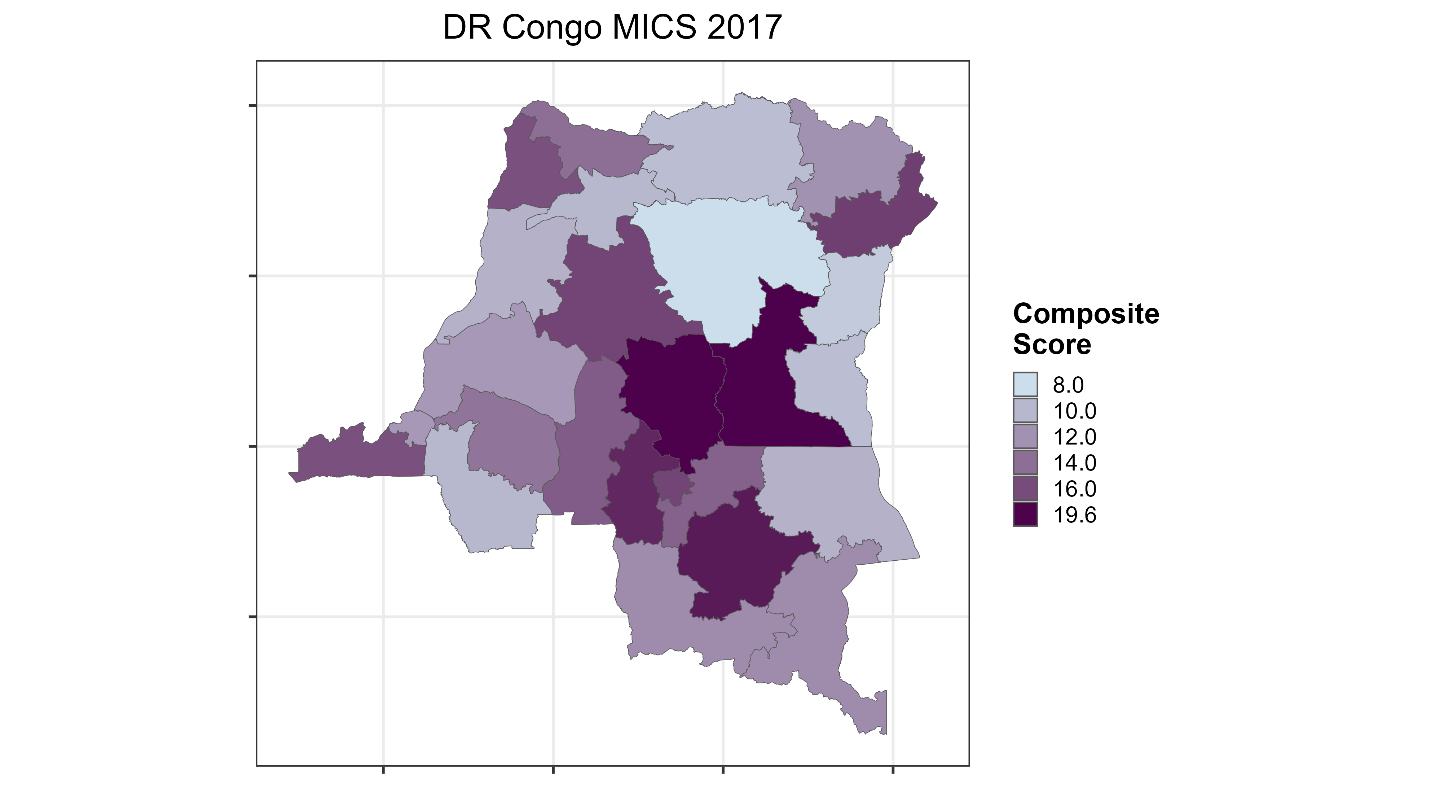

Figure S24. Sub-national ranking of Democratic Republic of Congo by highest mortality rate and highest prevalence of severe stunting and severe wasting as well as heatmap of composite score at the sub-national level

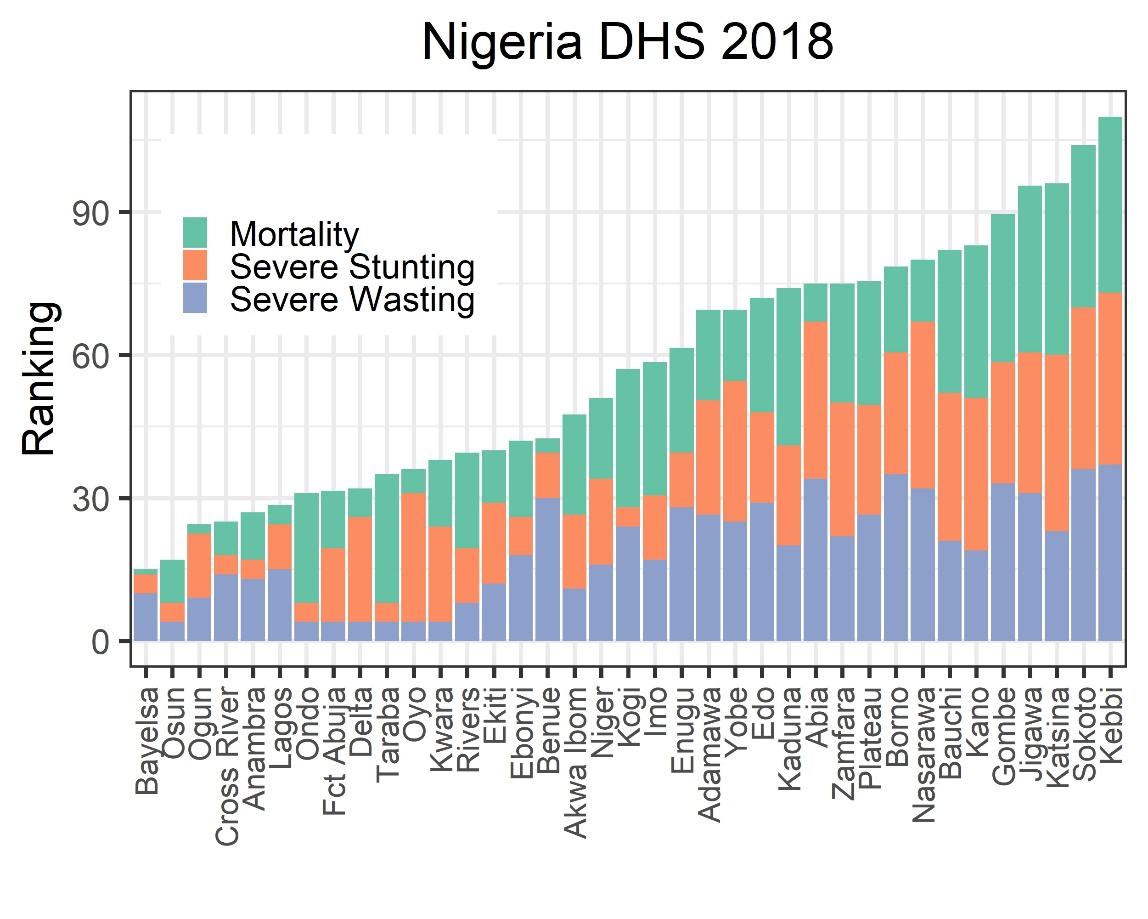

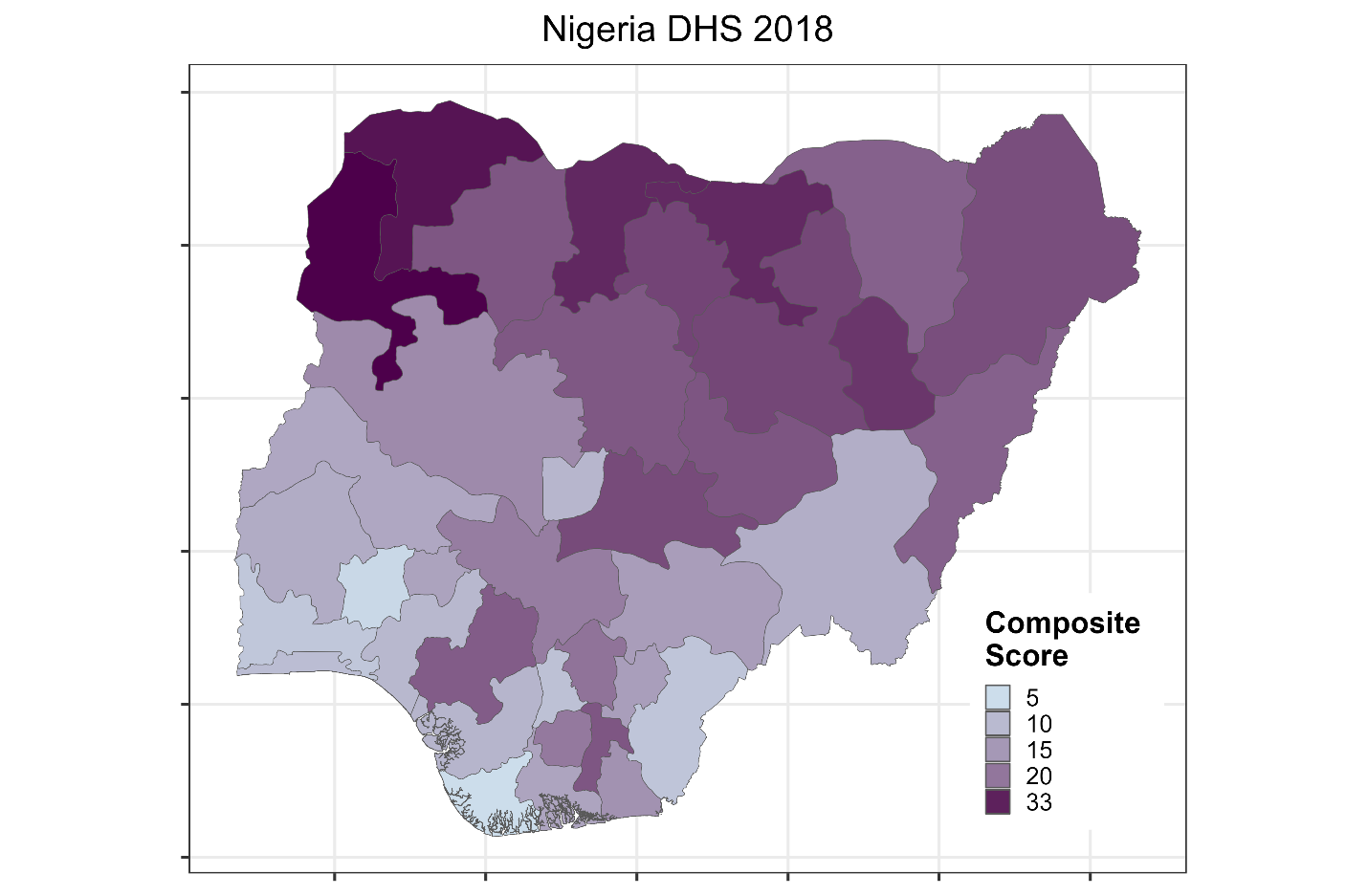

Figure S25. Sub-national ranking of Nigeria by highest mortality rate and highest prevalence of severe stunting and severe wasting as well as heatmap of composite score at the sub-national level

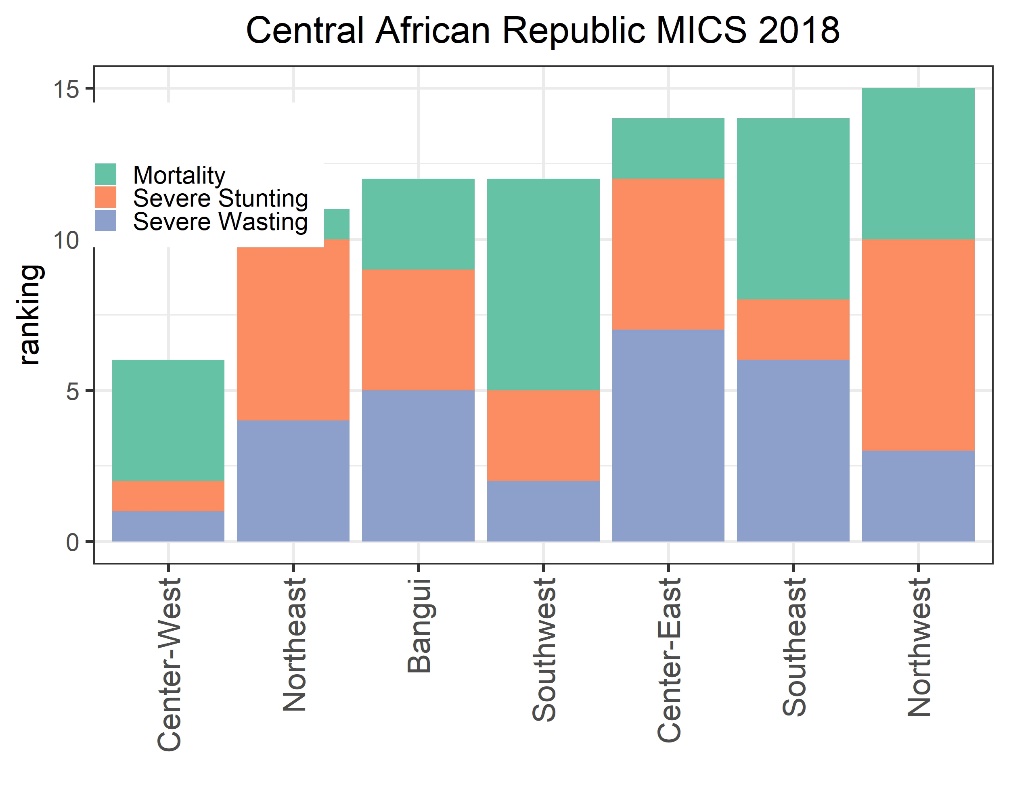

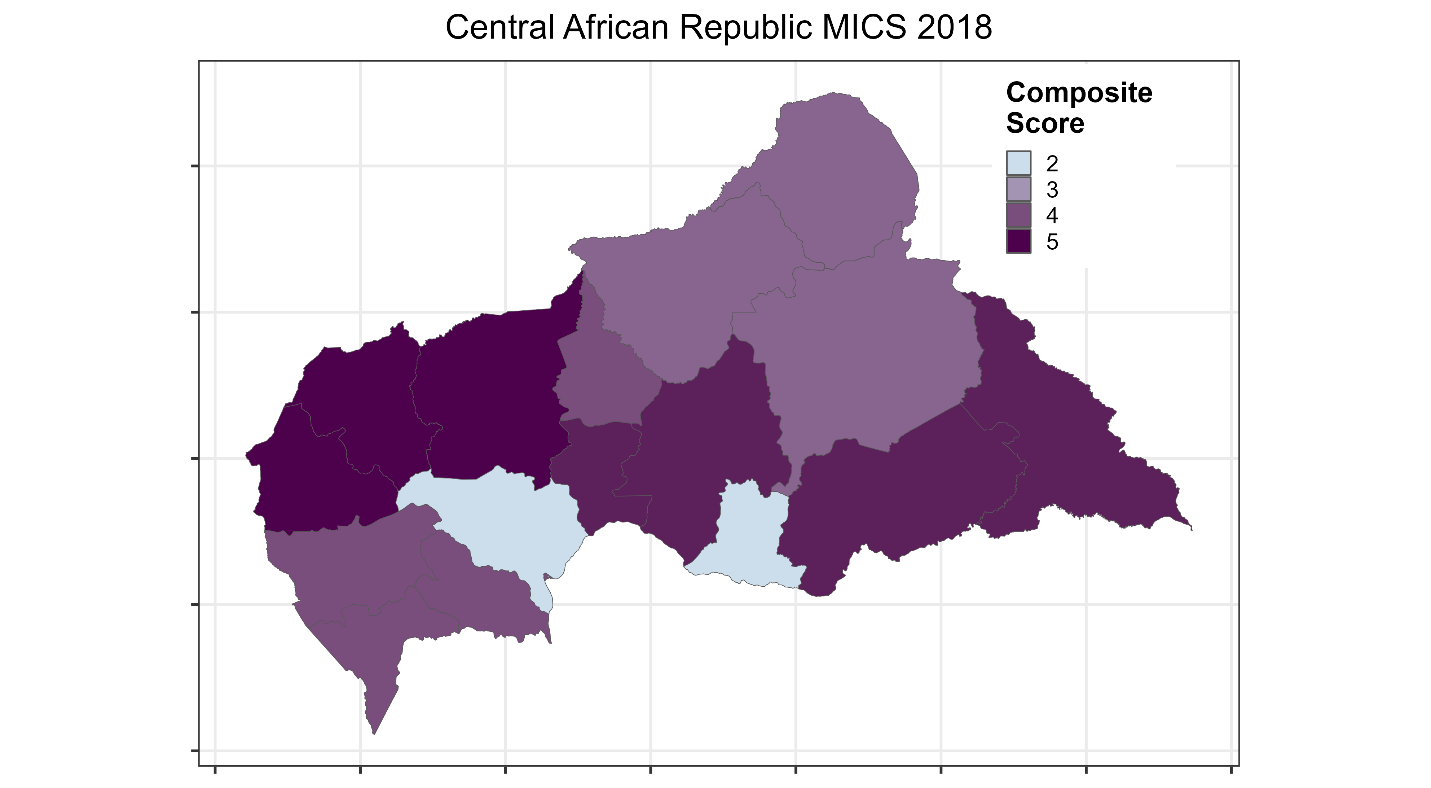

Figure S26. Sub-national ranking of Central African Republic by highest mortality rate and highest prevalence of severe stunting and severe wasting as well as heatmap of composite score at the sub-national level

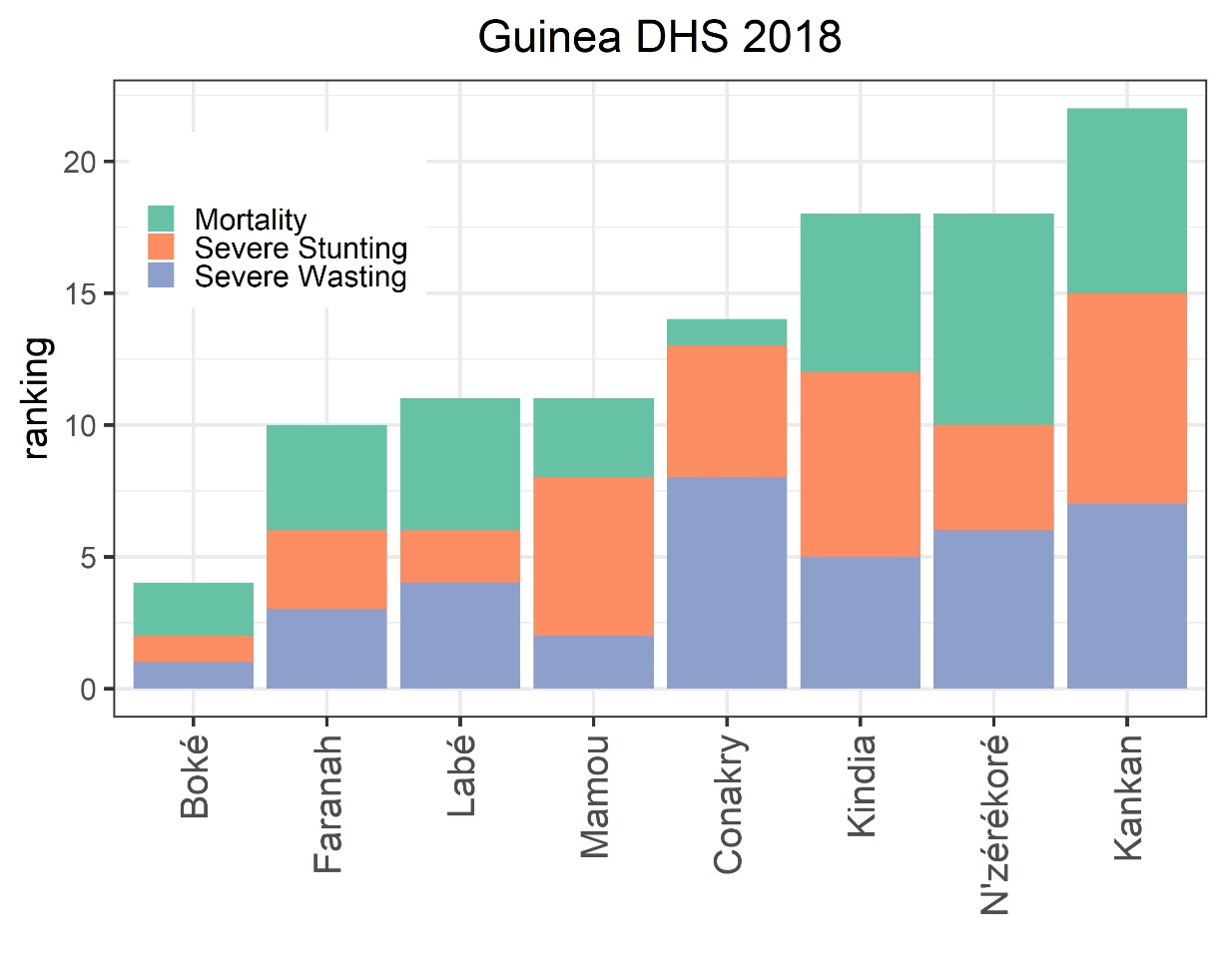

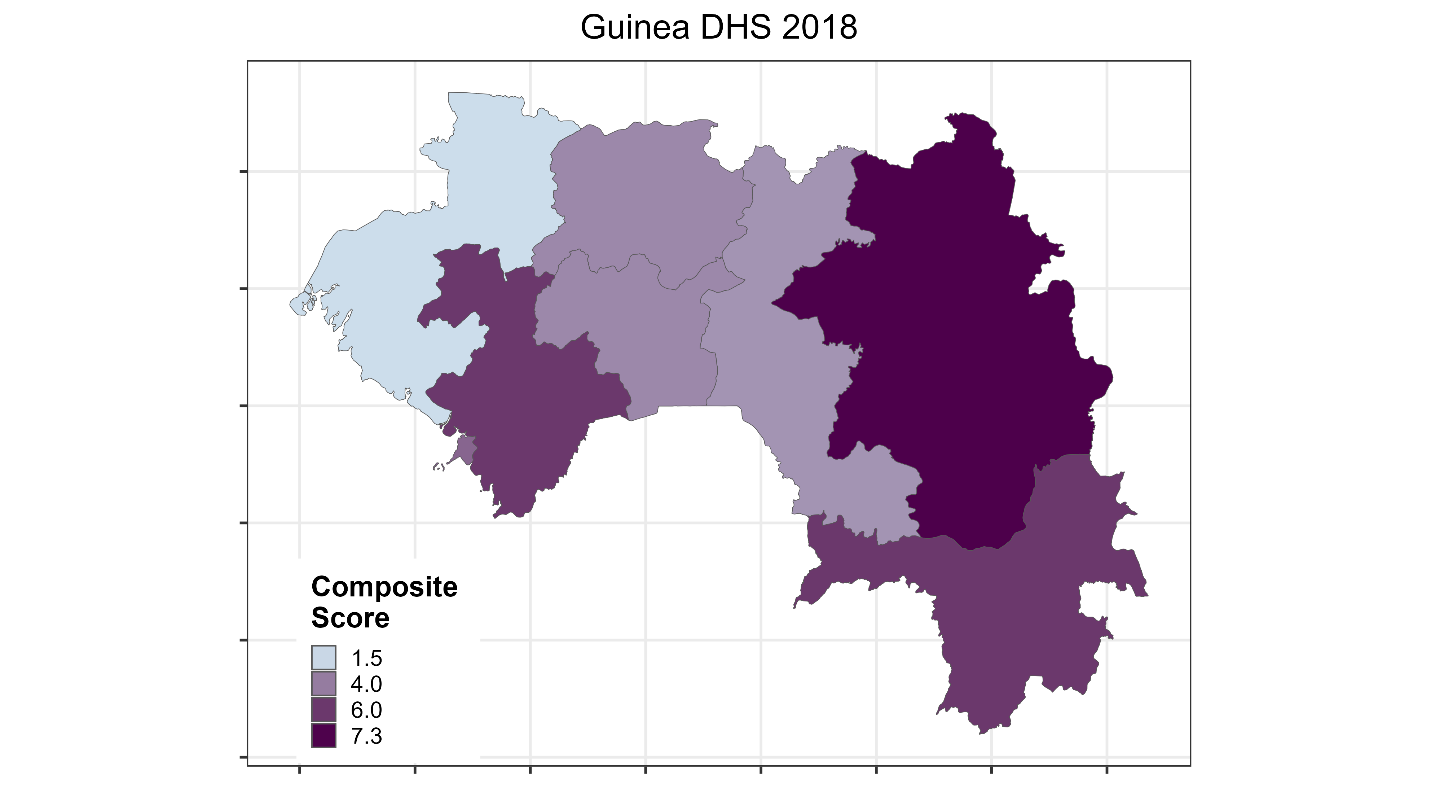

Figure S27. Sub-national ranking of Guinea by highest mortality rate and highest prevalence of severe stunting and severe wasting as well as heatmap of composite score at the sub-national level

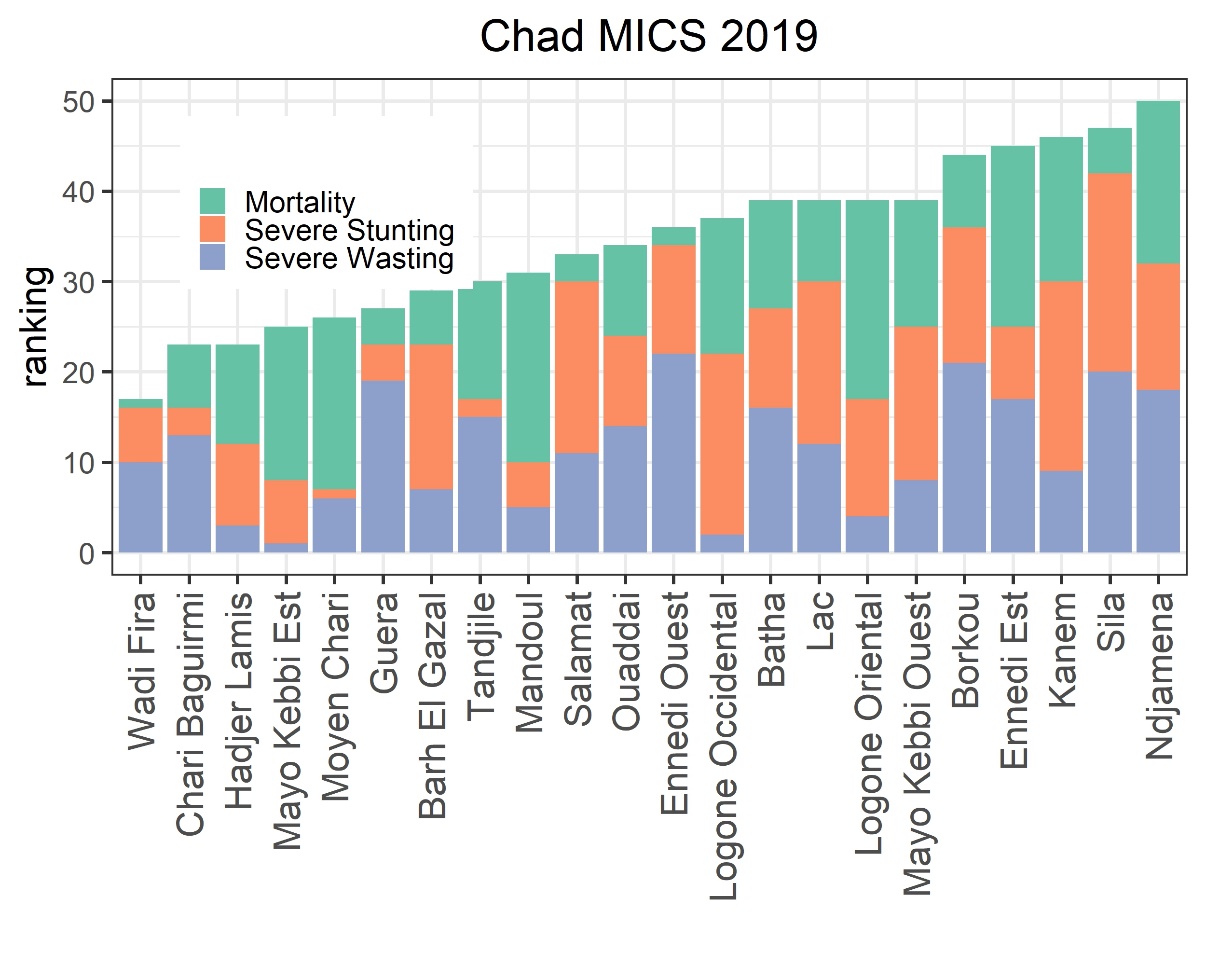

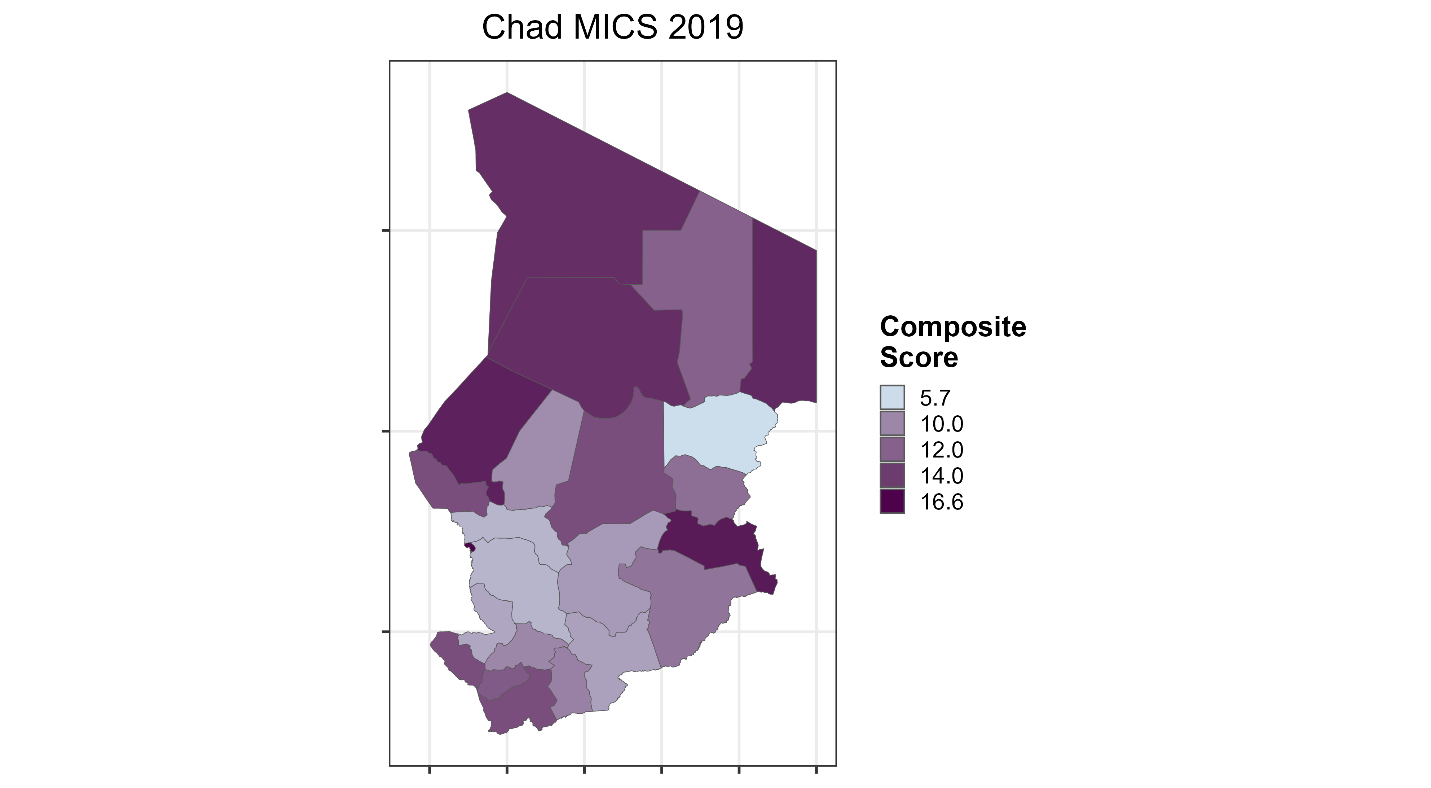

Figure S28. Sub-national ranking of Chad by highest mortality rate and highest prevalence of severe stunting and severe wasting as well as heatmap of composite score at the sub-national level

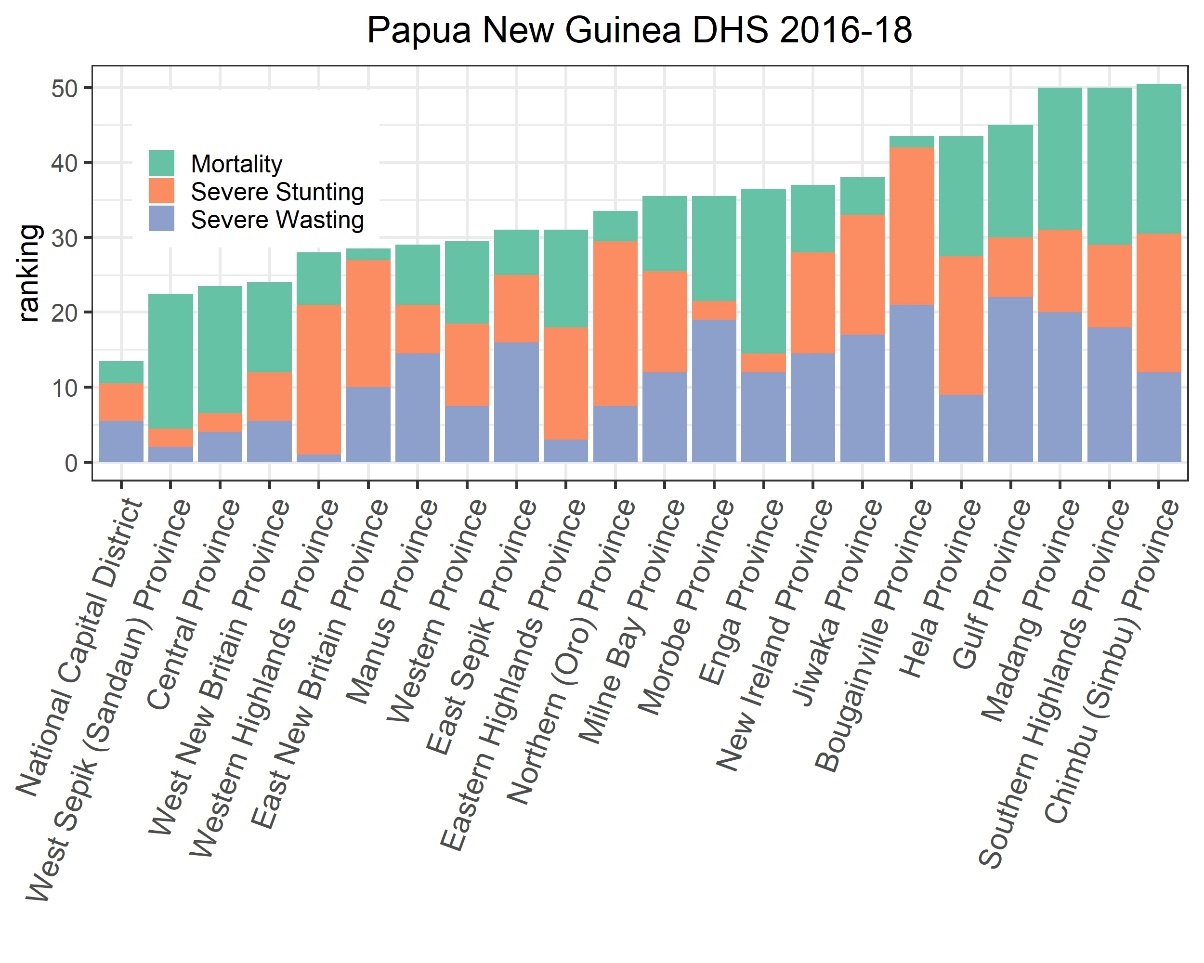

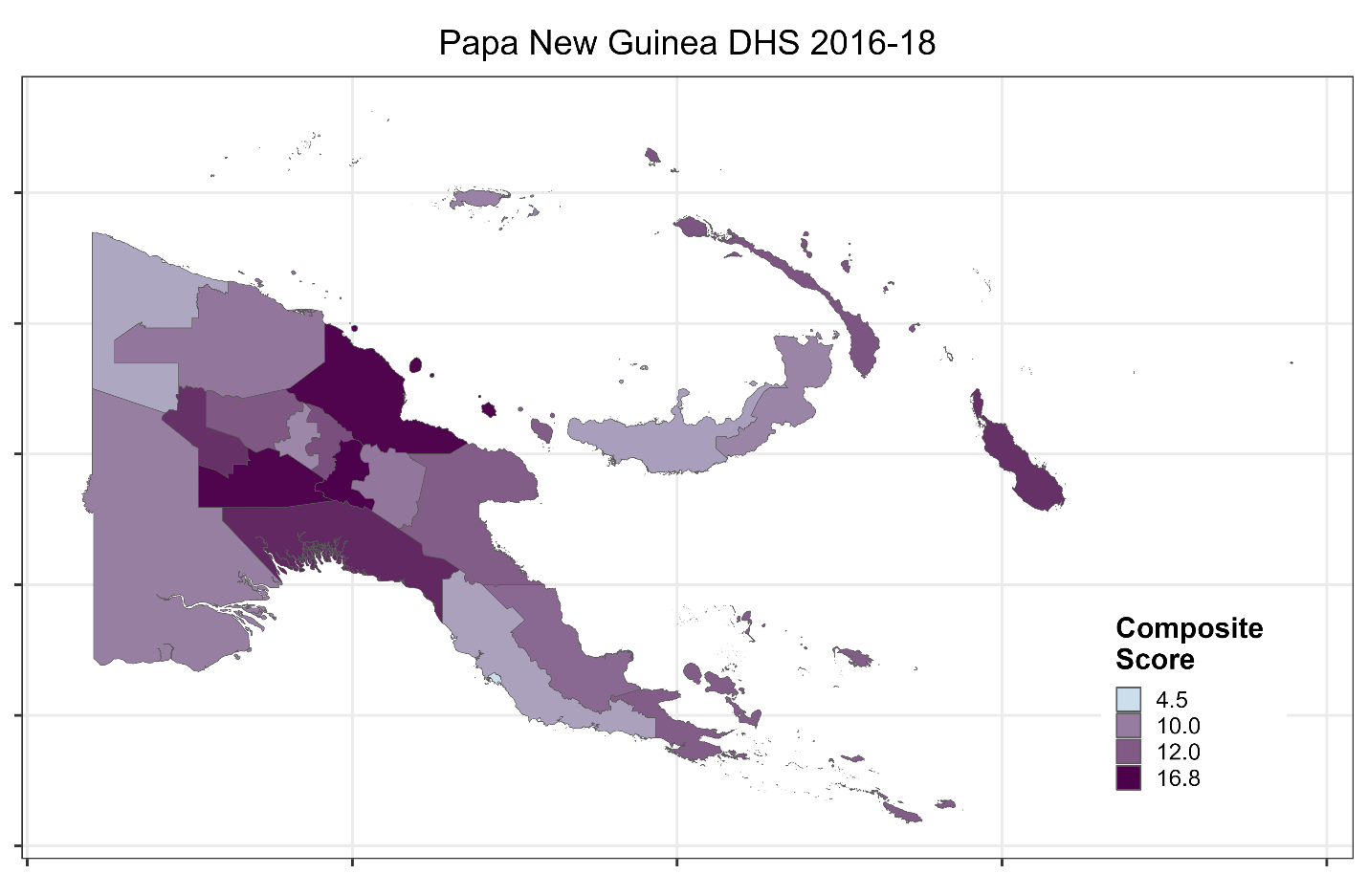

Figure S29. Sub-national ranking of Papua New Guinea by highest mortality rate and highest prevalence of severe stunting and severe wasting as well as heatmap of composite score at the sub-national level

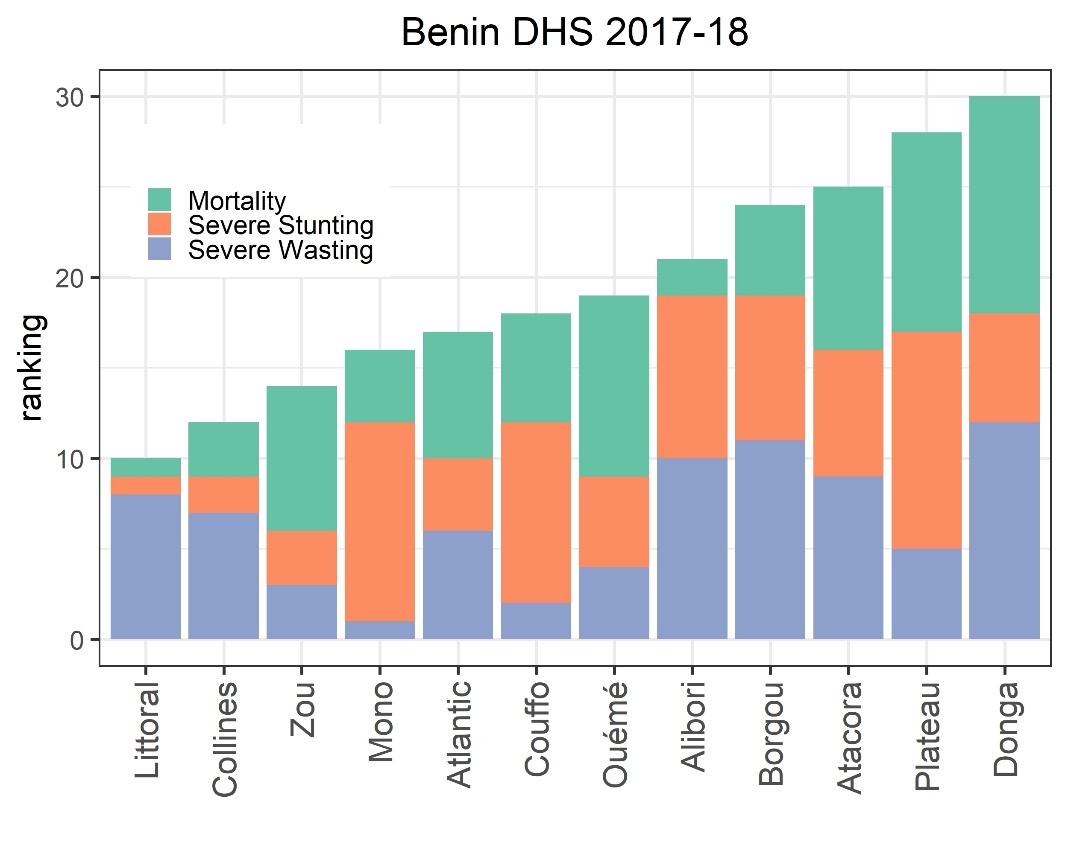

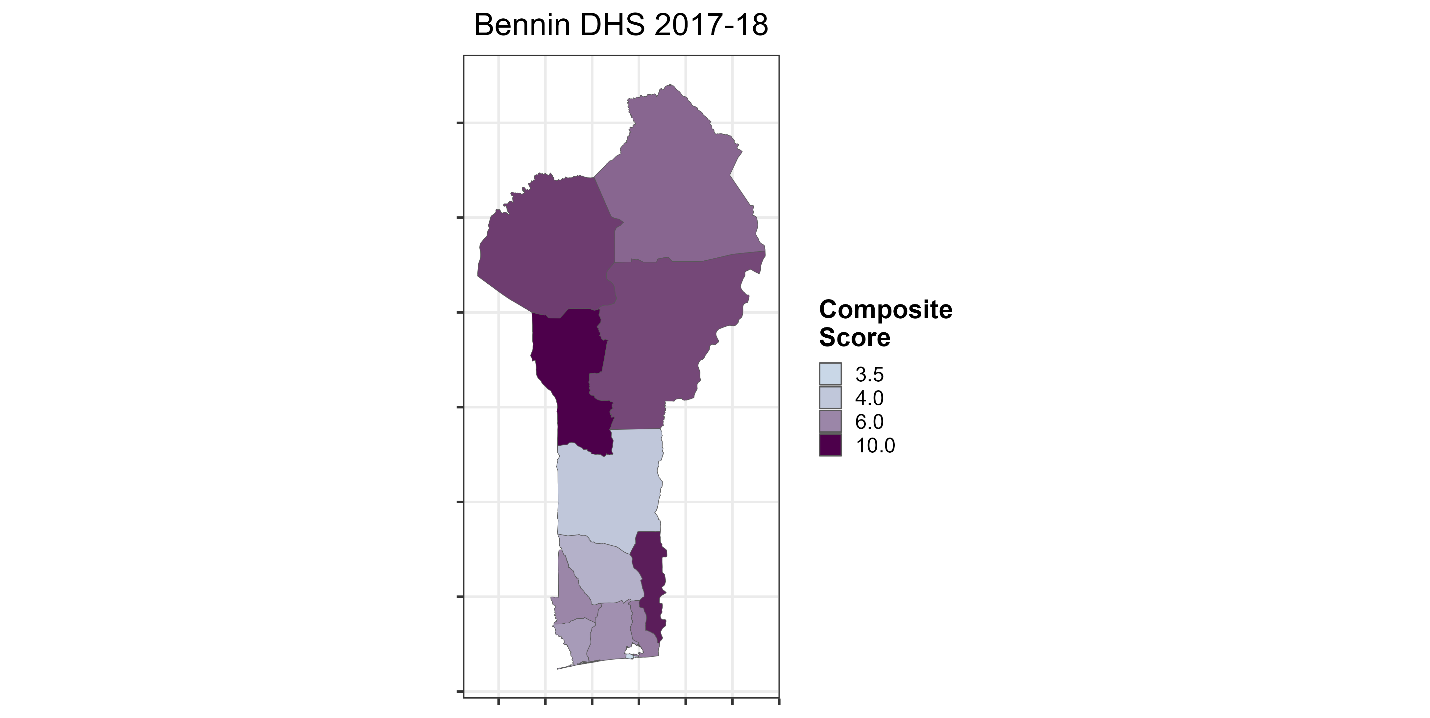

Figure S30. Sub-national ranking of Benin by highest mortality rate and highest prevalence of severe stunting and severe wasting as well as heatmap of composite score at the sub-national level

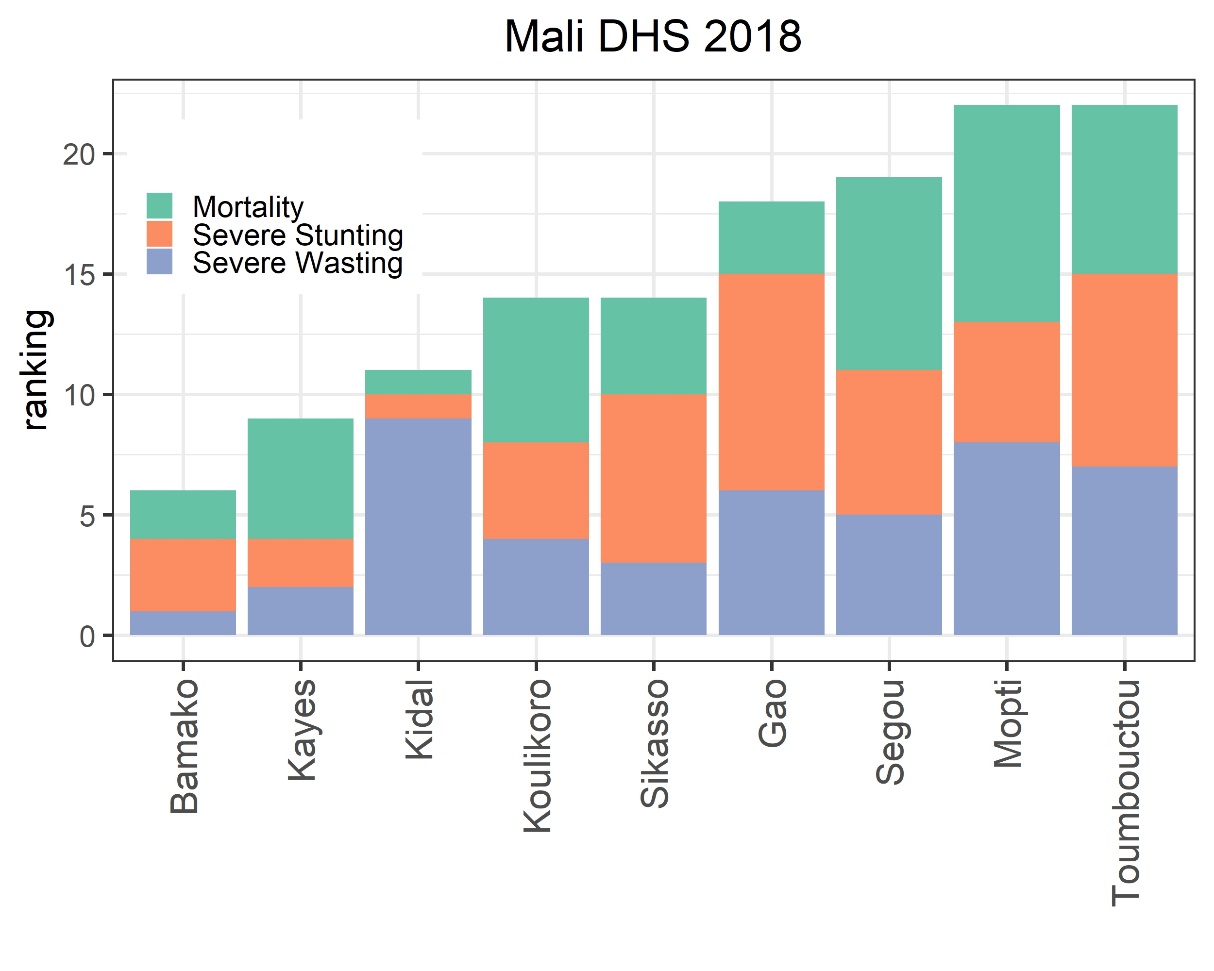

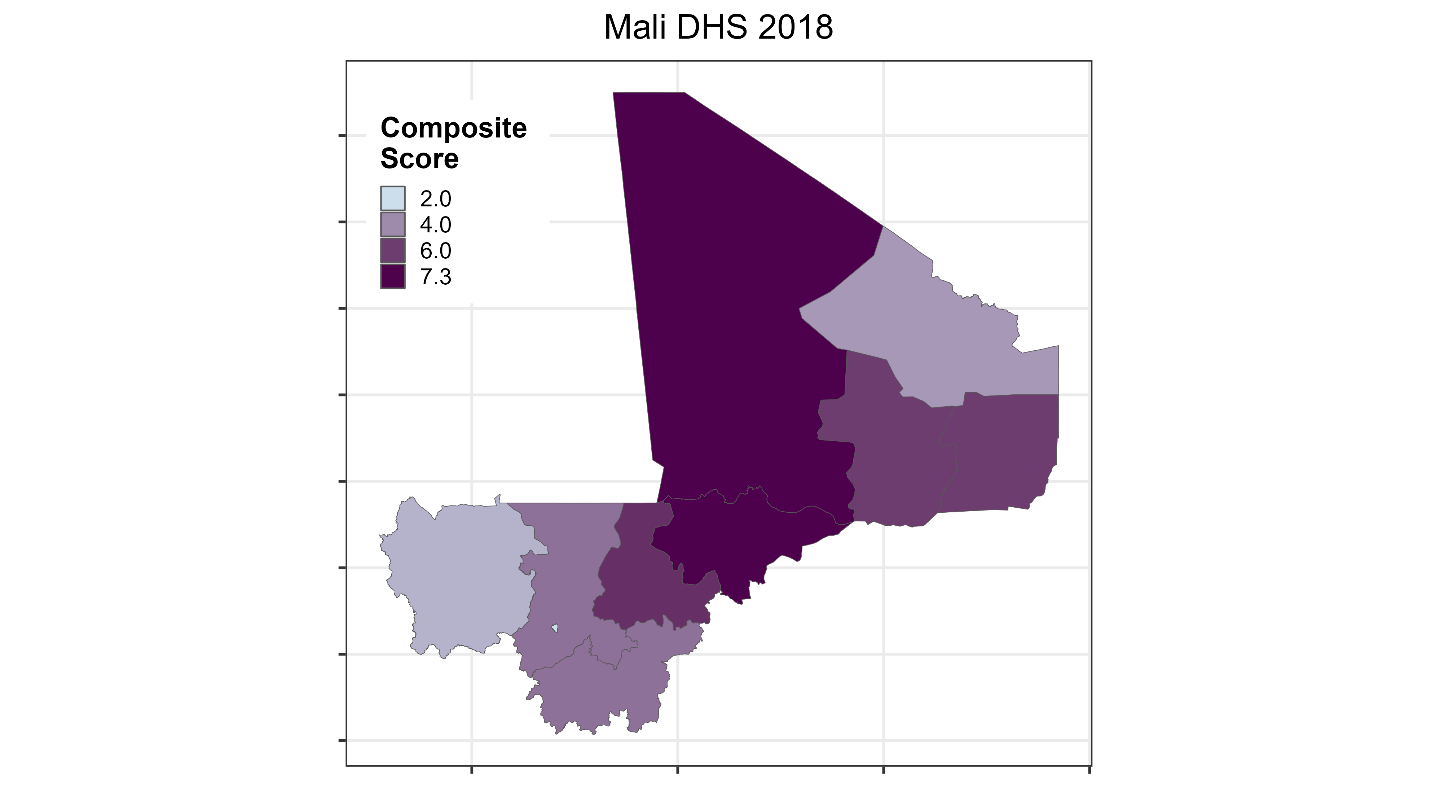

Figure S31. Sub-national ranking of Mali by highest mortality rate and highest prevalence of severe stunting and severe wasting as well as heatmap of composite score at the sub-national level

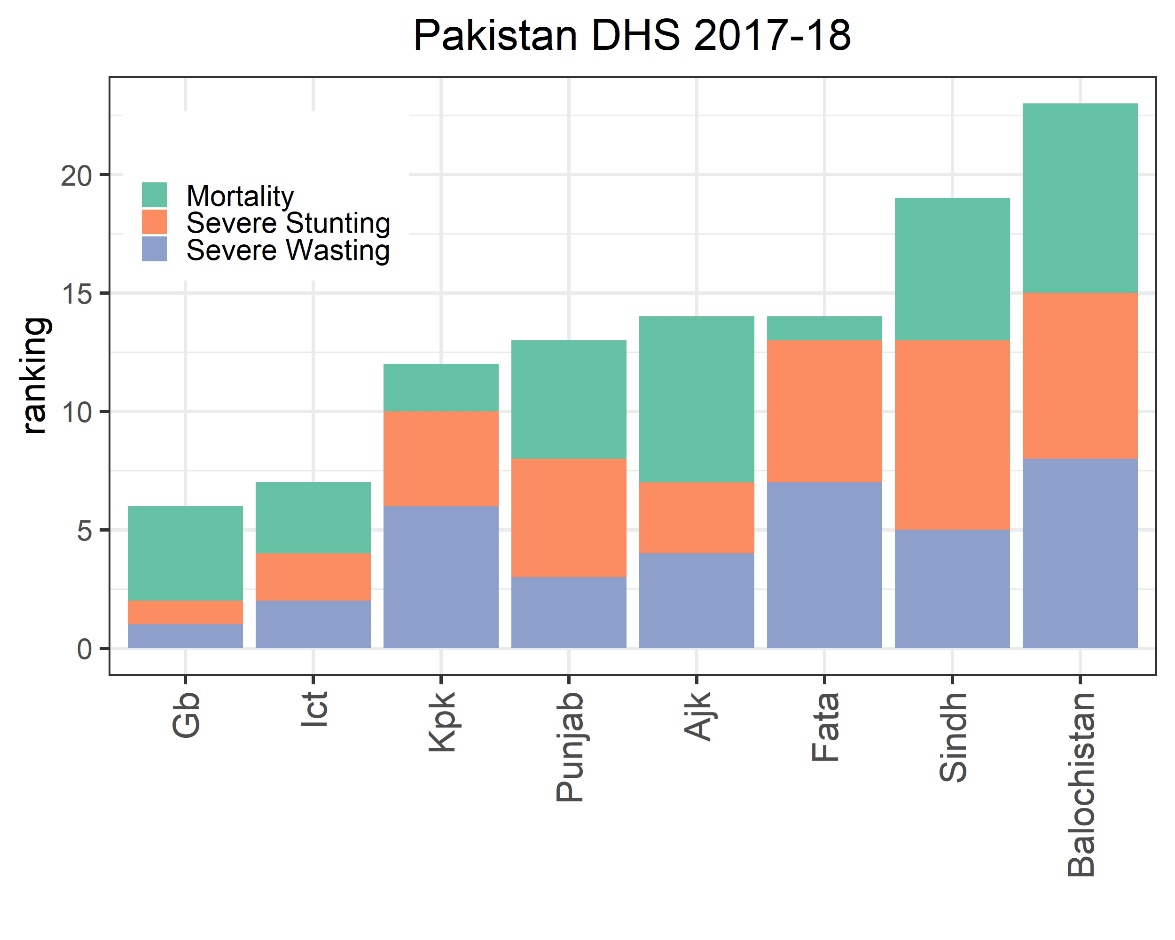

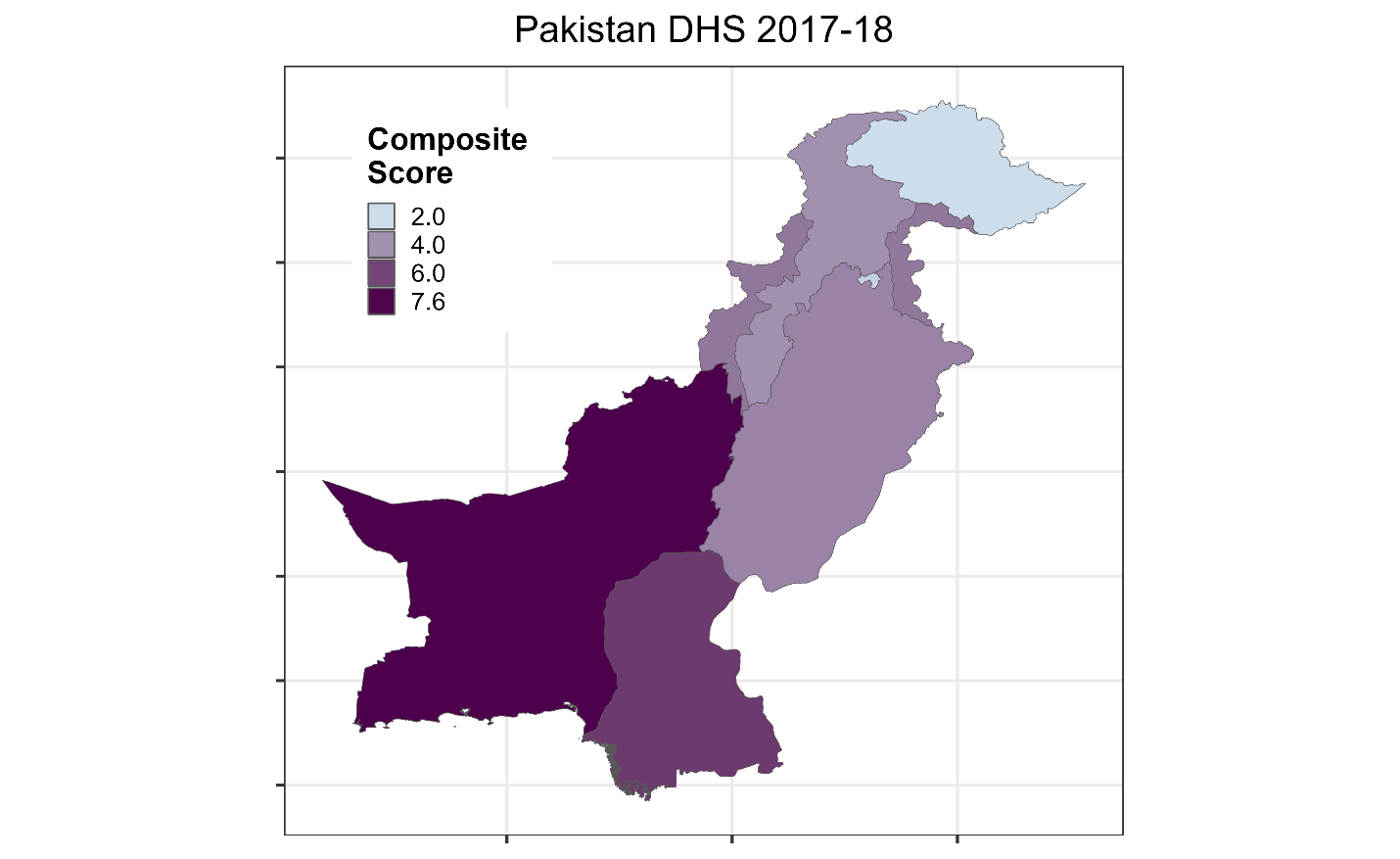

Figure S32. Sub-national ranking of Pakistan by highest mortality rate and highest prevalence of severe stunting and severe wasting as well as heatmap of composite score at the sub-national level

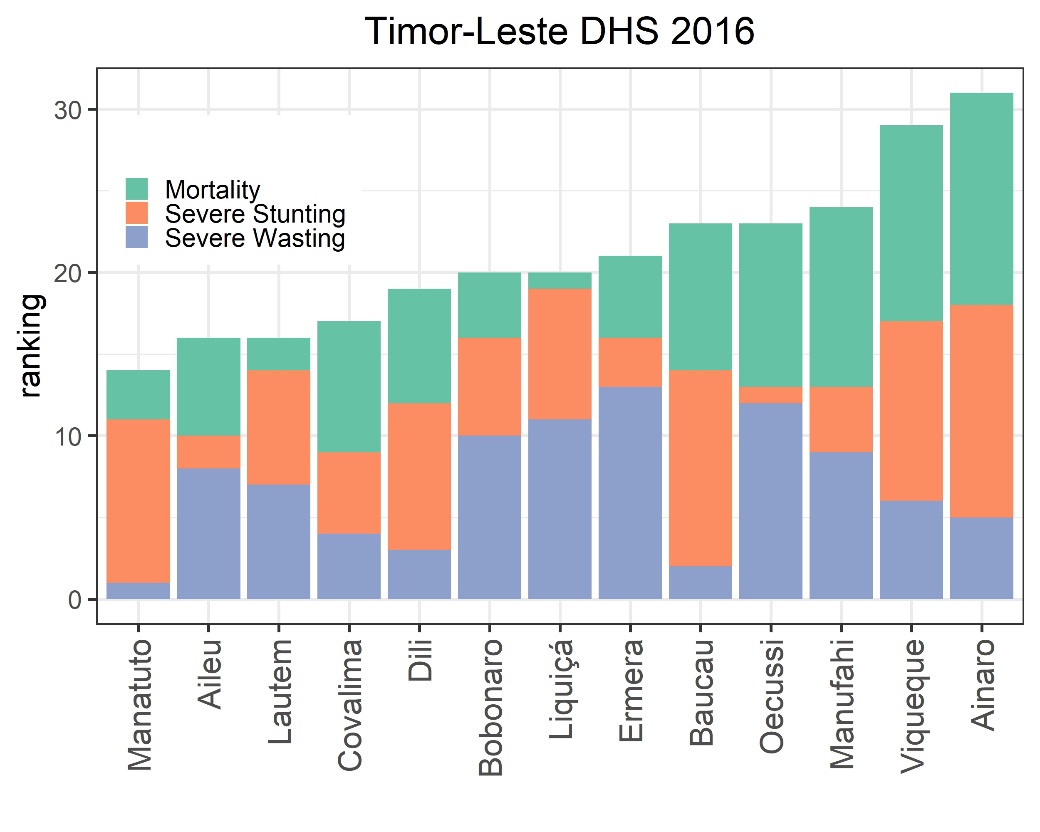

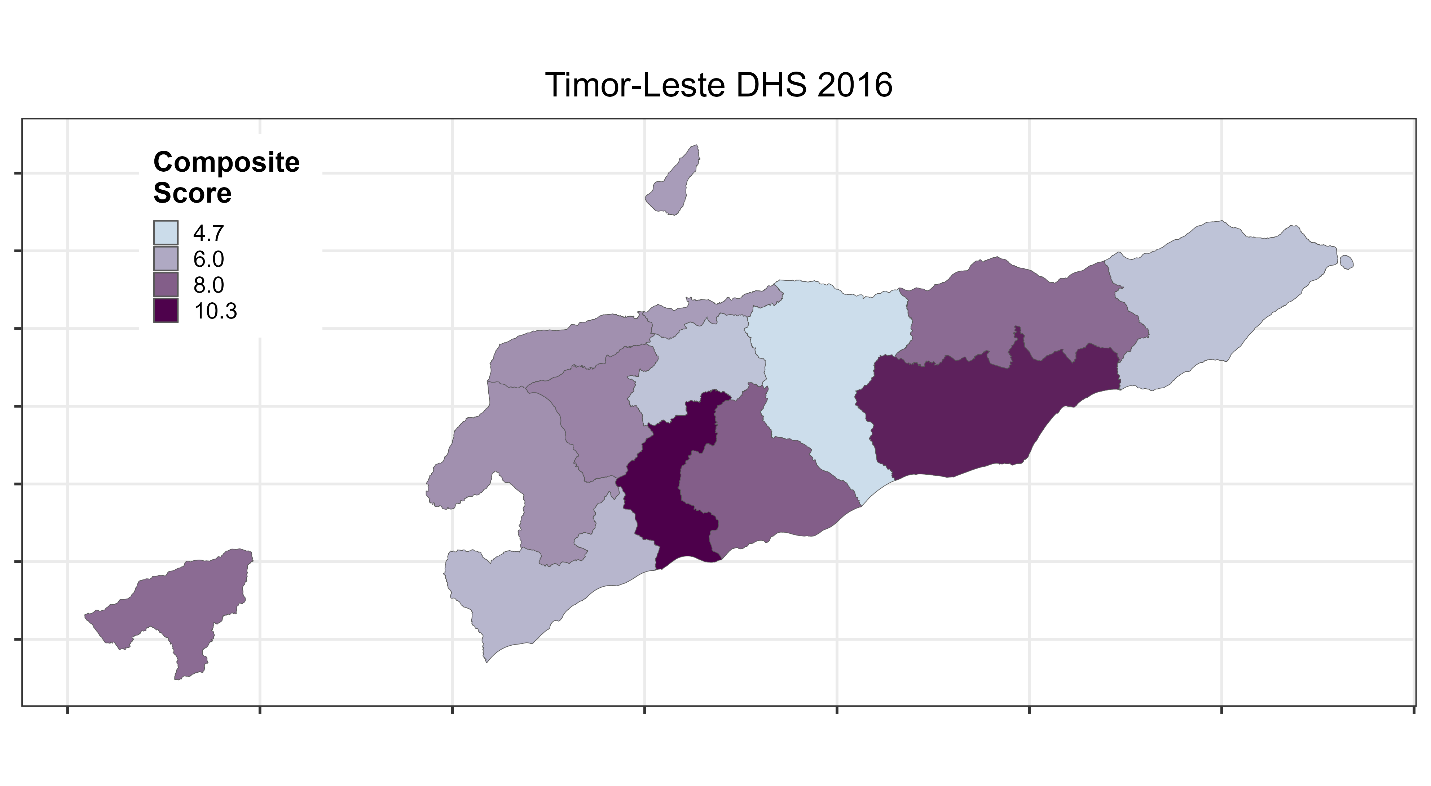

Figure S33. Sub-national ranking of Timor-Leste by highest mortality rate and highest prevalence of severe stunting and severe wasting as well as heatmap of composite score at the sub-national level

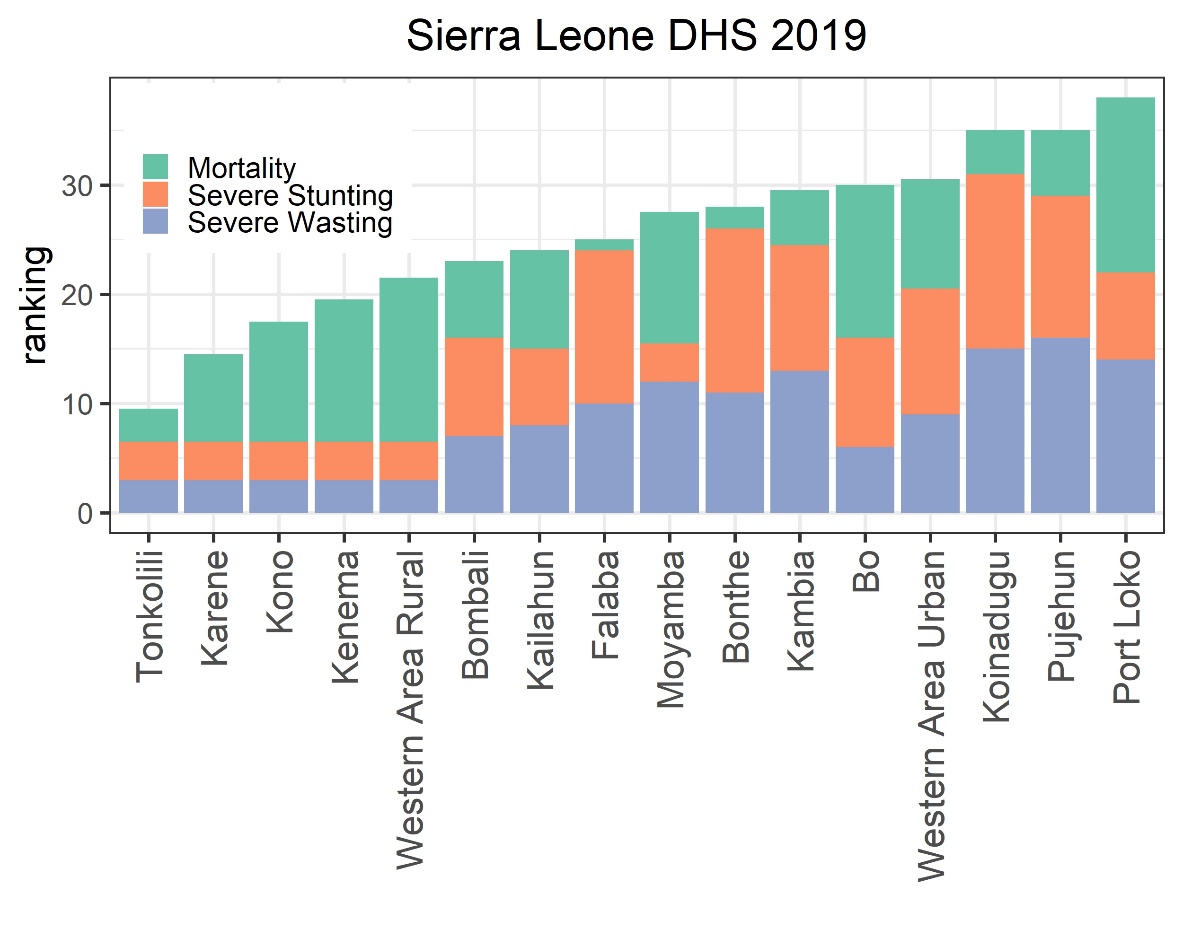

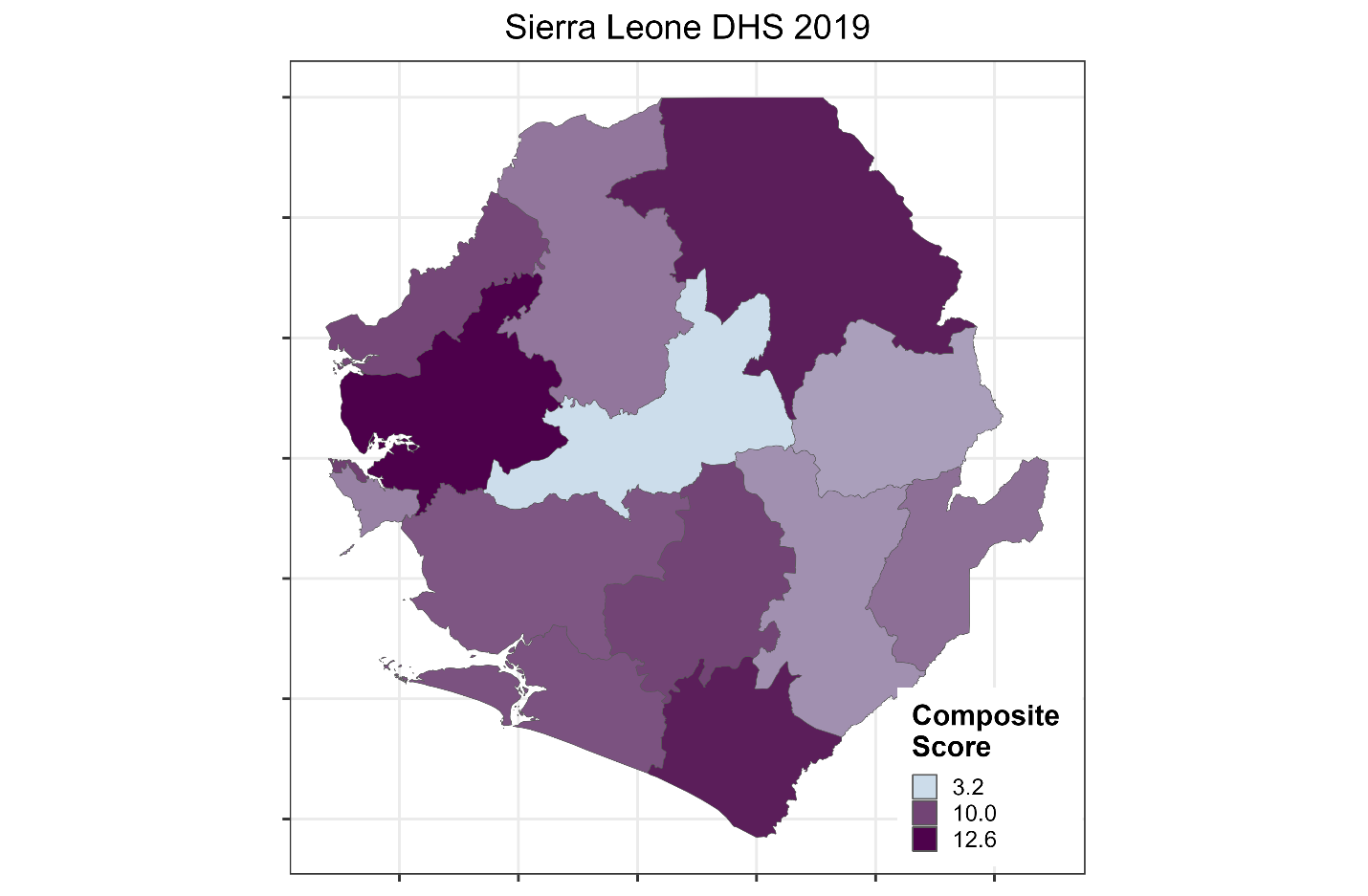

Figure S34. Sub-national ranking of Sierra Leone by highest mortality rate and highest prevalence of severe stunting and severe wasting as well as heatmap of composite score at the sub-national level

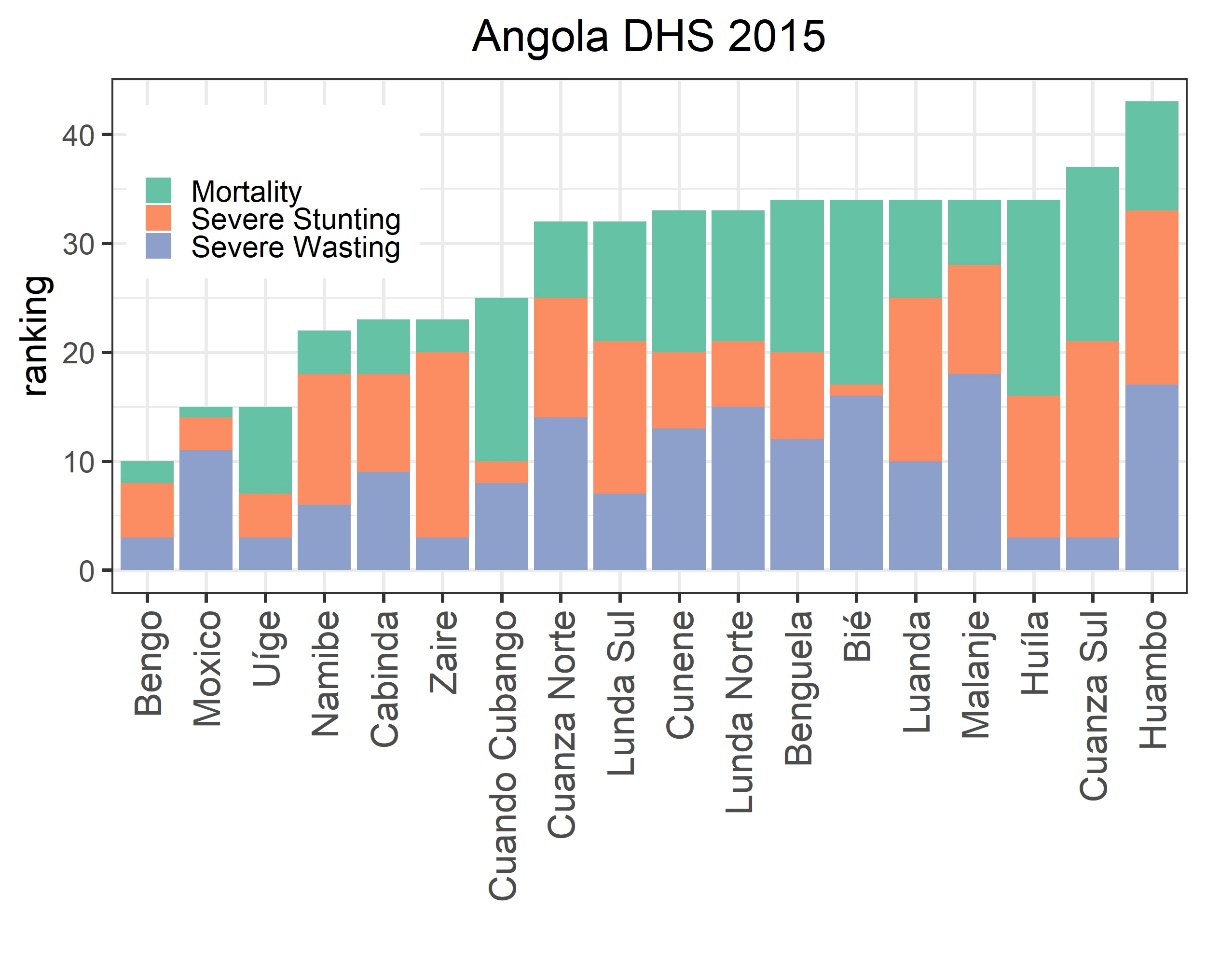

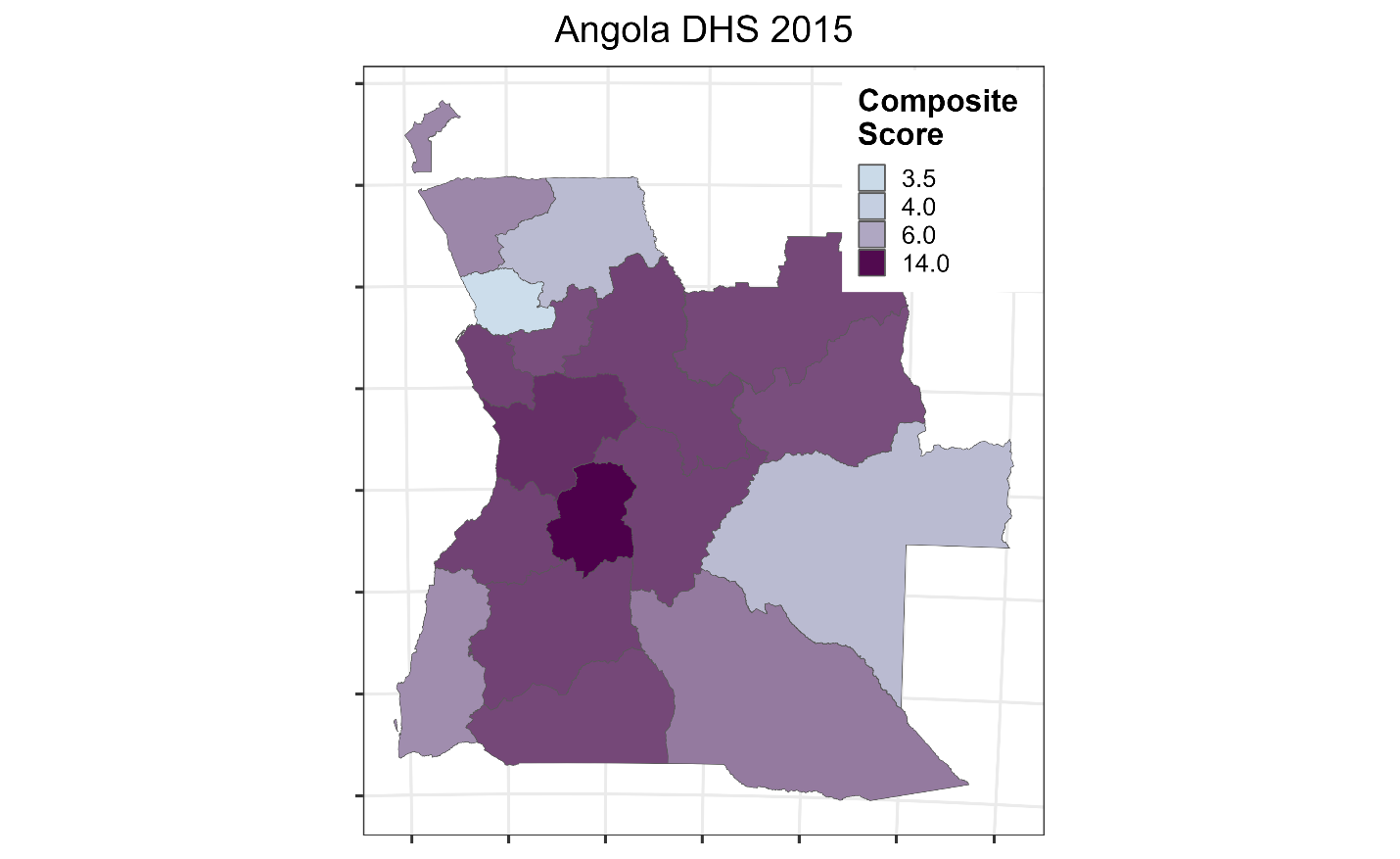

Figure S35. Sub-national ranking of Angola by highest mortality rate and highest prevalence of severe stunting and severe wasting as well as heatmap of composite score at the sub-national level

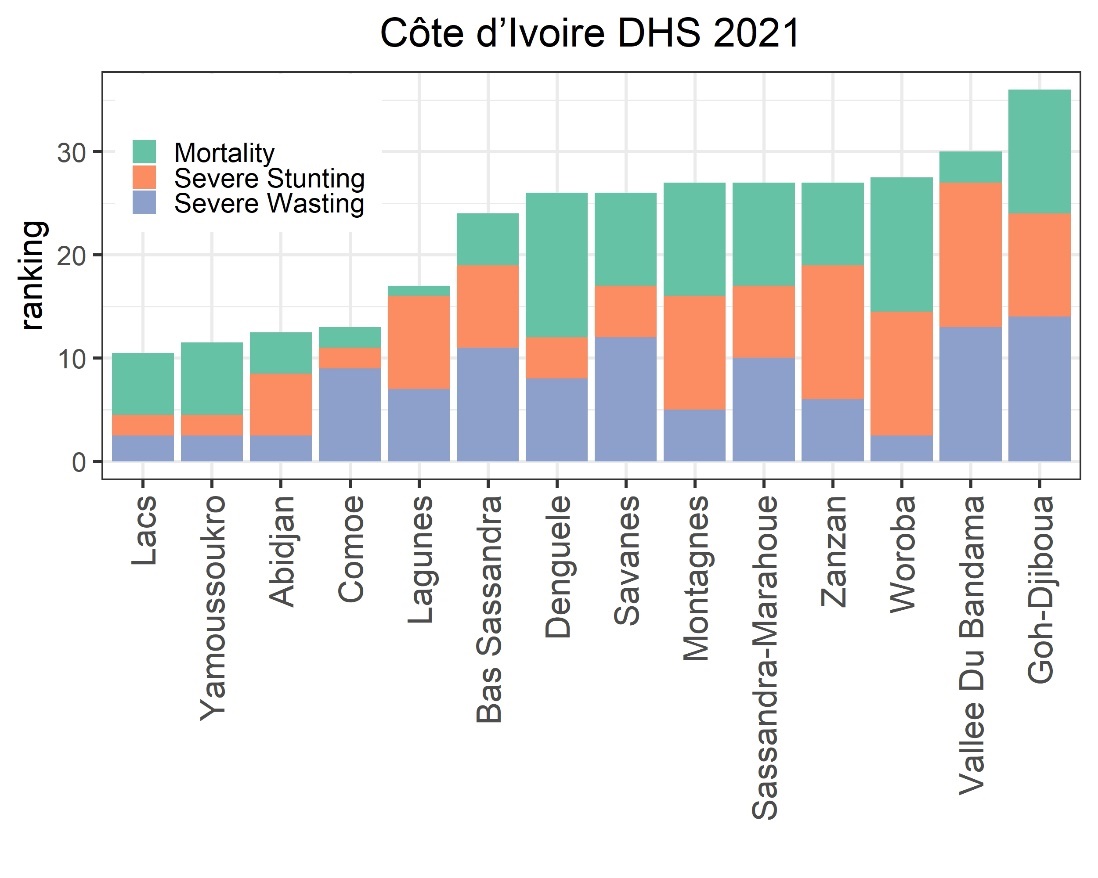

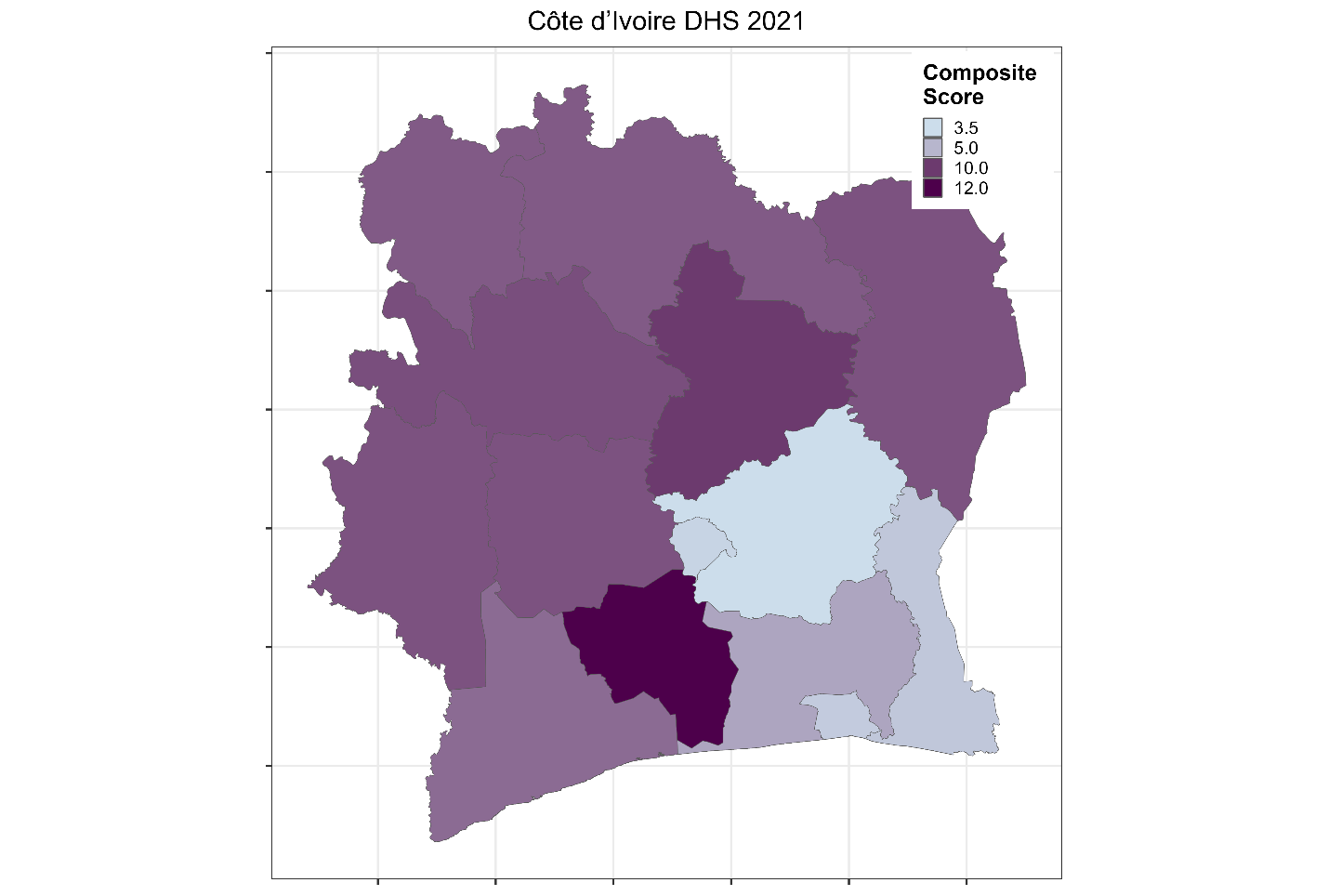

Figure S36. Sub-national ranking of Côte d’Ivoire by highest mortality rate and highest prevalence of severe stunting and severe wasting as well as heatmap of composite score at the sub-national level
